# Gut microbiome and metabolome reveal hormone-related and functional alterations in ER-positive breast cancer: a case–control study

**DOI:** 10.64898/2026.02.06.26345778

**Authors:** Ashley H Larnder, Kristin Campbell, Thaddeus J Edens, Julian J Lum, David R Goodlett, Jun Han, Kathryn Isaac, Rebecca Warburton, Michelle Goecke, Allen Hayashi, Alison Ross, Benoît Chassaing, Maeva Duquesnoy, Jessica Morgan, Jane Shearer, Parveen Bhatti, Amee R Manges, Rachel A Murphy

**Author notes:** Corresponding Author Rachel Murphy, 2206 East Mall, Vancouver, BC, V6T 1Z3. co-senior authors (Amee R Manges, Rachel A Murphy).

## Abstract

The gut microbiome has been linked to breast cancer, largely through microbial functions involved in estrogen metabolism (the “estrobolome”); however, specific microbial targets remain poorly defined in human studies. Here, we profiled the gut microbiome using whole-metagenome shotgun sequencing, and plasma and stool metabolites were quantified using targeted metabolomics, in a study of 70 postmenopausal female cases with treatment-naïve ER-positive breast cancer and 70 controls. Reduced species-level alpha and beta diversity were associated with breast cancer, whereas microbial functional-level diversity was not. Higher levels of DHEA-sulfate, estriol, and isoflavone phytoestrogen metabolites and lower lignan phytoestrogen metabolites were associated with breast cancer, while circulating estrogens and estrogen-related microbial functions were not. Beyond hormone-related pathways, higher levels of conjugated bile acids and carnitines were also associated with breast cancer. Compared to controls, cases exhibited depletion of *Blautia obeum, Alistipes shahii*, *A. finegoldii*, *A. putredinis*, and *Anaerotruncus rubiinfantis*, along with reduced abundance of functions related to menaquinol-8 biosynthesis, TCA cycle-related energy metabolism, and NAD salvage, indicating depletion of specific metabolic pathways rather than overall functional diversity. These results indicate a potential etiologic role for host–microbe metabolic interactions in breast cancer that extends beyond estrogen-centered mechanisms and warrants validation in independent cohorts.

## Introduction

Breast cancer is the most common cancer among females worldwide and a leading cause of cancer mortality.^1^ The most prevalent subtype, estrogen receptor–positive (ER+) breast cancer, highlights the central role of estrogen in disease etiology. While multiple potentially modifiable risk factors have been identified, including hormone exposure, dietary intake, body weight, physical activity, and alcohol intake,^2–4^ a large proportion of breast cancer risk remains unexplained, highlighting the need to better understand disease etiology and identify opportunities for prevention.

The gut microbiome has been increasingly implicated in human health and disease.^5^ The estrobolome hypothesis proposes that gut microbes capable of metabolizing estrogens may influence systemic estrogen levels and, in turn, ER+ breast cancer risk.^5^ Observational studies have reported taxa that are differentially enriched or depleted in breast cancer cases compared with controls,^6–14^ although substantial limitations remain. Many studies do not stratify by breast cancer subtype and rely on low-resolution 16S rRNA gene sequencing of bacterial composition which limits functional insight.^5^ Whole metagenome shotgun sequencing, on the other hand, supports functional profiling, which is critical for understanding whether compositional differences translate into meaningful biochemical effects on metabolism, immunity, and digestion.^5^

Metabolomics complements metagenomic sequencing by capturing the inputs and outputs of host and microbial pathways, providing additional insight into biologically relevant processes.^5^ Few observational studies of the gut microbiome, estrogen and breast cancer have integrated metabolomics with microbiome analyses.^15–22^ Moreover, metabolite profiles reflect downstream products of both microbial activity and dietary substrates. Diet modulates microbial composition and metabolite production, and many microbial metabolites rely entirely on microbial metabolism for activation into bioactive forms, such as phytoestrogens into equol and enterolignans.^22–24^

Experimental evidence has suggested that targeting the estrobolome may be a novel strategy for breast cancer prevention;^5^ however, a better understanding of the role of the estrobolome in breast cancer is needed. Here, we apply high-resolution metagenomic sequencing and targeted metabolomics, alongside dietary assessment, to investigate the role of estrogen-related microbial and metabolic features in females with treatment-naïve breast cancer. Specifically, we examine whether estrobolome targets—including microbial taxa, functions, and metabolites—are associated with breast cancer, whether additional microbial and metabolic features are linked to disease, and how integration of metagenomic and metabolomic data can provide insight into relevant pathways.

## Results

### Participant characteristics

Cases and controls were broadly similar with respect to most characteristics (**Table 1**), with mean age of 61 years and BMI of 24 kg/m^2^. A higher proportion of cases identified as non-White compared with controls (48% vs 17%); non-White participants included East Asian (n=19), Filipino/South Asian (n=9), and other groups (n=17). Cases reported lower use of menopausal hormone therapy, both for ever use (32% vs 41%) and current use (12% vs 26%).

**Table 1.**
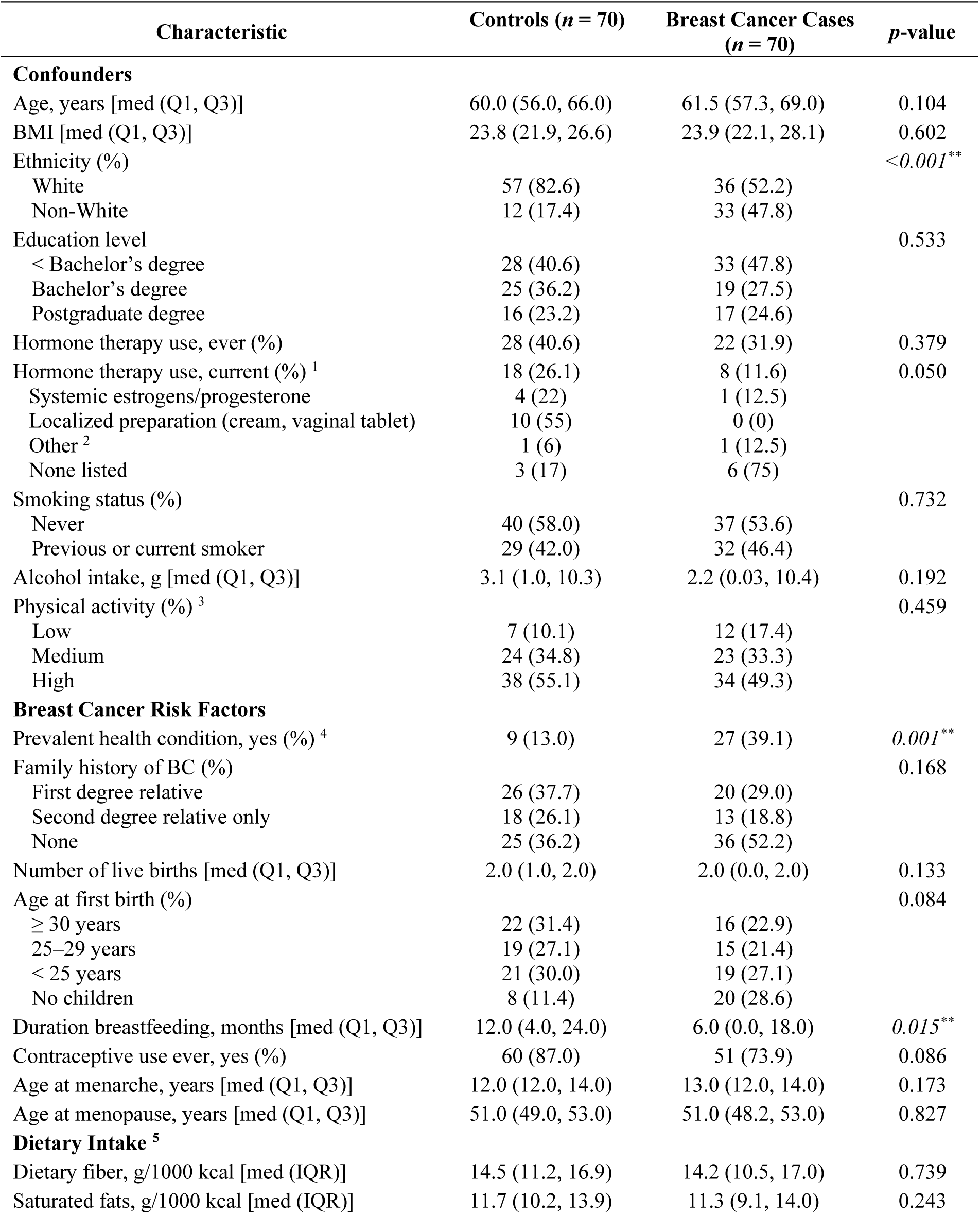
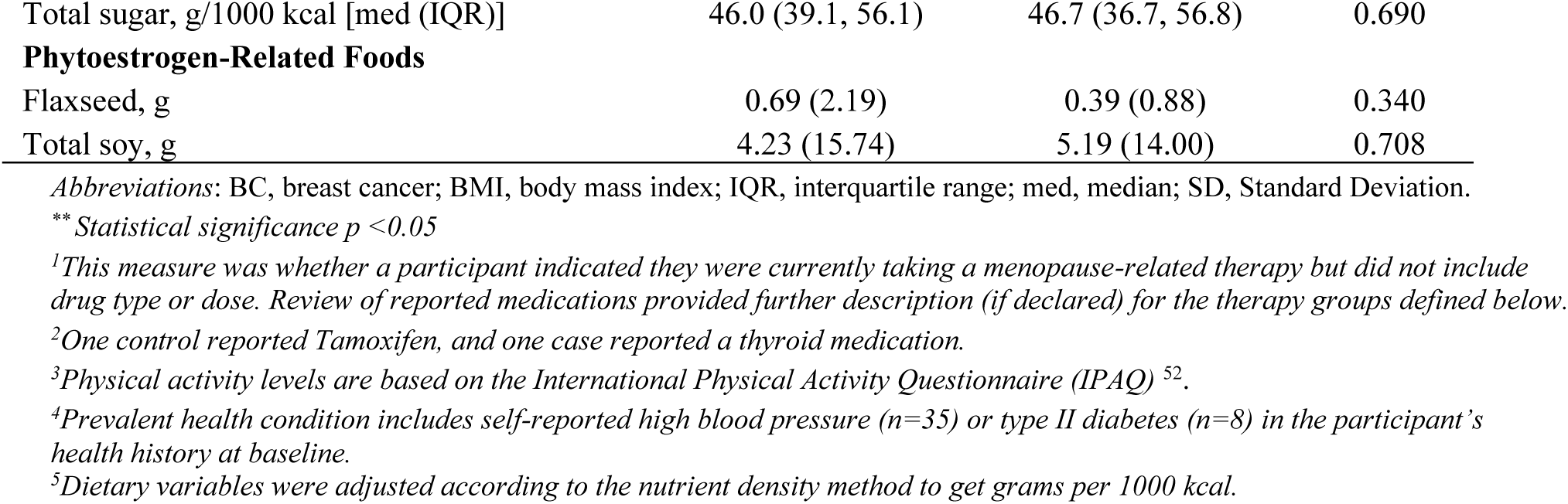
Descriptive characteristics of postmenopausal females in the study sample (*n* = 140).

| Characteristic | Controls ($n = 70$) | Breast Cancer Cases ($n = 70$) | $p$-value |
| --- | --- | --- | --- |
| **Confounders** |  |  |  |
| Age, years [med (Q1, Q3)] | 60.0 (56.0, 66.0) | 61.5 (57.3, 69.0) | 0.104 |
| BMI [med (Q1, Q3)] | 23.8 (21.9, 26.6) | 23.9 (22.1, 28.1) | 0.602 |
| Ethnicity (%) | | | $<0.001^{**}$ |
| White | 57 (82.6) | 36 (52.2) |  |
| Non-White | 12 (17.4) | 33 (47.8) |  |
| Education level |  |  | 0.533 |
| < Bachelor's degree | 28 (40.6) | 33 (47.8) |  |
| Bachelor's degree | 25 (36.2) | 19 (27.5) |  |
| Postgraduate degree | 16 (23.2) | 17 (24.6) |  |
| Hormone therapy use, ever (%) | 28 (40.6) | 22 (31.9) | 0.379 |
| Hormone therapy use, current (%) <sup>1</sup> | 18 (26.1) | 8 (11.6) | 0.050 |
| Systemic estrogens/progesterone | 4 (22) | 1 (12.5) |  |
| Localized preparation (cream, vaginal tablet) | 10 (55) | 0 (0) |  |
| Other <sup>2</sup> | 1 (6) | 1 (12.5) |  |
| None listed | 3 (17) | 6 (75) |  |
| Smoking status (%) |  |  | 0.732 |
| Never | 40 (58.0) | 37 (53.6) |  |
| Previous or current smoker | 29 (42.0) | 32 (46.4) |  |
| Alcohol intake, g [med (Q1, Q3)] | 3.1 (1.0, 10.3) | 2.2 (0.03, 10.4) | 0.192 |
| Physical activity (%) <sup>3</sup> |  |  | 0.459 |
| Low | 7 (10.1) | 12 (17.4) |  |
| Medium | 24 (34.8) | 23 (33.3) |  |
| High | 38 (55.1) | 34 (49.3) |  |
| **Breast Cancer Risk Factors** |  |  |  |
| Prevalent health condition, yes (%) <sup>4</sup> | 9 (13.0) | 27 (39.1) | $0.001^{**}$ |
| Family history of BC (%) |  |  | 0.168 |
| First degree relative | 26 (37.7) | 20 (29.0) |  |
| Second degree relative only | 18 (26.1) | 13 (18.8) |  |
| None | 25 (36.2) | 36 (52.2) |  |
| Number of live births [med (Q1, Q3)] | 2.0 (1.0, 2.0) | 2.0 (0.0, 2.0) | 0.133 |
| Age at first birth (%) |  |  | 0.084 |
| ≥ 30 years | 22 (31.4) | 16 (22.9) |  |
| 25–29 years | 19 (27.1) | 15 (21.4) |  |
| < 25 years | 21 (30.0) | 19 (27.1) |  |
| No children | 8 (11.4) | 20 (28.6) |  |
| Duration breastfeeding, months [med (Q1, Q3)] | 12.0 (4.0, 24.0) | 6.0 (0.0, 18.0) | $0.015^{**}$ |
| Contraceptive use ever, yes (%) | 60 (87.0) | 51 (73.9) | 0.086 |
| Age at menarche, years [med (Q1, Q3)] | 12.0 (12.0, 14.0) | 13.0 (12.0, 14.0) | 0.173 |
| Age at menopause, years [med (Q1, Q3)] | 51.0 (49.0, 53.0) | 51.0 (48.2, 53.0) | 0.827 |
| **Dietary Intake <sup>5</sup>** |  |  |  |
| Dietary fiber, g/1000 kcal [med (IQR)] | 14.5 (11.2, 16.9) | 14.2 (10.5, 17.0) | 0.739 |
| Saturated fats, g/1000 kcal [med (IQR)] | 11.7 (10.2, 13.9) | 11.3 (9.1, 14.0) | 0.243 |
| Total sugar, g/1000 kcal [med (IQR)] | 46.0 (39.1, 56.1) | 46.7 (36.7, 56.8) | 0.690 |
| **Phytoestrogen-Related Foods** |  |  |  |
| Flaxseed, g | 0.69 (2.19) | 0.39 (0.88) | 0.340 |
| Total soy, g | 4.23 (15.74) | 5.19 (14.00) | 0.708 |

*Abbreviations:* BC, breast cancer; BMI, body mass index; IQR, interquartile range; med, median; SD, Standard Deviation.
*\*\* Statistical significance $p < 0.05$*
<sup>1</sup>*This measure was whether a participant indicated they were currently taking a menopause-related therapy but did not include drug type or dose. Review of reported medications provided further description (if declared) for the therapy groups defined below.*
<sup>2</sup>*One control reported Tamoxifen, and one case reported a thyroid medication.*
<sup>3</sup>*Physical activity levels are based on the International Physical Activity Questionnaire (IPAQ) <sup>52</sup>.*
<sup>4</sup>*Prevalent health condition includes self-reported high blood pressure ( $n=35$ ) or type II diabetes ( $n=8$ ) in the participant's health history at baseline.*
<sup>5</sup>*Dietary variables were adjusted according to the nutrient density method to get grams per 1000 kcal.*

Among those reporting current use of hormone therapy (yes/no), only five participants listed systemic estrogens and/or progesterone in their reported current medications (controls=4, cases=1), 10 controls reported localized preparations (topical vaginal cream or vaginal tablet), and several listed no hormone-related medications despite indicating current use. Dietary intake was generally similar between cases and controls (**Supplementary Table 1**).

Among breast cancer cases, all tumors were estrogen receptor-positive (100%), as per study inclusion criterion, 77% were progesterone receptor-positive, and 10% were HER2-positive. Most cancers were Stage I (58%) and grade II (50%). Most tumors were unifocal (82%) with a median size of 15.0 mm (IQR 14.0 mm).

### Metagenome sequencing performance

On average, 41.9 million quality-filtered reads were generated per sample (range = 16.5–62.2 million; SD = 8.1 million). The mean percentage of non-annotated reads per sample was 21.4% (±2.6%). Thirteen negative controls yielded a mean of 1,027 quality-filtered reads (range = 171–2,440; SD = 565). Samples from three unique participants, collected at both baseline and follow-up, were extracted and sequenced in triplicate, resulting in a total of 18 samples to assess technical variation. Bray-Curtis distances among replicate groups were low, ranging from 0.022 to 0.045, and principal coordinate analysis (PCoA) confirmed minimal variation between replicates (**Supplementary Fig. 1**). Genus-level relative abundances similarly demonstrated high reproducibility and minor variation across replicates (**Supplementary Fig. 1**).

### The overall gut microbiome differed in composition but not function between cases and controls

After prevalence and abundance threshold filtering, 631 species were identified. All were bacterial species except for two archaeal species (*Methanobrevibacter smithii and Nitrosopumilus* SGB14899). *Faecalibacterium prausnitzii* was predominant, followed by *Ruminococcus bromii, Phocaeicola vulgatus, Blautia wexlerae,* and *Eubacterium rectale.* Cases had fewer species (n=197) compared to controls (n=247), which contributed to a significantly lower α-diversity (**Fig. 1**): richness measure (p=0.0009) and Shannon index (p=0.005). Species differences were also apparent when examining β-diversity (PERMANOVA; R^2^=0.013; p=0.015; **Fig. 1**).

**Fig. 1.**
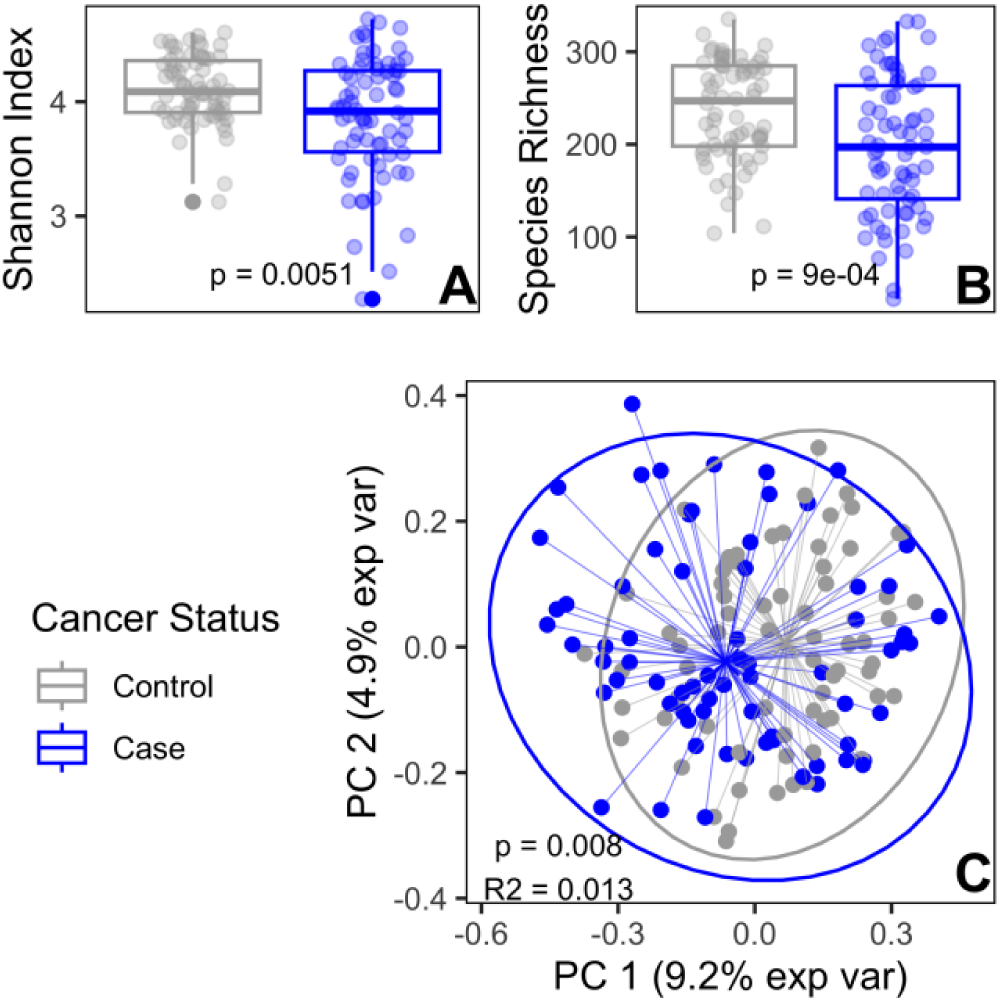
Diversity measures of species-level taxonomic data. Alpha diversity by (A) Shannon index and (B) species richness, and (C) beta diversity using Bray-Curtis dissimilarity visualized by principal coordinate analysis (PCoA), showed species to be significantly different between cases and controls (p<0.05).

A total of 452 metabolic pathways and 2,102 enzymes were identified, with the most abundant features including sucrose biosynthesis II (PWY-7238), glycolysis IV (PWY-1042), histidine kinase (EC 2.7.13.3), and DNA-directed RNA polymerase (EC 2.7.7.6). Neither pathways nor enzymes differed significantly in α- or β-diversity between cases and controls (**Supplementary Fig. 2**).

### Established estrobolome targets showed limited case–control differences

Microbial features linked to estrogen-related functions (“estrobolome targets”) identified in the filtered data included 10 genera, 39 unique species (54 with subspecies variation), one pathway, and 6 enzymes (**Supplementary Table 2**). Several features were statistically significant in unadjusted models (Model 0); however, after adjustment for potential confounders (Model 1), these associations had smaller effect sizes (narrower x-axis distribution, **Fig. 2**) and none remained significant after FDR correction. In Model 1, nine features still showed marginal associations (p<0.05), including one genus (*Ruminococcus*) and eight species: five species linked to β-glucuronidase/β-glucosidase activity (*L. eligens*, *B. bifidum*, *C. butyricum*, *E. clostridioformis, E. ramosum*), two *Blautia obeum* sub-species linked to β-glucosidase activity, and one species linked to equol production (*Adlercreutzia equolifaciens*). Most were associated with lower odds of breast cancer, except *C. butyricum E. clostridioformis* and *E. ramosum*, which were associated with higher odds. Further adjustment for dietary variables (Model 2) had little impact on model estimates relative to Model 1 estimates (**Supplementary Table 2**).

**Fig. 2.**
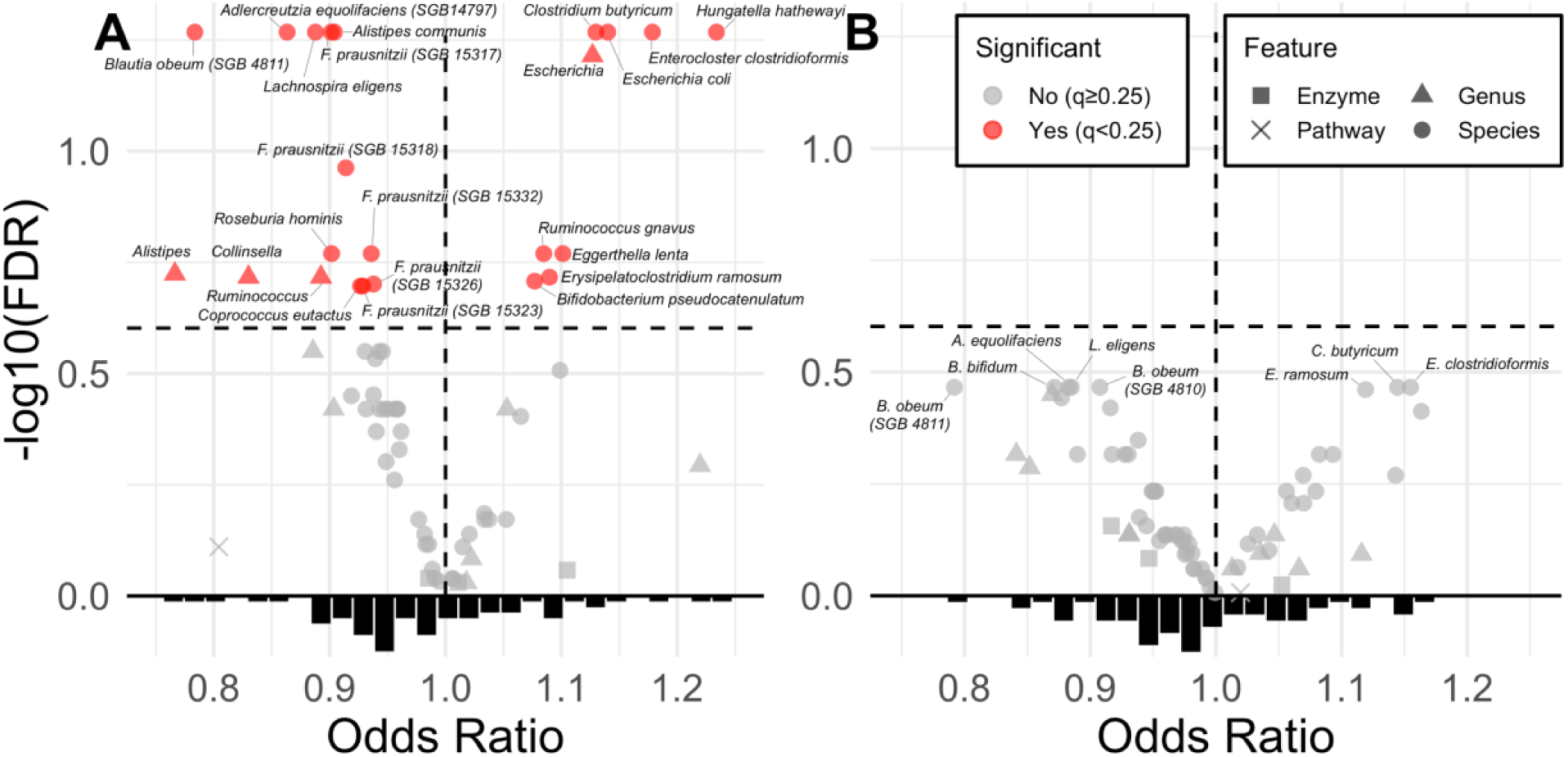
Volcano plots of logistic regression results for estrobolome targets. A: unadjusted (Model 0), B: confounder-adjusted (Model 1: age, BMI, ethnicity, education, current hormone therapy use, smoking status, alcohol intake, physical activity). Shapes indicate feature type (circle = species, square = genus, triangle = enzyme, diamond = pathway). FDR-corrected p-values (*q*-values) were plotted as –log10(*q*-value). Odds ratios were truncated at 1.25 for visualization; three enzymes exceeded this range but were not significant.

### Broader metagenomic features were predictive of breast cancer

XGBoost model performance varied by metagenomic feature types (**Supplementary Table 3**). Enzymes showed the strongest discrimination of breast cancer status (pseudo-R^2^ 29.7%, AUC 0.82), followed by species (pseudo-R^2^ 13.6%, AUC 0.71), and pathways (pseudo-R^2^ 7.4%, AUC 0.66). Several biologically relevant features ranked highly in the XGBoost models (**Fig. 3**).

**Fig. 3.**
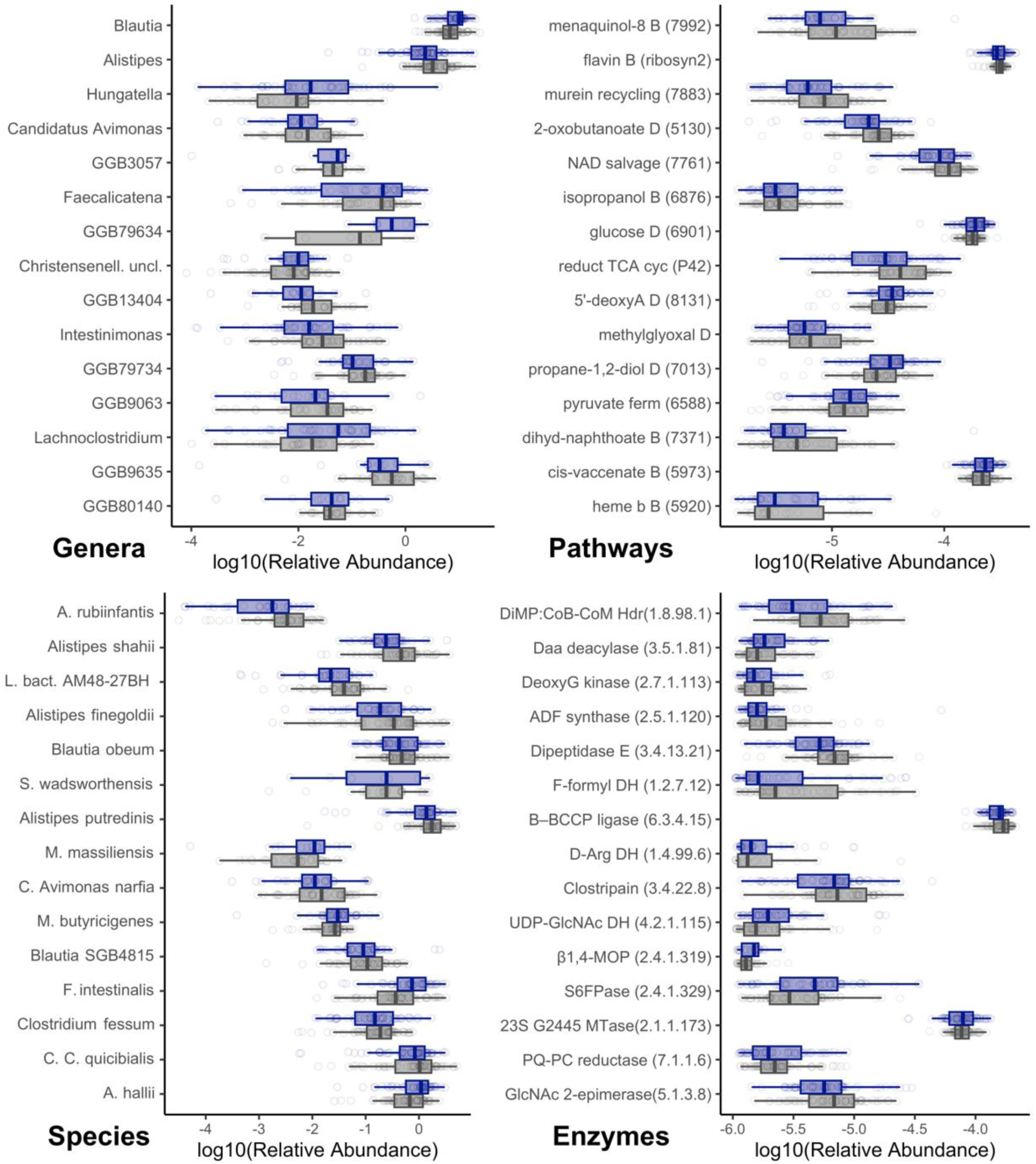
Boxplots of the relative abundance of top features discriminating breast cancer status in XGBoost. Shown by features type (genus, species, pathways, and enzymes), with cases in blue and controls in grey. The full feature list is available in **Supplementary Table 3**. Pathway abbreviations: D = degradation, B = biosynthesis. Enzyme abbreviations: DH = dehydrogenase, Pase = phosphorylase, MTase = methyltransferase.

To provide directionality and effect-size estimates, logistic regression models were fit to the top XGBoost features (Model 1; **Supplementary Table 4**). Top discriminating genera included *Alistipes* and *Blautia*, and among species, *Anaerotruncus rubiinfantis* was a top discriminating species in XGBoost and was associated with lower breast cancer odds (OR=0.76, 95% CI: 0.64–0.89). Additional high-ranking species included *Blautia obeum* (SGB4811, OR=0.79), *Alistipes shahii* (OR=0.92), *A. finegoldii* (OR=0.92), and *A. putredinis* (OR=0.89).

Top discriminating pathways in XGBoost included vitamin and cofactor biosynthesis, including menaquinol-8 (PWY-7992; OR=0.86), 1,4-dihydroxy-6-naphthoate (PWY-7371; OR=0.84), heme b (PWY-5920; OR=1.12), NAD salvage II (PWY-7761; OR=0.44). Central carbon and energy-related pathways also ranked highly, including isopropanol biosynthesis (PWY-6876; OR=0.78), incomplete reductive TCA cycle (42-PWY; OR=0.53), and 2-oxobutanoate degradation (PWY-5130; OR=0.49).

Top discriminating enzymes included hydrolases such as N-acyl-D-amino-acid deacylase (EC 3.5.1.81; OR=1.37) and dipeptidase E (EC 3.4.13.21; OR=0.38); oxidoreductases including dihydromethanophenazine:CoB–CoM heterodisulfide reductase (EC 1.8.98.1; OR=0.50) and formylmethanofuran dehydrogenase (EC 1.2.7.12; OR=0.80); enzymes involved in cofactor biosynthesis such as aminodeoxyfutalosine synthase (EC 2.5.1.120; OR=0.85); and enzymes involved in carbohydrate degradation such as sucrose 6(F)-phosphate phosphorylase (EC 2.4.1.329; OR=1.37) and UDP-N-acetylglucosamine 4,6-dehydratase (EC 4.2.1.115; OR=1.22).

### Plasma metabolic signals differentiated breast cancer cases and controls

Logistic regression of 182 plasma metabolites (**Supplementary Table 5**) identified eight metabolites significantly associated with breast cancer odds in unadjusted analyses (Model 0) and 12 in adjusted analyses (Model 1; **Fig. 4A–B**). This included two steroid hormone-related metabolites (estriol, DHEA-sulfate), two bile acids (murocholic acid, hyocholic acid 3-sulfate), two carnitines (glutaryl-, 3-methylglutaryl-carnitine), hypoxanthine, methionine, and four phytoestrogens. Among phytoestrogens, plasma isoflavone metabolites (genistein, daidzein) were higher in cases than controls while lignan metabolites (matairesinol, pinoresinol) were lower (**Supplementary Fig. 3**). Adjustment for dietary intake (Model 2) had minimal impact, except for 16a-hydroxyestrone, which met thresholds for significance.

**Fig. 4.**
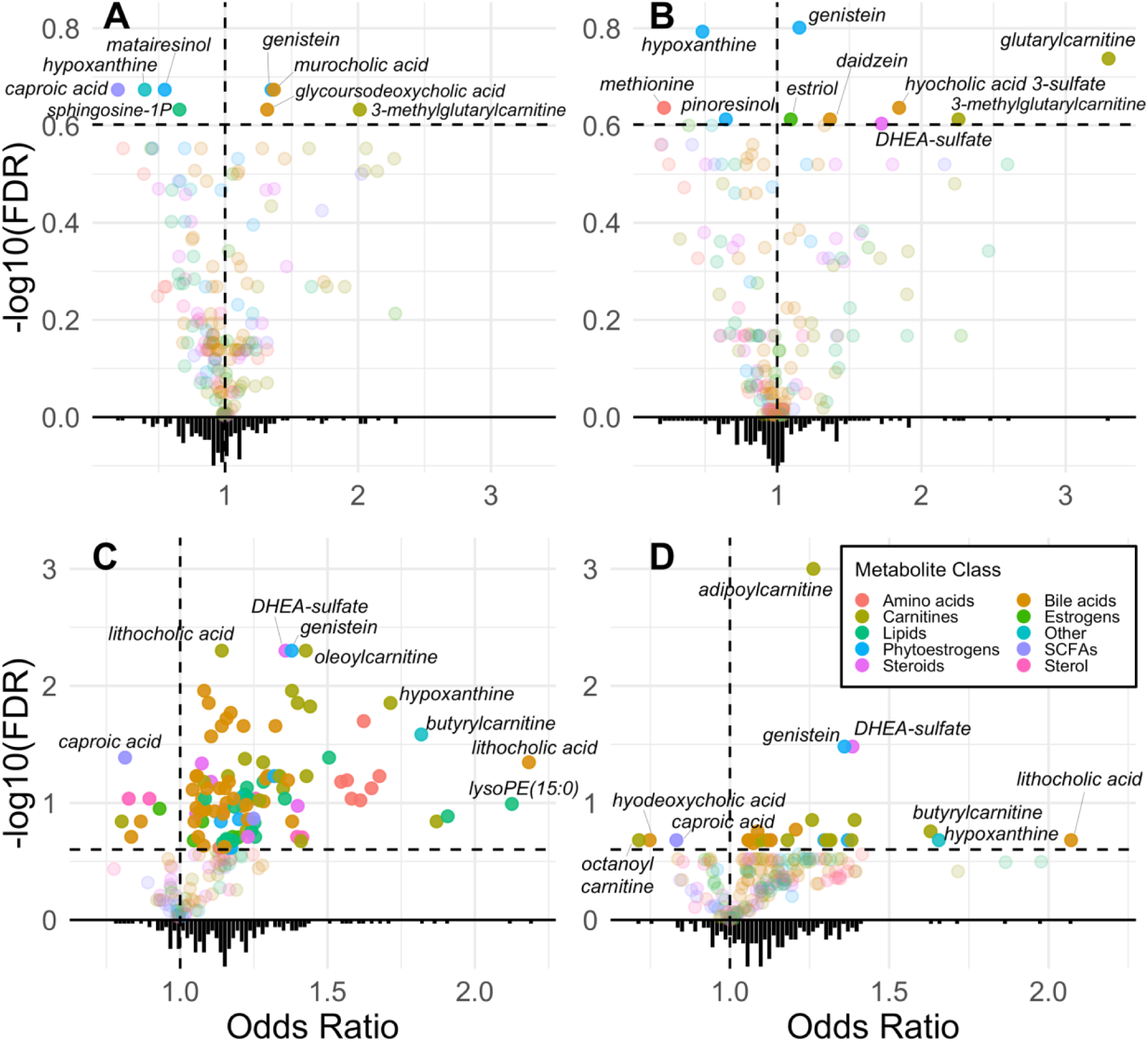
Volcano plot of odds ratios from logistic regression of plasma and stool metabolites. This included 182 plasma metabolites (A/B) and 185 stool metabolites (C/D). The left (A/C) are results from unadjusted models (Model 0), while the right (B/D) are from the confounder-set adjusted model (Model 1). Variables above the horizontal dotted line are significant at *q*<0.25.

Multivariate OPLS–DA showed moderate separation (R^2^Y=0.567) but limited ability to predict case/control status (Q^2^=0.22) (**Supplementary Fig. 4**). Permutation testing supported model validity (pR²Y=0.01, pQ²=0.01). Metabolites with the highest variable importance in projection (VIP) scores (**Supplementary Table 5**) largely overlapped with those identified by regression, including murocholic acid, genistein, daidzein, matairesinol, 16a-hydroxyestrone, hyocholic acid 3-sulfate, 3-methylglutaryl-carnitine, and taurolithocholic acid. Top features across models are summarized in **Fig. 5**, reporting ORs from Model 1.

**Fig. 5.**
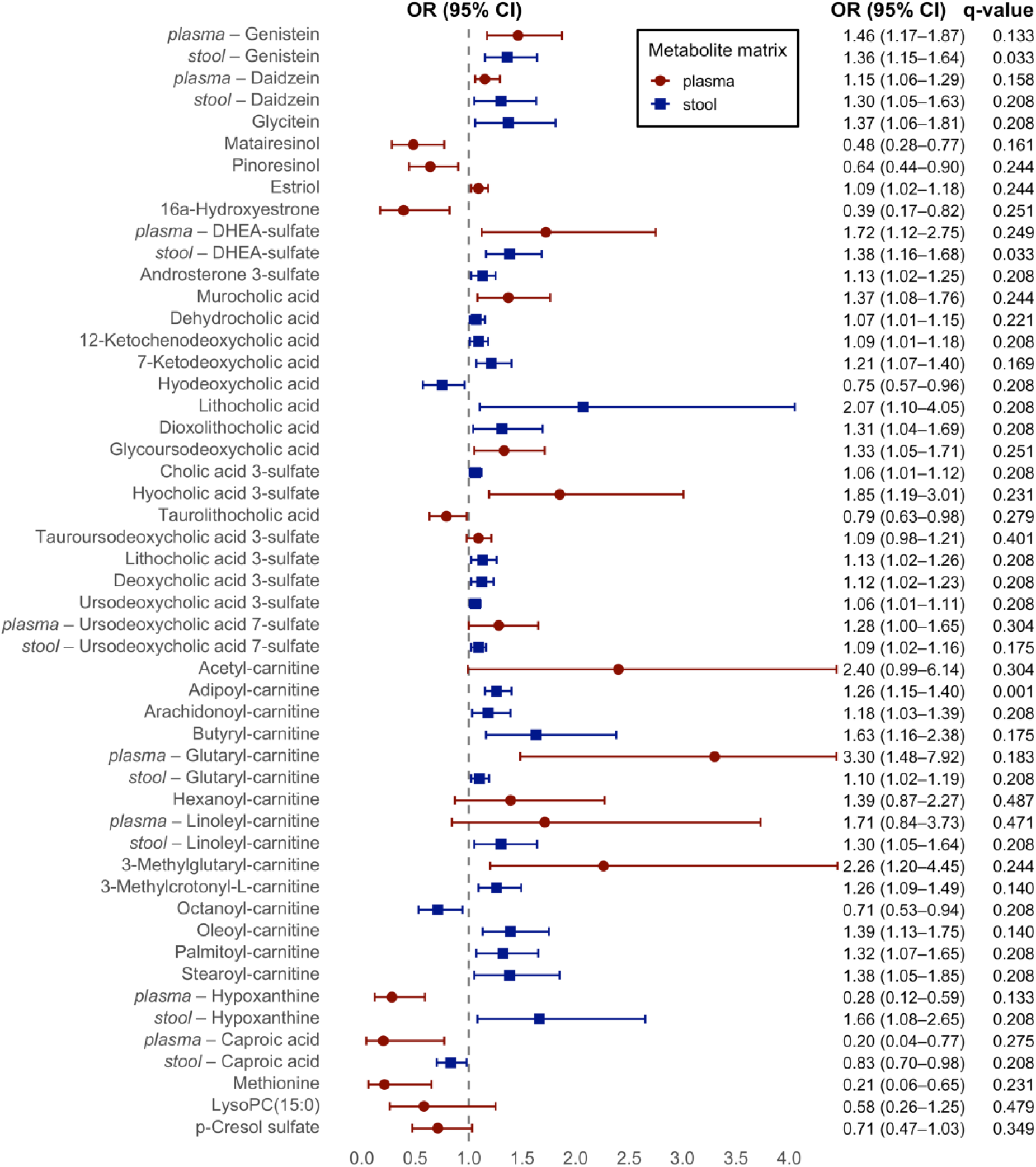
Metabolite differences between cases and controls shown by odds ratios from logistic regression. Top metabolites distinguishing cases and controls identified by univariate logistic regression and multivariate OPLS–DA. Odds ratios and q-values are reported from confounder-adjusted (Model 1) logistic regression. Upper confidence intervals for plasma acetyl- and glutaryl-carnitine were truncated for visualization.

### Individual stool metabolites differed by breast cancer status, but overall stool metabolite profiles did not

Although the overall stool metabolite profile (185 metabolites) did not yield a valid OPLS-DA model due to minimal variation between cases and controls, several individual metabolites showed significant differences by case–control status (**Supplementary Table 6**). Logistic regression identified 78 metabolites associated with breast cancer odds in unadjusted analyses (Model 0), and 28 in adjusted analyses (Model 1; **Fig. 4C–D**). This included 11 bile acids and derivatives (e.g., lithocholic acid, deoxycholic acid 3-sulfate, ursodeoxycholic acid 7-sulfate), 10 acylcarnitines (e.g., adipoyl-, oleoyl-, octanoyl-, palmitoyl-, stearoyl-carnitine), two steroid-hormone related (DHEA-sulfate, androsterone 3-sulfate), hypoxanthine, and three phytoestrogens (genistein, daidzein, glycitein). Among phytoestrogens, both stool isoflavone and lignan metabolites were higher in cases than controls (**Supplementary Fig. 3**). With adjustment for dietary intake (Model 2), glycoursodeoxycholic acid met thresholds for significance, while other associations were minimally impacted. Top features are shown in **Fig. 5**.

### Multi-omics reveals limited overall correlation across data types

Multiblock PLS-DA was applied using DIABLO to evaluate whether features identified in single-omics analyses jointly discriminated breast cancer case–control status (**Supplementary Table 7**). Model performance was modest (overall error rate = 0.308; BER = 0.311), corresponding to ∼70% correct classification of breast cancer status. Although clear separation between cases and controls was not observed, plasma metabolites and microbial species contributed most strongly to discrimination (**Supplementary Fig. 5**).

The DIABLO-derived correlation circle plot (**Fig. 6**) showed clear separation on the first component across omics layers, with stool and plasma metabolites loaded negatively and metagenomic features loading positively. While cross-omic clustering was limited, three groups of interest are noted. Box 1 highlights two methanogenic enzymes typically carried by archaea, dihydromethanophenazine:CoB–CoM heterodisulfide reductase (1.8.91.1) and formylmethanofuran dehydrogenase (1.2.7.12). Box 2 highlights two vitamin K_2_-related pathways (PWY-7992: menaquinol-8 biosynthesis III and PWY-7371: 1,4-dihydroxy-6-naphthoate biosynthesis II) and two TCA-related pathways (PWY-5130: 2-oxobutanoate and P42-PWY: incomplete reductive TCA cycle) connected by *Anaerotruncus rubiinfantis*. Box 3 highlights plasma methionine near methylglyoxal degradation (METHGLYUT-PWY).

**Fig. 6.**
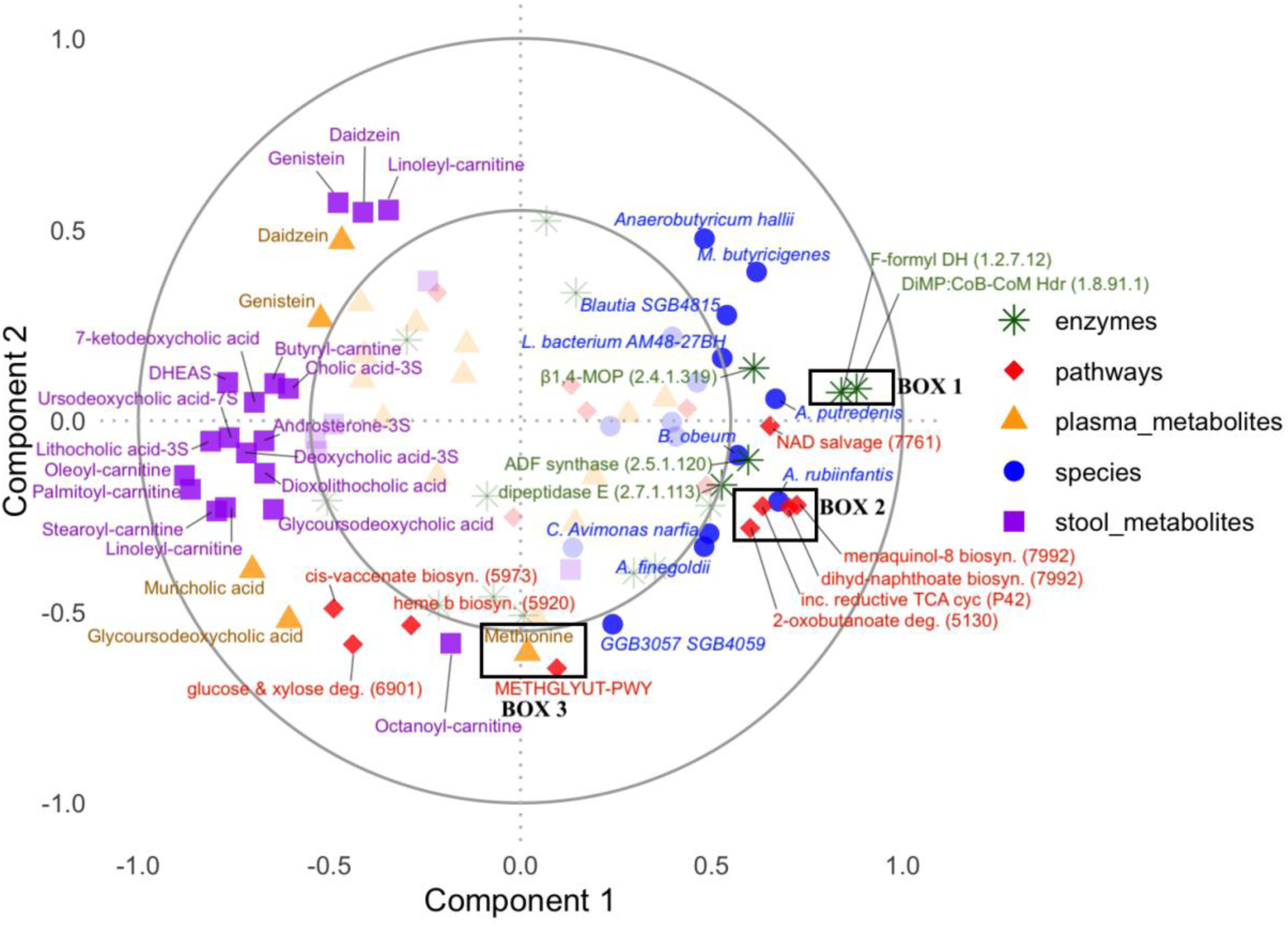
Correlation circle plot from DIABLO showing metagenomic and metabolomic feature contributions to components 1 and 2. DIABLO used multiblock PLS–DA modelling. Feature types are distinguished by symbol and colour. Features with scores greater than |0.5| in both components were bold and labelled; features below this threshold were made transparent.

## Discussion

In this case–control study of postmenopausal ER+ breast cancer, the association of reduced species-level alpha and beta diversity with breast cancer status was consistent with previous reports.^6,8,9,13,25^ However, associations with estrogen levels and estrogen-related microbial functions, such as the key estrobolome targets β-glucuronidase and arylsulfatase, were not observed. Although these deconjugation functions have been highlighted in experimental studies,^5^ their effects may be too modest or nonspecific relative to broader host and environmental factors to yield detectable differences in humans. Moreover, previous observational studies have not directly measured these functions due to sequencing method limitations.

We observed nine estrobolome taxa with modest, non-FDR-significant associations in logistic regression, most of which were depleted in cases, opposite the hypothesized direction for increased deconjugation functions; only *Clostridium butyricum, Eubacterium clostridioformis,* and *E. ramosum* were associated with higher breast cancer odds. Comparisons with previous studies were limited to the genus level, where *Ruminococcus*,^12,25,26^ *Lachnospira*,^8,10^ and *Bifidobacterium*^8,25^ were depleted, broadly consistent with prior reports, whereas *Clostridium* was enriched, contrasting an earlier study.^10^ Among XGBoost top discriminating features, the estrobolome genus-level target *Alistipes* was identified, along with three species (*A. putredinis, A. shahii, A. finegoldii*) that could represent novel species-level targets, although their estrogen-processing roles have not been validated beyond the genus level. These *Alistipes* features were all associated with lower odds of breast cancer, consistent with previous reports of depleted *Alistipes* in cases,^6,26,27^ but again opposite the expected direction for deconjugation functions.

*Alistipes* species have also been linked to protective immune effects in conditions such as inflammatory bowel disease.^28–30^ Overall, these findings highlight the variability of previously proposed estrobolome taxa in human populations and the need for further research to clarify which microbial species or enzymatic functions are most relevant for breast cancer risk. Future studies should also explore strain-level relationships using metagenome-assembled genomes to better characterize functional potential.

Case–control differences in steroid precursors and estrogen metabolites suggested broader influences on hormone exposure between cases and controls. DHEA-sulfate, and its downstream metabolite androsterone 3-sulfate, were associated with an increased odds of breast cancer, consistent with prior findings,^22,31,32^ though experimental studies also suggest potential protective effects through reduced breast cancer cell migration and invasion,^33^ leaving its role in cancer uncertain. Estriol showed an increased odds of breast cancer and has been linked to increased risk.^34,35^

Dietary phytoestrogens and their microbially produced metabolites represent another source of estrogen exposure, generally considered protective for breast cancer due to preferential ERβ binding.^24^ Isoflavones, which can suppress lymphocyte proliferation,^36^ enhance T-cell cytotoxicity,^36^ and reduce tumor growth,^37^ were higher in plasma and stool of cases, possibly reflecting differences in dietary sources of phytoestrogens, such as soy. Lignans were lower in plasma but higher in stool of cases, with significant differences in pinoresinol and matairesinol precursors, suggesting reduced microbial conversion of lignans into bioactive enterolignans, such as enterolactone which has been linked to lower breast cancer risk^23^ and exhibits immune-modulating and anti-cancer properties.^36,38^ Consistent with this, taxa implicated in phytoestrogen metabolism—*Adlercreutzia equolifaciens* (equol producer), and *Blautia obeum* and *Alistipes* spp. (β-glucosidase carriers)^353442404123,38,39^—were depleted in cases, also suggesting reduced microbial capacity to produce bioactive metabolites. Together, these findings indicate that both dietary intake and microbial metabolism of phytoestrogens may contribute to case–control differences, although more detailed dietary data are needed to confirm these relationships.

Overall, these findings suggest that, beyond circulating estrogens, steroid precursors, estrogen metabolites, and diet–microbiome interactions collectively shape the hormone environment and may be relevant for breast cancer risk. The estrobolome and other hormone-related microbial functions remain incompletely defined, particularly for microbial effects on androgen precursors, estrogen metabolites, and dietary phytoestrogens, and the current reference databases limit the ability to fully assess these microbiome–hormone interactions. Breast cancer is also a heterogeneous disease arising from complex exposures, and additional microbiome effects on disease etiology beyond estrogen- and hormone-related mechanisms may be relevant.

In our study, the top XGBoost-discriminating taxa were largely gut commensals involved in fiber fermentation and short-chain fatty acid metabolism, including *Anaerobutyricum, Blautia,* and *Lachnospiraceae* spp., which contribute to host–microbe metabolic and immune pathways rather than direct pathogenic or pro-inflammatory processes. Although overall functional diversity did not differ between cases and controls, several XGBoost-identified pathways and enzymes were highlighted, primarily reflecting core microbial functions such as carbohydrate and amino acid metabolism. Most of these functions had lower abundance in cases, suggesting reduced microbial functional capacity in cases. DIABLO-based multi-omics analysis showed limited overall correlation across data types, with modest separation along the first component and a small number of clustered features.

Menaquinol-8 (vitamin K_2_) biosynthesis pathways and two related enzymes—UDP-N-acetylglucosamine 4,6-dehydratase and aminodeoxyfutalosine synthase—were associated with lower odds of breast cancer, suggesting reduced microbial capacity to produce vitamin K_2_.

Supporting this, a Mendelian randomization study identified a related menaquinol-8 pathway (PWY-6263) linked to breast cancer risk.^41^ Vitamin K_2_, produced by microbes and found in fermented foods and animal products, may have antiproliferative and antimetastatic effects.^42,43^ These findings suggest that microbial vitamin K2 pathways warrant further investigation in breast cancer.

Incomplete reductive TCA cycle (P42-PWY) and 2-oxobutanoate degradation were associated with a lower odds of breast cancer. 2-Oxobutanoate feeds into the TCA cycle via succinyl-CoA, reflecting microbial energy and amino acids metabolism, including sulfur-containing amino acids such as methionine. DIABLO analysis clustered these features near *Anaerotruncus rubiinfantis* and vitamin K_2_ pathways. Plasma methionine, central to one-carbon metabolism, DNA methylation, and redox status,^44^ was also associated with lower odds of breast cancer and clustered near the top discriminating pathway methylglyoxal degradation (METHGLYUT-PWY), which catabolizes methionine into intermediates such as 2-oxobutanoate. Previous mendelian randomization analyses have observed links between methylglyoxal degradation and breast cancer.^41^ These findings point to reduced microbial capacity for energy metabolism, amino acid catabolism, and redox pathways in breast cancer cases.

Higher levels of conjugated bile acids were seen in cases, and given that bile acid deconjugation primarily occurs via bile salt hydrolases expressed by gut microbes,^45^ this may suggest reduced microbial capacity to process bile acids. Similarly, acylcarnitines, intermediates of fatty acid β-oxidation that reflect mitochondrial energy metabolism and carnitine-mediated transport of fatty acids into the TCA cycle,^46^ were generally higher in cases, suggesting increased lipid flux and altered energy metabolism. Individual associations were mixed relative to prior studies: octanoyl-carnitine,^46^ 3-methylglutaryl-carnitine,^47^ and oleoyl-carnitine^16^ aligned with some earlier reports, while oleoyl-carnitine,^48^ arachidonoyl- and linoleyl-carnitine^20,48^ contrasted with other earlier study results. Such inconsistencies, as with those seen in the microbiome findings, likely reflect differences across cohorts, including study populations (e.g., inclusion/exclusion criteria, menopausal status, cancer subtype), sequencing and metabolomics platforms, study design, confounding control, and analytic approaches.

Limitations of this study include the relatively small sample size, which may have limited power to detect small to moderate differences. There was modest imbalance in ethnicity and current hormone therapy use between cases and controls; however, the potential effect of these differences in unclear. Prior studies suggest that environment factors outweigh host genetics in shaping the gut microbiome.^49^ Lifestyle factors and dietary intake were balanced between cases and controls (Table 1), and exploratory analyses stratified by ethnicity revealed no differences.

Current hormone therapy use was captured both at screening (yes/no) and at enrollment (detailed medication list). Comparison of these two measures revealed inconsistencies: several participants who reported “yes” at screening did not list any hormone-related medications at enrollment, while others reported only localized formulations (e.g., topical creams or vaginal tablets) and only a few reported systemic estrogens or progesterone. As a result, it is unclear whether these self-reported differences reflect meaningful variation in estrogen exposure. Non-fasting blood samples were for metabolomics to align with pre-surgical clinical blood work which did not require fasting, and previous work suggests fasting minimally affects overall metabolite variability.^50^ Finally, the targeted metabolite panel measured estrogen metabolites and select phytoestrogens but did not capture broader microbial metabolites, limiting integration with metagenomic data.

The study has several strengths. All cases were treatment-naïve at sampling, reducing bias from cancer therapies and post-surgical antibiotics; while physiological effects of cancer development or stress on microbiome function cannot be ruled out, Mendelian randomization evidence does not support reverse causation.^41^ Careful consideration of inclusion and exclusion criteria, such as excluding participants with gastrointestinal conditions, reduced potential biases. Analyzing multiple data types—metagenomics with species-level resolution and functional annotation, plasma and stool metabolomics to compare circulating versus excreted metabolites, and dietary intake—enabled multi-level analysis and integration across data types.

Comprehensive adjustment for potential confounding factors further strengthened the validity of observed associations. Finally, the study extended our understanding of the estrobolome to include metabolism of androgen precursors, estrogen metabolites, and phytoestrogens, and identified other novel microbial/metabolic associations with breast cancer.

In conclusion, estrogens and their direct microbial functional targets did not show clear associations with breast cancer. Novel associations were identified for androgen precursors, estrogen metabolites, and phytoestrogens with breast cancer. Phytoestrogen metabolites and microbes known or hypothesized to metabolize phytoestrogens suggest reduced microbial capacity for phytoestrogen processing among cases. The estrobolome remains incompletely defined, highlighting the need for improved reference database annotations, as many microbial taxa and functional genes involved in hormone and phytoestrogen metabolism are poorly characterized, limiting their detection and interpretation in metagenomic studies. Beyond hormone-related functions, several microbial and metabolic features were associated with breast cancer, including conjugated bile acids, carnitines, *Anaerotruncus rubiinfantis*, *Blautia obeum, Alistipes* species, menaquinol-8 biosynthesis, TCA cycle-related energy pathways, and NAD salvage—many suggesting reduced microbial functional capacity in cases. Larger multi-omic studies with functional profiling and dietary integration are needed to validate results and refine targets linking the gut microbiome to breast cancer.

## Methods

### Study design and participants

Female breast cancer cases (n=70) were recruited, from 2022-2024, from four breast cancer surgical clinics in Vancouver and Victoria, British Columbia, Canada. Female controls (n=70) were recruited during the time period using multiple strategies, including infographics posted at community sites and mammography clinics, outreach to friends of cases, and REACH BC—a provincial platform connecting volunteers to research studies.^51^

To be included in the study, cases had to be postmenopausal biological females residing in British Columbia, Canada and have a hormone receptor-positive breast cancer diagnosis (stage I-III) with no prior surgical or systemic therapy for the current diagnosis, as confirmed by electronic medical records and pathology reports. Potential participants were excluded if they had a previous cancer diagnosis (except non-melanoma skin cancer), body mass index >35 kg/m^2^, functional gastrointestinal disorders, prior gastric banding or by-pass surgery, a history of other gastric or intestinal surgery within the previous six months, and antibiotic use within the 30 days prior to enrollment. A flowchart of study inclusion is shown in **Supplementary Fig. 7**. Ethics approval was obtained from the University of British Columbia–BC Cancer Research Ethics Board (REB: H21-02319).

### Data collection

Upon enrollment, participants completed online, self-administered questionnaires covering health history (medical conditions, current medications, family history of cancer), sociodemographic and lifestyle factors (age, BMI, ethnicity, education, smoking, reproductive history), physical activity (International Physical Activity Questionnaire [IPAQ]^52^), and dietary intake over the past 12 months (Canadian Diet History Questionnaire [CDHQ], version III, developed by the National Cancer Institute, MD, US^53^).

Participants were asked to collect the first void stool of the day. Stool samples were then taken to their blood draw appointment. For breast cancer cases, the blood draw, collected in 2x4mL sodium heparin tubes, could be completed alongside routine pre-surgical blood work to reduce participant burden. Samples were shipped to the BC Centre for Disease Control lab (Vancouver, CA) where whole stool and blood plasma were stored in −80°C freezers until processing.

### DNA extractions, library preparation and sequencing

Total microbial DNA was extracted from 200 mg of feces using the DNeasy PowerSoil Pro Kit (Qiagen, DE) following the manufacturer’s instructions. DNA quantity was assessed by fluorometry (Qubit, ThermoFisher, US), and quality was confirmed by spectrophotometry (SimpliNano^TM^, Fisher Scientific, AU). Extracted DNA was sheared to an average fragment size of 350–400 bp using the Bioruptor Pico (Diagenode, BE). For metagenomic library preparation, 1 μg of extracted DNA was used as input. Libraries were constructed using 125 ng of DNA with the NEBNext Ultra II DNA Library Prep Kit for Illumina (New England Biolabs, US). The concentration and fragment size of the libraries were assessed by qPCR and TapeStation (Agilent, US). DNA-free negative controls were included in all DNA extraction and library preparation steps. Libraries were randomized and then pooled into batches of 24 samples, each including one negative control (n=13). A subset of specimens underwent replicate DNA extraction, library preparation, and sequencing to estimate technical variability and batch effects among samples. Whole metagenomic sequencing was performed on the Illumina NovaSeq platform (BC Genome Sciences Centre, Vancouver, CA), with 24 libraries per lane, generating approximately 16 million paired-end reads per sample.

### Bioinformatics

The high-performance computing cluster (Cedar) maintained by the Digital Research Alliance of Canada was used for all bioinformatic analyses. Sequence reads were trimmed of adapters and filtered to remove low-quality, short (<60 base-pairs), and duplicate reads as well as those of human, other animal or plant origin, using *KneadData*.^54^ Species composition was determined by identifying clade-specific markers from reads using MetaPhlAn4 with default settings.^55,56^ Relative abundances of species were estimated from known assigned reads, with proportions of unclassified reads inferred based on total, assigned, and unassigned reads. A minimum abundance threshold of > 3^rd^ percentile (0.0008) and ≥10% prevalence was applied to all taxonomic features in all downstream analysis.^56^ MetaPhlAn4 also assigns a subspecies-level Species Genome Bin (SGB) identifier to each species, with some species represented by multiple SGBs; these were retained to maximize resolution, though SGBs are not currently linked to NCBI taxonomy codes.

Functional gene and metabolic pathway composition was determined using HUMAnN3 with default settings against the UniRef90 database.^56,57^ Functionally annotated reads were further classified into level four enzyme commission (EC) categories.^58^ Enzyme and pathway abundance estimates were normalized using reads per kilobase per million mapped reads (RPKM) and then re-normalized to relative abundance. A minimum relative abundance threshold of ≥3^rd^ percentile (pathway features >8.37e-07; EC features >1.30e-07) and ≥5% prevalence were applied to all functional microbiome features in all downstream analysis.^56^

Metagenomic data were centered log-ratio transformed for linear-based modeling using a pseudo-count of half the smallest non-zero value (taxa=3.0e-5; pathways 2.7e-7; ECs=4.1e-8). Estrobolome targets were selected primarily from *in vitro* and animal model findings summarized in the literature,^5,24,59,60^ encompassing estrogens, their precursors and metabolites, and phytoestrogens, as well as microbial features involved in their processing. This included 16 genera, 71 species, 17 enzymes, and six pathway targets (**Supplementary Table 2)**.

### Metabolomics

Targeted metabolomics of plasma and stool samples were run using Agilent 1290 series Ultra-High Performance Liquid Chromatography coupled with Bruker Impact II high resolution quadrupole time-of-flight Mass Spectrometry (UHPLC-MS; Proteomics Centre, Victoria, BC).

A targeted panel of metabolites was run across plasma (n=230) and stool (n=237) including steroid hormones, bile acids, amino acids, lipids, SCFAs and carnitines. The full list is provided in **Supplementary Table 8**. Dry mass normalized concentrations were used to reduce variability due to water content and better reflect actual metabolite concentrations in the biological matrix and for comparing concentration differences between samples. Metabolites with no detectable level in ≥ 90% of samples were excluded in subsequent analyses,^61^ resulting in a total of 182 metabolites in plasma and 185 in stool for analysis.

Metabolite data were log_2_-transformed using a pseudo-count of half the smallest non-zero value (plasma=8.24e^-06^; stool=4.67e^-05^). Extreme outliers (>3 SD from the mean based on z-score scaling) were removed from individual metabolite models. For metabolites measurements, two participants were missing plasma (n=138), and one was missing stool (n=139).

### Participant characteristics

All analyses were conducted in R (v4.5.1). Dietary variables were normalized using the nutrient density method to g/1000 kcal and SD-scaled.^181,182^ The sample was described using frequency counts with percentages for categorical variables and median with interquartile ranges for continuous variables. Characteristics were compared across cancer status using chi-square tests (or Fisher’s for non-parametric data) and Wilcoxon rank-sum tests. Four participants were missing general health and/or dietary survey data and were excluded from adjusted models due to missing covariate information.

### Metagenomics analyses

Alpha diversity metrics, including Shannon index and species richness, were calculated using the *vegan* package (v2.6-10), and group differences were assessed with Wilcoxon tests. Beta-diversity was estimated using Bray–Curtis dissimilarities and visualized with principal coordinate analysis (PCoA) to explore clustering by cancer case-control status; group differences were assessed using permutational multivariate analysis of variance (PERMANOVA).

Multivariable logistic regression was used to investigate the independent associations between estrobolome features and the odds of breast cancer. Model 0 was unadjusted. Model 1 included a minimally sufficient adjustment set of potential confounders, selected based on domain knowledge and prior literature.^8,18,47^ Covariates included age (years), BMI (kg/m^2^), ethnicity (White, non-White), education level (≤ Bachelor’s, Bachelor’s, Postgraduate), current use of hormone replacement therapy for menopause (yes/no), smoking status (ever/never), physical activity (IPAQ categories: low, medium, high), and alcohol intake (g/day). Building on Model 1 covariates, Model 2 additionally adjusted for dietary intake—fiber, saturated fat, and total sugar—to approximate diet quality, with values normalized per 1000 kcal using the nutrient density method.^181,182^ Associations were reported as odds ratios with 95% confidence intervals (CIs). P-values were adjusted for false-discovery rate (FDR, or *q*-value) using the Benjamini-Hochberg procedure,^62^ with *q*<0.25 applied for statistical significance.

Extreme gradient boosting (XGBoost) was used to identify compositional and functional microbiome features that may contribute to breast cancer status in an untargeted analysis. XGBoost has been previously applied in microbiome research and offers variable importance outputs that support feature discovery in exploratory contexts.^56,58^ XGBoost models were developed separately for each microbiome feature set (genera, species, pathways, or enzymes) using the H20.ai engine and *h2o* R package interface with the *xgboost* package. Each XGBoost model was fit using a three-stage Bayesian optimization booster method as described previously.^58^ In stage one, the *BayesianOptimization* function with 5-fold cross-validation was used to select hyperparameters by minimizing the mean squared error (MSE). Models in the lowest 5^th^ percentile of MSE were retained, and from these models the features contributing to the top 95% of variable importance by proportion were retained. In stage two, these retained features were used to build new models, tuned as in stage one but with leave-one-out cross-validation (LOOCV), and features were again selected based on variable importance. In stage three, retained features and optimized hyperparameters from stage two were used to fit the final models, except for the parameter *ntrees* was increased while holding *ntrees × learning_rate* constant to obtain ∼20 minutes of runtime, and run using LOOCV. Final model performance was evaluated using pseudo-R^2^ and mean absolute error, with pseudo-R^2^ < 0 indicating performance worse than the mean response. This three-stage method was used to build models for each microbiome feature set, both without epidemiologic covariates and with epidemiologic covariates included.

### Metabolomics analyses

Unadjusted concentrations of estrobolome-related metabolites were described as mean values for case and control groups. Associations between individual metabolites and the odds of breast cancer were examined using multivariable logistic regression, consistent with the metagenomics analyses. Overall metabolite profiles were examined using supervised multivariate modelling with Orthogonal Partial Least Squares–Discriminant Analysis (OPLS–DA). Metabolites were ranked by their Variable Importance in Projection (VIP) scores, with higher scores indicating features that contribute most to the predictive component while accounting for correlations among all other features. OPLS–DA models were run separately for plasma and stool metabolites. Model performance was assessed by separation (R^2^Y), predictive ability (Q^2^), and statistical validity (pR^2^Y, pQ_2_).

### Multi-omics integration

Integration of metagenomic and metabolomic data was performed using DIABLO, a supervised multiblock PLS-DA method (*mixOmics*; v6.3.2). Features identified as significant in prior single-omics analyses for each data type were included in the DIABLO model.^63^ Cross-block correlations between selected features were visualized using circle plots, and model performance was assessed using 10-fold cross-validation repeated 10 times. This approach highlights co-varying features across omics layers that jointly contribute to group separation and may reflect underlying biological pathways.

### Plasma quantification of anti-LPS and anti-Flagellin IgGs by ELISA

Plasma IgG levels against lipopolysaccharide and flagellin were quantified by ELISA as potential markers of systemic bacterial exposure in this cohort (Supplementary Methods; Supplementary Fig. 6).

## Declarations

### Data Availability

Data generated or analyzed during this study are not publicly available due to privacy considerations and risk of subject identification given the small group of participants. Sharing of data with investigators may be possible upon access request to the corresponding author and approval by the institutional research ethics committee at the University of British Columbia.

## Author Contributions

R.A.M., A.R.M., J.J.L., J.S., and P.B. wrote the initial grant that funded this work. R.A.M. and A.R.M. supervised the study, while A.L. coordinated participant recruitment and data collection and managed the research study. K.I., R.W., M.G., A.H., and A.R. assisted with recruitment of breast cancer cases and coordination with their clinics. J.H. and D.R.G. processed blood and stool samples for metabolomics, and B.C. and M.D. processed plasma IgG biomarkers. T.E. performed metagenomic data assembly, bioinformatics analyses, and machine learning modeling. A.L. conducted data analysis and drafted the manuscript. All authors reviewed and edited the manuscript.

## Acknowledgements

The authors sincerely thank all study participants for generously giving their time and providing samples that made this research possible. The authors also thank Wilson Chan for assistance with stool sample library preparation, and Truman Wood and Ezra Yu for support with participant recruitment. We also acknowledge the research assistants and students at the breast clinics who contributed to patient screening and chart reviews, as well as the physicians who aided recruitment, including those at Providence Breast Centre, Fraser Health, and Island Health. We are grateful to Jessica Morgan and Sam Punch for their exploratory work on breast cancer cell lines and metabolites, which is not included in the present manuscript.

