## Supplementary Information for "Gut microbiome and metabolome reveal hormone-related and functional alterations in ER-positive breast cancer: a case–control study"

*co-senior authors

### Supplementary Methods

#### Plasma quantification of anti-LPS and anti-Flagellin IgGs by ELISA

Bacterial translocation across a compromised gut barrier has been implicated in carcinogenesis, with prospective studies linking elevated antibodies against lipopolysaccharide (LPS) and flagellin to increased risk of hepatocellular carcinoma[1] and colorectal cancer.[2,3] These findings suggest that chronic immune activation from gut-derived bacterial products may contribute to tumor development. We therefore examined circulating IgG levels against these bacterial components as potential markers of systemic exposure in this breast cancer cohort.

Quantification of specific anti-Lipopolysaccharides (LPS) and anti-flagellin IgG was performed by coating 96-well microtiter plates (Nunc™ MaxiSorp™ flat-bottom 96-well plates; Thermo Fisher Scientific) with 100 ng/well of laboratory-made Salmonella typhimurium-derived flagellin, or 2 µg/well lipopolysaccharides (from Escherichia coli 0128:B12, Sigma) in bicarbonate buffer (pH 9.6, Sigma) overnight at 4°C. After a wash, plasma samples were applied at a dilution of 1:200 in wash buffer (0.005% BSA, 0.01% Tween 20 in PBS) for 1h at 37°C. After incubation and washing, wells were incubated for 1h at 37°C with sheep HRP-linked anti-human IgG 1:1000 in wash buffer (GE Healthcare, NA933). Detection was carried out using 3,3′,5,5′-Tetramethylbenzidine substrate, and reaction was stopped with 2 N $H_2SO_4$. Optical density was measured at 450 nm with background correction at 540 nm using a microplate reader. Anti-LPS and anti-flagellin IgG levels were visualized using boxplots and compared between groups using p-values from logistic regression on log-transformed, z-scored data.

Anti-flagellin and anti-LPS IgG levels did not differ between cases and controls. Median anti-flagellin IgG levels were 206 (IQR 114) in cases versus 215 (IQR 164) in controls. Median anti-LPS IgG levels were 50.8 (IQR 33.2) in cases versus 46.3 (IQR 35.9) in controls.

#### Supplementary Figures

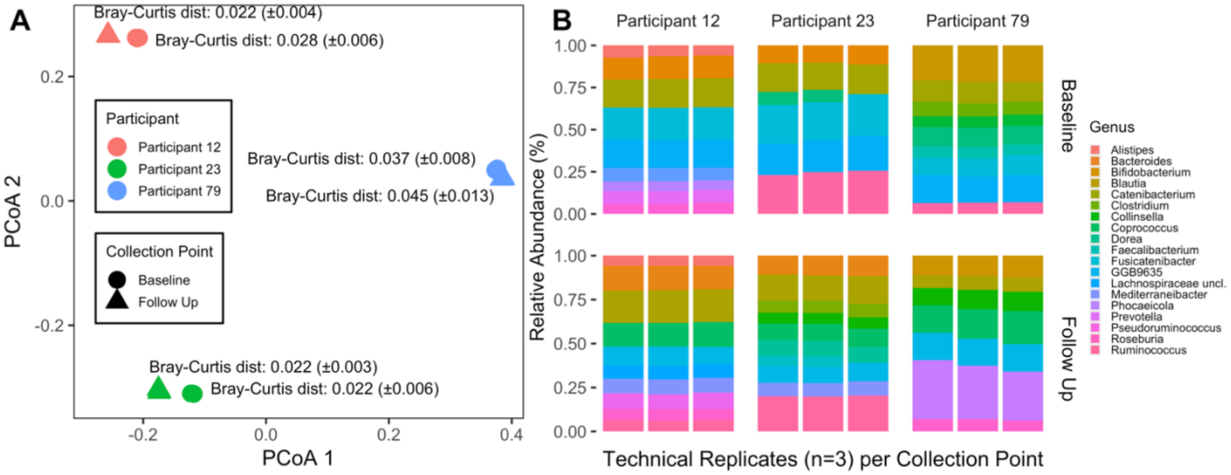


**Supplementary Figure 1.** Whole metagenome sequencing performance among 18 replicates. A) Bray-Curtis distances (± standard deviation) among replicate groups visualized using principal coordinate analysis (PCoA), and B) genus-level relative abundances showed high reproducibility and minimal technical variation.

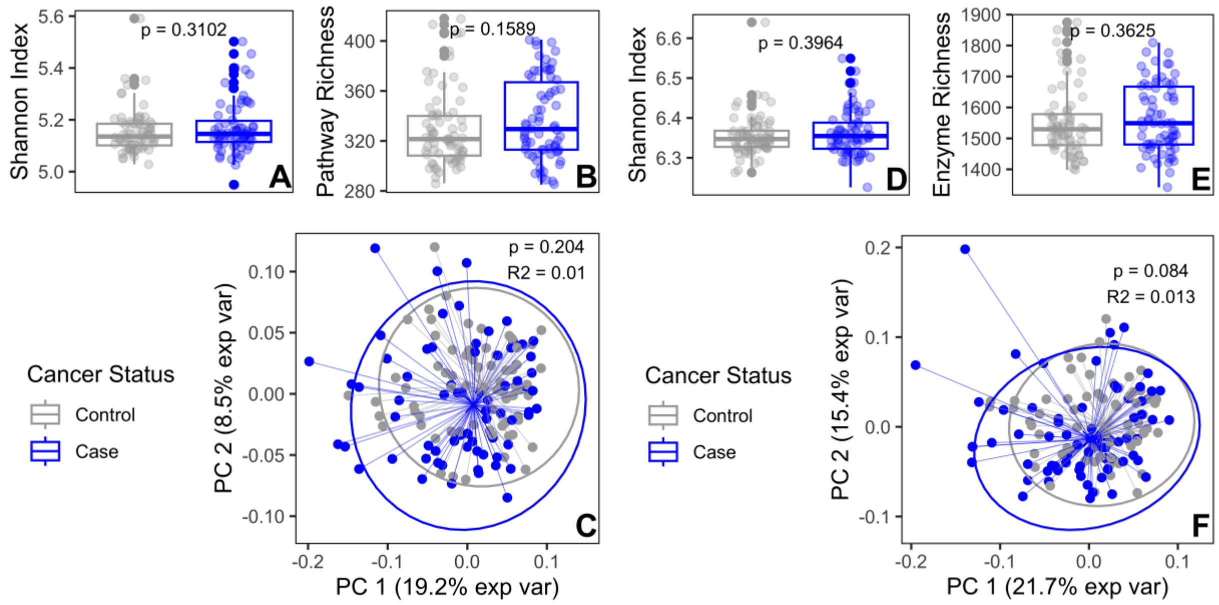


**Supplementary Figure 2.** Diversity measures of functional pathways (A-C) and enzymes (D-F). Alpha diversity by (A/D) Shannon index and (B/E) species richness, and (C/F) beta diversity using Bray-Curtis dissimilarity visualized by principal coordinate analysis (PCoA), showed no significant differences between cases and controls for both functional pathways and enzymes.

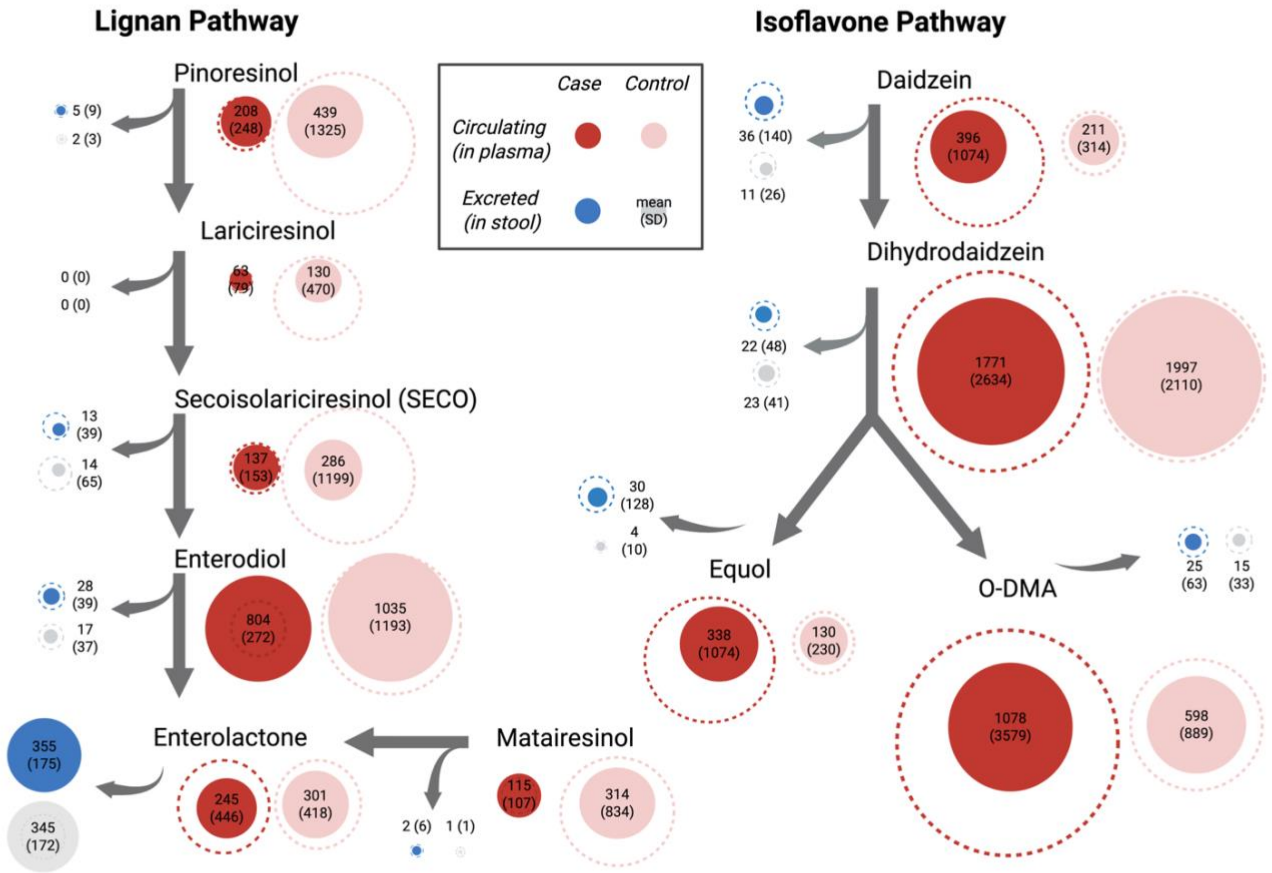


**Supplementary Figure 3.** Concentration differences in phytoestrogen pathways, including lignans and isoflavones. Lignans include the pinoresinol and matairesinol to enterolactone pathways. Isoflavones include daidzein to equol pathway. Plasma (red) and stool (blue) metabolites are shown as mean (SD) for cases (darker) and controls (lighter). Filled circles indicate group means, dotted circle indicate SD. Isoflavones were generally higher in cases in circulating and excreted levels, while lignans were lower in circulating but slightly higher in excreted levels.

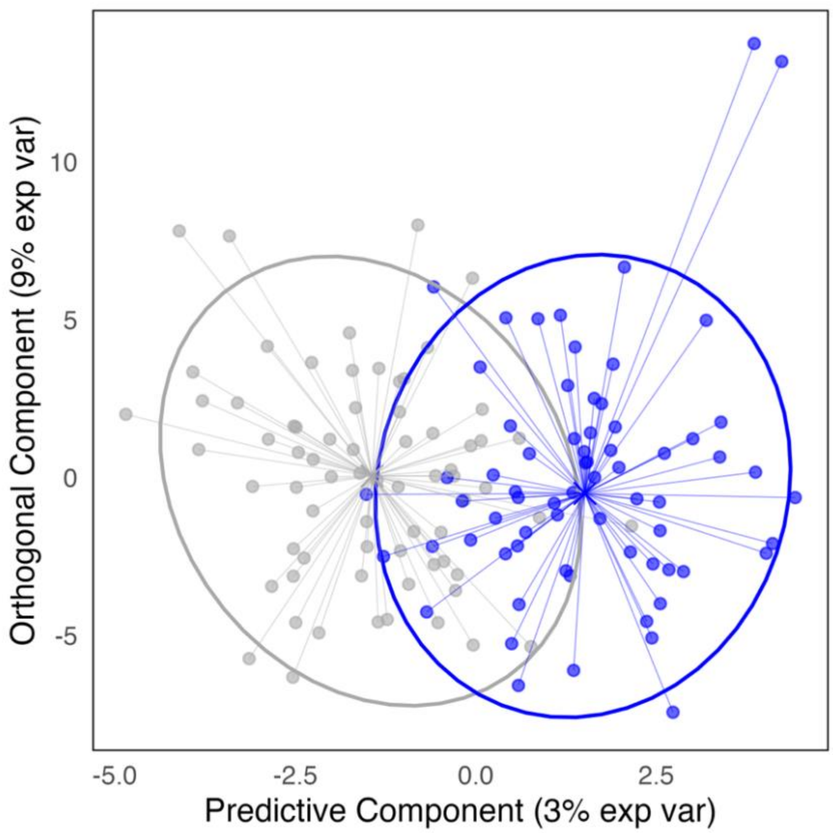


**Supplementary Figure 4.** OPLS-DA plot of plasma metabolite scores for predictive and orthogonal components. Separation is shown between case (blue) and controls (grey).

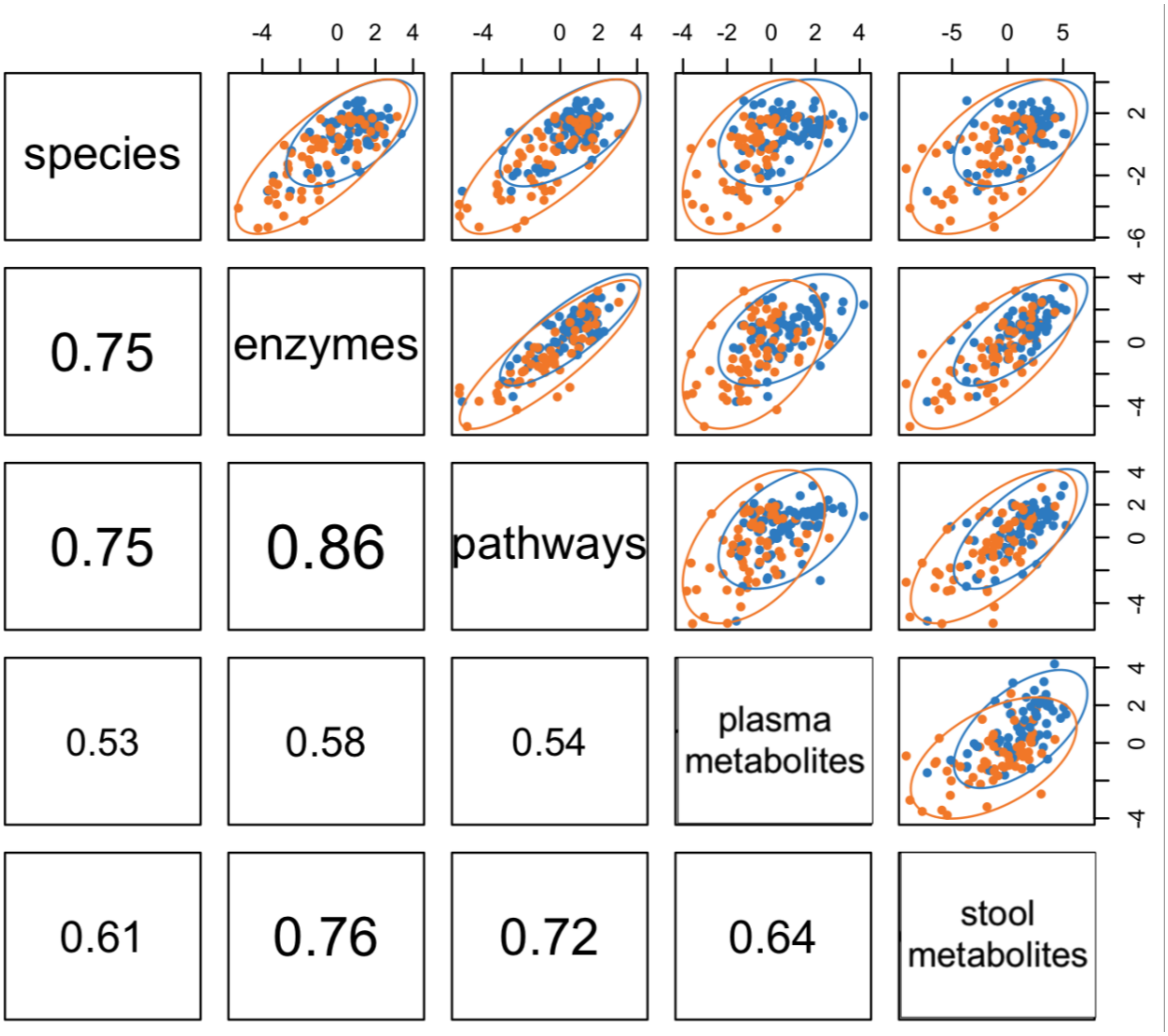


**Supplementary Figure 5.** Cross-block integration of metagenomic and metabolomics using DIABLO via multiblock PLS–DA modelling. Scatterplots show correlations between first components for each data type and separation of cases (orange) and controls (blue), with 95% confidence ellipses.

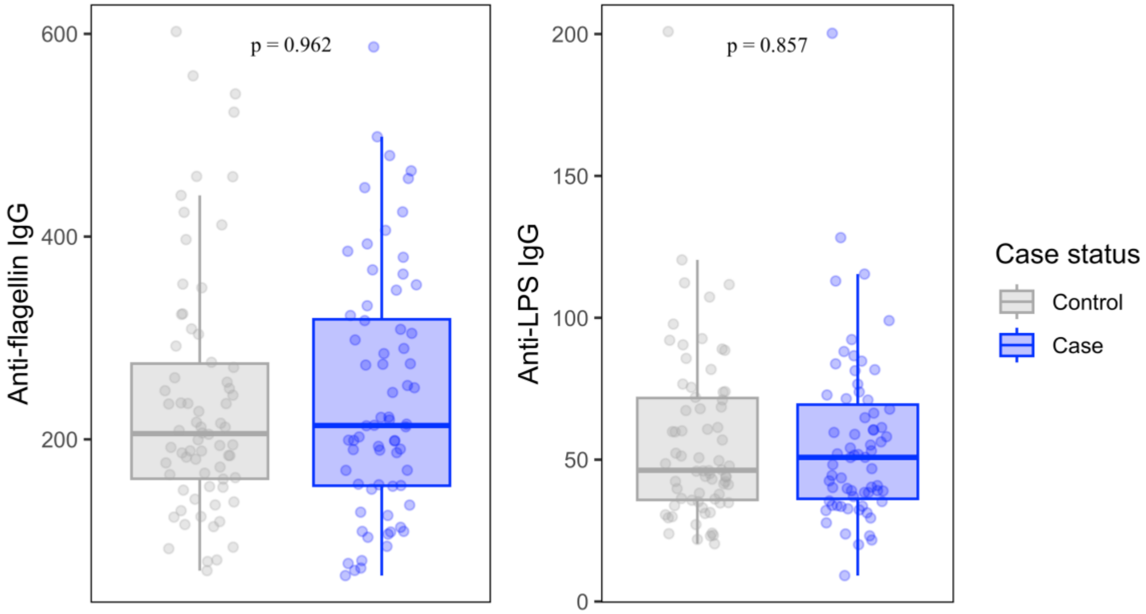


**Supplementary Figure 6.** Plasma anti-flagellin and anti-LPS IgG levels in cases and controls.

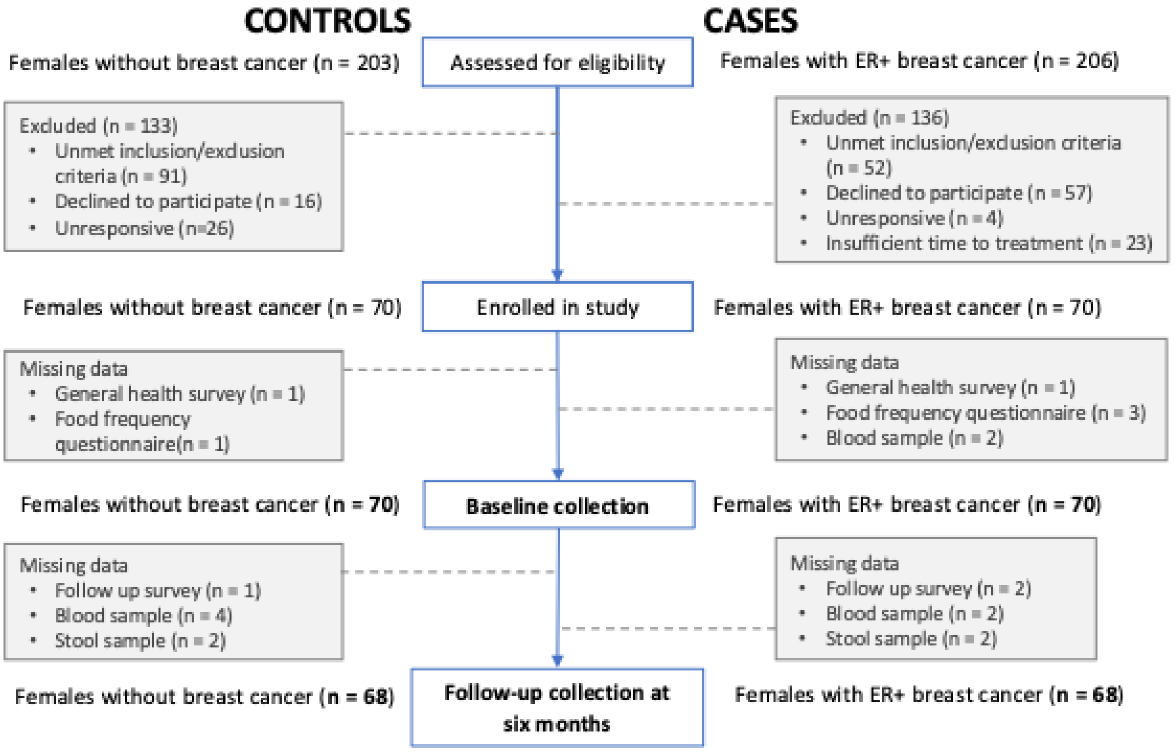


**Supplementary Figure 7.** Flow chart of recruitment of postmenopausal females to the case-control study and data collection at each time point. Time points include baseline, and follow-up approximately 6 months after baseline. Dotted lines describe excluded participants or missing data. Final data counts at each collection point are noted. The current study only included baseline data and analyses.

### Supplementary Tables

**Supplementary Table 1.** Past year dietary intake from food frequency questionnaires (CDHQ-III).

| Nutrient (/1000 kcal) | Controls (*n* = 69) mean (SD) | Cases (*n* = 67) mean (SD) | *p OR* | *p aOR* |
|---|---|---|---|---|
| **Macros** | | | | |
| Carbohydrates, g | 117.5 (19.6) | 120.5 (25.2) | 0.437 | 0.844 |
| Total dietary fibre, g | 14.28 (4.23) | 14.11 (4.72) | 0.827 | 0.734 |
| Total sugar, g | 48.49 (12.25) | 49.81 (22.43) | 0.669 | 0.985 |
| Protein, g | 41.57 (8.09) | 40.54 (9.04) | 0.485 | 0.603 |
| Fat, g | 39.27 (6.62) | 38.51 (8.16) | 0.549 | 0.892 |
| Total saturated fatty acids, g | 12.25 (2.99) | 11.66 (3.57) | 0.297 | 0.865 |
| Total monounsaturated fatty acids, g | 14.89 (3.23) | 15.25 (4.42) | 0.580 | 0.447 |
| Total polyunsaturated fatty acids, g | 8.73 (2.16) | 8.33 (2.19) | 0.286 | 0.569 |
| Omega 3 fatty acids | | | | |
| Alpha linolenic acid (ALA), g | 1.07 (0.61) | 0.97 (0.34) | 0.241 | 0.406 |
| Eicosapentaenoic acid (EPA), g | 0.05 (0.06) | 0.08 (0.07) | **0.052** | ***0.031*** |
| Docosapentaenoic acid (DPA), g | 0.02 (0.01) | 0.02 (0.02) | 0.252 | 0.138 |
| Docosahexaenoic acid (DHA), g | 0.10 (0.09) | 0.13 (0.11) | 0.081 | **0.051** |
| Omega 6 fatty acids | | | | |
| Arachidonic acid, g | 7.33 (1.81) | 6.99 (2.01) | 0.299 | 0.557 |
| Linoleic acid, g | 0.08 (0.05) | 0.08 (0.04) | 0.775 | 0.843 |
| **Amino Acids** | | | | |
| Leucine, g | 2.99 (0.67) | 2.98 (0.74) | 0.936 | 0.938 |
| Isoleucine, g | 1.75 (0.42) | 1.74 (0.45) | 0.865 | 0.957 |
| Valine, g | 2.01 (0.45) | 2.01 (0.49) | 0.967 | 0.904 |
| Methionine, g | 0.84 (0.25) | 0.84 (0.25) | 0.951 | 0.888 |
| Tryptophan, g | 0.47 (0.10) | 0.47 (0.11) | 0.743 | 0.931 |
| Arginine, g | 2.28 (0.49) | 2.25 (0.59) | 0.793 | 0.814 |
| Glutamic acid, g | 7.91 (1.43) | 8.01 (2.55) | 0.779 | 0.621 |
| Glycine, g | 1.63 (0.40) | 1.62 (0.49) | 0.875 | 0.785 |
| Histidine, g | 1.07 (0.25) | 1.06 (0.28) | 0.919 | 0.956 |
| Phenylalanine, g | 1.77 (0.34) | 1.75 (0.39) | 0.783 | 0.999 |
| Proline, g | 2.41 (0.44) | 2.38 (0.56) | 0.720 | 0.846 |
| **Vitamins** | | | | |
| Niacin, NE | 20.74 (3.54) | 20.59 (4.92) | 0.840 | 0.912 |
| Dietary folate equivalents, mcg | 266.4 (68.8) | 268.3 (76.3) | 0.874 | 0.519 |
| Vitamin B12, mcg | 2.04 (0.89) | 2.28 (1.18) | 0.189 | 0.095 |
| Vitamin C, mg | 87.0 (36.2) | 91.7 (52.7) | 0.549 | 0.366 |
| Vitamin D, mcg | 2.57 (1.50) | 2.96 (1.51) | 0.142 | 0.109 |
| Vitamin E, mg | 3.98 (2.41) | 4.52 (2.80) | 0.230 | 0.178 |
| Vitamin K, mcg | 212.1 (207.7) | 258.7 (285.7) | 0.281 | 0.374 |
| **Minerals** | | | | |
| Calcium, mg | 541.2 (175.4) | 553.7 (168.7) | 0.670 | 0.180 |
| Iron, mg | 7.64 (1.82) | 7.69 (2.08) | 0.885 | 0.614 |
| Magnesium, mg | 228.3 (52.2) | 242.2 (64.8) | 0.173 | 0.083 |
| Potassium, mg | 1931.2 (385.7) | 1990.7 (539.5) | 0.457 | 0.359 |
| Selenium, mcg | 55.32 (10.55) | 56.2 (15.4) | 0.690 | 0.918 |
| Sodium, mg | 1274.5 (249.1) | 1261.1 (314.9) | 0.781 | 0.991 |
| Zinc, mg | 5.62 (0.81) | 5.64 (1.09) | 0.882 | 0.394 |
| **Phytoestrogen-Related Foods** | | | | |
| Flaxseed, g | 0.69 (2.19) | 0.39 (0.88) | 0.340 | 0.367 |
| Total soy, g | 4.23 (15.74) | 5.19 (14.00) | 0.708 | 0.737 |

*Abbreviations: g, grams; mcg, micrograms; mg, milligrams; NE, niacin equivalents; OR, odds ratio; aOR, adjusted odds ratio; SD, standard deviation. Dietary variables were adjusted according to the nutrient density method to get grams per 1000 kcal and standardized by the population SD. Odds ratios were run for unadjusted models (Model 0) and adjusted (Model 1) and p-values were reported.*

**Supplementary Table 1**. Logistic Regression modelling results for the metagenomic estrobolome targets.

| Metagenomic Estrobolome Target | | Role | Model 0 (Unadjusted) N=140 | | | Model 1 (Confounder set) N=136 | | | Model 2 (Diet quality) N=136 | | |
|---|---|---|---|---|---|---|---|---|---|---|---|
| | | | OR (95% CI) | pval | qval | OR (95% CI) | pval | qval | OR (95% CI) | pval | qval |
| **Genus** | **NCBI ID** | | | | | | | | | | |
| Ruminococcus | 1263 | β-glucuronidase activity | 0.89 (0.79–0.99) | ***0.048*** | 0.192 | 0.87 (0.75–0.99) | ***0.045*** | 0.355 | 0.86 (0.74–0.99) | ***0.040*** | 0.314 |
| Alistipes | 239759 | β-glucuronidase activity | 0.77 (0.56–0.94) | ***0.043*** | 0.189 | 0.84 (0.64–1.02) | 0.117 | 0.483 | 0.84 (0.64–1.02) | 0.119 | 0.448 |
| Collinsella | 102106 | β-glucuronidase activity | 0.83 (0.67–0.98) | 0.051 | 0.192 | 0.85 (0.67–1.04) | 0.160 | 0.517 | 0.85 (0.66–1.04) | 0.147 | 0.481 |
| Faecalibacterium | 216851 | β-glucuronidase activity | 0.89 (0.74–1.01) | 0.106 | 0.282 | 0.93 (0.77–1.09) | 0.400 | 0.731 | 0.93 (0.76–1.08) | 0.366 | 0.696 |
| Escherichia | 561 | β-glucuronidase activity | 1.13 (1.03–1.24) | ***0.009*** | 0.061 | 1.05 (0.94–1.17) | 0.401 | 0.731 | 1.05 (0.94–1.17) | 0.387 | 0.704 |
| Roseburia | 841 | β-glucuronidase activity | 0.90 (0.75–1.06) | 0.225 | 0.38 | 0.93 (0.74–1.12) | 0.484 | 0.731 | 0.93 (0.74–1.12) | 0.475 | 0.717 |
| Lactobacillus | 1578 | β-glucuronidase activity | 1.02 (0.91–1.16) | 0.720 | 0.825 | 1.04 (0.90–1.19) | 0.630 | 0.804 | 1.04 (0.90–1.20) | 0.593 | 0.752 |
| Bacteroides | 816 | β-glucuronidase activity | 1.22 (0.81–1.85) | 0.344 | 0.509 | 1.12 (0.69–1.82) | 0.657 | 0.808 | 1.13 (0.69–1.86) | 0.634 | 0.787 |
| Bifidobacterium | 1678 | β-glucuronidase activity | 1.05 (0.98–1.14) | 0.193 | 0.38 | 1.01 (0.93–1.11) | 0.782 | 0.871 | 1.01 (0.92–1.11) | 0.785 | 0.885 |
| Clostridium | 1485 | β-glucuronidase activity | 1.02 (0.67–1.55) | 0.933 | 0.933 | 1.07 (0.65–1.74) | 0.797 | 0.871 | 1.05 (0.64–1.75) | 0.836 | 0.899 |
| *Propionibacterium* | *1743* | β-glucuronidase activity | *Below relative abundance prevalence threshold* | | | | | | | | |
| *Citrobacter* | *544* | β-glucuronidase activity | *Below relative abundance prevalence threshold* | | | | | | | | |
| *Marvinbryantia* | *248744* | β-glucuronidase activity | *Below relative abundance prevalence threshold* | | | | | | | | |
| *Dermabacter* | *36739* | β-glucuronidase activity | *Not found* | | | | | | | | |
| *Edwardsiella* | *635* | β-glucuronidase activity | *Not found* | | | | | | | | |
| *Tannerella* | *195950* | β-glucuronidase activity | *Not found* | | | | | | | | |
| **Species** | **NCBI ID** | | | | | | | | | | |
| Bifidobacterium bifidum (SGB17256) | 1681 | β-glucuronidase and β-glucusidase activity | 0.94 (0.86–1.03) | 0.189 | 0.380 | 0.87 (0.77–0.97) | ***0.017*** | 0.342 | 0.86 (0.76–0.97) | ***0.016*** | 0.293 |
| Adlercreutzia equolifaciens (SGB14797) | 446660 | Equol producer | 0.86 (0.79–0.94) | ***0.001*** | ***0.054*** | 0.88 (0.79–0.98) | ***0.018*** | 0.342 | 0.88 (0.79–0.98) | ***0.019*** | 0.293 |
| Lachnospira eligens (SGB5082) | 39485 | β-glucusidase activity | 0.89 (0.81–0.96) | ***0.005*** | ***0.054*** | 0.89 (0.80–0.98) | ***0.019*** | 0.342 | 0.88 (0.79–0.97) | ***0.014*** | 0.293 |
| Clostridium butyricum (SGB4037) | 1492 | β-glucuronidase and β-glucusidase activity | 1.13 (1.04–1.24) | ***0.007*** | ***0.054*** | 1.14 (1.02–1.29) | ***0.020*** | 0.342 | 1.15 (1.03–1.29) | ***0.019*** | 0.293 |
| Blautia obeum (SGB4811) | 40520 | β-glucusidase activity | 0.78 (0.63–0.91) | ***0.007*** | ***0.054*** | 0.79 (0.61–0.95) | ***0.031*** | 0.342 | 0.79 (0.61–0.95) | ***0.030*** | 0.293 |
| Blautia obeum (SGB4810) | 40520 | β-glucusidase activity | 0.94 (0.88–1.01) | 0.102 | 0.282 | 0.91 (0.83–0.99) | ***0.034*** | 0.342 | 0.90 (0.82–0.99) | ***0.029*** | 0.293 |
| Enterocloster clostridioformis (SGB4760) | 1531 | β-glucuronidase and β-glucusidase activity | 1.18 (1.06–1.33) | ***0.004*** | ***0.054*** | 1.15 (1.02–1.33) | ***0.034*** | 0.342 | 1.17 (1.02–1.35) | ***0.024*** | 0.293 |
| Erysipelatoclostridium ramosum (SGB6744) | 1547 | β-glucuronidase and β-glucusidase activity | 1.09 (1.00–1.19) | ***0.051*** | ***0.192*** | 1.12 (1.01–1.25) | ***0.039*** | 0.346 | 1.13 (1.01–1.26) | ***0.033*** | 0.293 |
| Bacteroides uniformis (SGB1836) | 820 | β-glucuronidase and β-glucusidase activity | 0.94 (0.84–1.05) | 0.271 | 0.427 | 0.88 (0.76–1.00) | 0.051 | 0.361 | 0.87 (0.76–1.00) | ***0.049*** | 0.346 |
| Alistipes communis (SGB2290) | 2585118 | Sulfatase activity | 0.90 (0.84–0.97) | ***0.007*** | ***0.054*** | 0.92 (0.83–1.00) | 0.059 | 0.380 | 0.92 (0.83–1.00) | 0.060 | 0.373 |
| Hungatella hathewayi (SGB4742) | 154046 | Sulfatase activity | 1.23 (1.09–1.42) | ***0.002*** | ***0.054*** | 1.16 (1.00–1.38) | 0.065 | 0.387 | 1.17 (1.00–1.39) | 0.063 | 0.373 |
| Faecalibacterium prausnitzii (SGB15332) | 853 | β-glucuronidase activity | 0.94 (0.88–1.00) | ***0.036*** | ***0.170*** | 0.94 (0.87–1.01) | 0.082 | 0.449 | 0.93 (0.87–1.01) | 0.073 | 0.401 |
| Ruminococcus gnavus (SGB4584) | 33038 | β-glucuronidase and 3β-HSD activity | 1.08 (1.01–1.17) | ***0.032*** | ***0.170*** | 1.08 (0.99–1.19) | 0.100 | 0.483 | 1.09 (0.99–1.20) | 0.087 | 0.44 |
| Lacticaseibacillus rhamnosus (SGB7144) | 47715 | β-glucuronidase activity | 0.92 (0.81–1.03) | 0.165 | 0.355 | 0.89 (0.77–1.02) | 0.102 | 0.483 | 0.89 (0.76–1.02) | 0.101 | 0.448 |
| Coprococcus eutactus (SGB5121) | 33043 | β-glucusidase activity | 0.93 (0.86–1.00) | 0.063 | 0.201 | 0.93 (0.85–1.02) | 0.123 | 0.483 | 0.93 (0.84–1.02) | 0.115 | 0.448 |
| Eggerthella lenta (SGB14809) | 84112 | 3β-HSD and 17β-HSD activity | 1.10 (1.01–1.21) | ***0.035*** | ***0.170*** | 1.09 (0.98–1.23) | 0.127 | 0.483 | 1.10 (0.98–1.24) | 0.107 | 0.448 |
| Roseburia hominis (SGB4936) | 301301 | β-glucuronidase and β-glucusidase activity | 0.90 (0.82–0.99) | ***0.030*** | ***0.170*** | 0.92 (0.82–1.02) | 0.134 | 0.483 | 0.91 (0.81–1.02) | 0.120 | 0.448 |
| Faecalibacterium prausnitzii (SGB15323) | 853 | β-glucuronidase activity | 0.93 (0.85–1.00) | 0.068 | 0.201 | 0.93 (0.84–1.02) | 0.136 | 0.483 | 0.92 (0.83–1.02) | 0.128 | 0.455 |
| Lacticaseibacillus paracasei (SGB7142) | 1597 | Equol producer | 1.05 (0.90–1.23) | 0.512 | 0.673 | 1.14 (0.94–1.40) | 0.181 | 0.538 | 1.15 (0.95–1.42) | 0.160 | 0.494 |
| Faecalibacterium prausnitzii (SGB15322) | 853 | β-glucuronidase activity | 1.03 (0.95–1.13) | 0.460 | 0.653 | 1.07 (0.97–1.18) | 0.182 | 0.538 | 1.07 (0.97–1.19) | 0.179 | 0.529 |
| Faecalibacterium prausnitzii (SGB15317) | 853 | β-glucuronidase activity | 0.9 (0.84–0.97) | ***0.005*** | ***0.054*** | 0.95 (0.87–1.03) | 0.215 | 0.584 | 0.95 (0.87–1.03) | 0.205 | 0.554 |
| Slackia isoflavoniconvertens (SGB14773) | 498718 | Equol producer | 1.04 (0.93–1.16) | 0.504 | 0.673 | 1.08 (0.96–1.23) | 0.224 | 0.584 | 1.08 (0.96–1.23) | 0.220 | 0.555 |
| Eubacterium siraeum (SGB4198) | 39492 | β-glucusidase activity | 0.96 (0.90–1.02) | 0.213 | 0.380 | 0.95 (0.88–1.03) | 0.236 | 0.584 | 0.95 (0.88–1.03) | 0.243 | 0.555 |
| Faecalibacterium prausnitzii (SGB15318) | 853 | β-glucuronidase activity | 0.91 (0.85–0.98) | ***0.017*** | ***0.109*** | 0.95 (0.87–1.03) | 0.238 | 0.584 | 0.95 (0.86–1.03) | 0.211 | 0.554 |
| Bifidobacterium pseudocatenulatum (SGB17237) | 28026 | β-glucusidase activity | 1.08 (1.00–1.16) | ***0.055*** | ***0.196*** | 1.06 (0.96–1.16) | 0.246 | 0.584 | 1.06 (0.96–1.16) | 0.232 | 0.555 |
| Coprococcus eutactus (SGB5117) | 33043 | β-glucusidase activity | 0.96 (0.9–1.03) | 0.270 | 0.427 | 0.95 (0.87–1.03) | 0.248 | 0.584 | 0.95 (0.86–1.03) | 0.209 | 0.554 |
| Escherichia coli (SGB10068) | 562 | β-glucuronidase, β-glucusidase, sulfatase activity | 1.14 (1.04–1.25) | ***0.005*** | ***0.054*** | 1.06 (0.95–1.18) | 0.285 | 0.621 | 1.06 (0.95–1.19) | 0.277 | 0.595 |
| Parabacteroides johnsonii (SGB1948) | 387661 | β-glucuronidase activity | 1.07 (0.96–1.19) | 0.239 | 0.395 | 1.07 (0.95–1.22) | 0.289 | 0.621 | 1.07 (0.95–1.22) | 0.276 | 0.595 |
| Coprococcus eutactus (SGB5118) | 33043 | β-glucusidase activity | 0.95 (0.85–1.05) | 0.331 | 0.499 | 0.94 (0.82–1.06) | 0.320 | 0.668 | 0.94 (0.82–1.07) | 0.343 | 0.695 |
| Faecalibacterium prausnitzii (SGB15333) | 853 | β-glucuronidase activity | 0.96 (0.86–1.06) | 0.379 | 0.549 | 0.94 (0.83–1.06) | 0.354 | 0.698 | 0.94 (0.83–1.06) | 0.328 | 0.685 |
| Eubacterium rectale (SGB4933) | 39491 | β-glucusidase activity | 0.94 (0.87–1.01) | 0.120 | 0.293 | 0.96 (0.87–1.06) | 0.414 | 0.731 | 0.96 (0.87–1.05) | 0.372 | 0.696 |
| Roseburia inulinivorans (SGB4940) | 360807 | β-glucuronidase and β-glucusidase activity | 0.95 (0.87–1.03) | 0.215 | 0.380 | 0.96 (0.86–1.07) | 0.438 | 0.731 | 0.96 (0.86–1.07) | 0.451 | 0.717 |
| Bacteroides fragilis (SGB1855) | 817 | β-glucuronidase, β-glucusidase, sulfatase, 3β-HSD and 17β-HSD activity | 1.02 (0.95–1.1) | 0.572 | 0.726 | 1.03 (0.95–1.12) | 0.442 | 0.731 | 1.03 (0.95–1.12) | 0.461 | 0.717 |
| Faecalibacterium prausnitzii (SGB15342) | 853 | β-glucuronidase activity | 0.93 (0.85–1.01) | 0.109 | 0.282 | 0.96 (0.86–1.07) | 0.457 | 0.731 | 0.96 (0.85–1.07) | 0.419 | 0.717 |
| Collinsella aerofaciens (SGB14535_group) | 74426 | β-glucuronidase activity | 0.95 (0.88–1.01) | 0.111 | 0.282 | 0.97 (0.89–1.05) | 0.462 | 0.731 | 0.97 (0.89–1.05) | 0.445 | 0.717 |
| Bifidobacterium adolescentis (SGB17244_group) | 1680 | β-glucuronidase and β-glucusidase activity | 0.99 (0.93–1.05) | 0.636 | 0.766 | 0.97 (0.91–1.05) | 0.475 | 0.731 | 0.97 (0.9–1.04) | 0.431 | 0.717 |
| Intestinibacter bartlettii (SGB6140) | 261299 | β-glucuronidase activity | 0.96 (0.89–1.04) | 0.304 | 0.469 | 0.97 (0.88–1.06) | 0.478 | 0.731 | 0.97 (0.88–1.06) | 0.472 | 0.717 |
| Roseburia intestinalis (SGB4951) | 166486 | β-glucuronidase and β-glucusidase activity | 0.96 (0.89–1.03) | 0.217 | 0.380 | 0.97 (0.89–1.06) | 0.517 | 0.754 | 0.97 (0.89–1.06) | 0.486 | 0.719 |
| Pseudoflavonifractor capillosus (SGB15140) | 106588 | β-glucuronidase activity | 0.93 (0.83–1.04) | 0.222 | 0.380 | 0.96 (0.83–1.1) | 0.527 | 0.754 | 0.96 (0.83–1.11) | 0.549 | 0.745 |
| Parabacteroides merdae (SGB1949) | 46503 | β-glucuronidase activity | 0.98 (0.91–1.04) | 0.491 | 0.673 | 0.97 (0.9–1.06) | 0.531 | 0.754 | 0.97 (0.9–1.06) | 0.520 | 0.745 |
| Blautia obeum (SGB4809) | 40520 | β-glucusidase activity | 0.98 (0.92–1.05) | 0.632 | 0.766 | 1.03 (0.94–1.12) | 0.556 | 0.766 | 1.03 (0.94–1.12) | 0.556 | 0.745 |
| Ruminococcus bicirculans (SGB4262) | 1160721 | β-glucusidase activity | 0.96 (0.9–1.02) | 0.184 | 0.380 | 0.98 (0.91–1.05) | 0.561 | 0.766 | 0.98 (0.91–1.05) | 0.560 | 0.745 |
| Hungatella hathewayi (SGB4741) | 154046 | Sulfatase activity | 1.1 (0.97–1.25) | 0.131 | 0.311 | 1.04 (0.9–1.22) | 0.591 | 0.792 | 1.05 (0.9–1.23) | 0.534 | 0.745 |
| Adlercreutzia equolifaciens (SGB14798) | 446660 | Equol producer | 0.95 (0.88–1.03) | 0.203 | 0.380 | 0.98 (0.89–1.07) | 0.609 | 0.801 | 0.97 (0.89–1.07) | 0.593 | 0.752 |
| Bifidobacterium longum (SGB17248) | 216816 | β-glucuronidase and β-glucusidase activity, equol production | 1.01 (0.95–1.09) | 0.667 | 0.776 | 0.98 (0.91–1.06) | 0.634 | 0.804 | 0.98 (0.9–1.06) | 0.643 | 0.787 |
| Faecalibacterium prausnitzii (SGB15316) | 853 | β-glucuronidase activity | 0.94 (0.85–1.02) | 0.159 | 0.353 | 0.98 (0.87–1.09) | 0.660 | 0.808 | 0.97 (0.86–1.08) | 0.567 | 0.745 |
| Bacteroides thetaiotaomicron (SGB1861) | 818 | β-glucuronidase, β-glucusidase, and sulfatase activity | 0.99 (0.91–1.08) | 0.901 | 0.928 | 1.02 (0.92–1.12) | 0.730 | 0.864 | 1.02 (0.92–1.13) | 0.731 | 0.865 |

| | | | | | | | | | | | |
|---|---|---|---|---|---|---|---|---|---|---|---|
| Parabacteroides distasonis (SGB1934) | 823 | β-glucuronidase and β-glucusidase activity | 1.01 (0.92–1.1) | 0.877 | 0.916 | 0.98 (0.88–1.1) | 0.749 | 0.871 | 0.98 (0.88–1.11) | 0.781 | 0.885 |
| Roseburia faecis (SGB4925) | 301302 | β-glucuronidase and β-glucusidase activity | 0.98 (0.92–1.05) | 0.569 | 0.726 | 0.99 (0.92–1.06) | 0.762 | 0.871 | 0.99 (0.92–1.07) | 0.780 | 0.885 |
| Hungatella hathewayi (SGB4614) | 154046 | Sulfatase activity | 0.99 (0.89–1.10) | 0.861 | 0.913 | 0.98 (0.86–1.12) | 0.797 | 0.871 | 0.99 (0.86–1.13) | 0.833 | 0.899 |
| Faecalibacterium prausnitzii (SGB15326) | 853 | β-glucuronidase activity | 0.94 (0.88–1.00) | 0.059 | 0.199 | 0.99 (0.91–1.08) | 0.850 | 0.913 | 0.99 (0.91–1.08) | 0.831 | 0.899 |
| Phocaeicola vulgatus (SGB1814) | 39485 | β-glucuronidase and β-glucusidase activity | 0.99 (0.91–1.07) | 0.773 | 0.871 | 0.99 (0.91–1.09) | 0.861 | 0.913 | 0.99 (0.91–1.09) | 0.876 | 0.915 |
| Bacteroides ovatus (SGB1871) | 28116 | β-glucuronidase and β-glucusidase activity | 1.03 (0.94–1.14) | 0.499 | 0.673 | 1 (0.89–1.11) | 0.932 | 0.959 | 1 (0.89–1.12) | 0.954 | 0.981 |
| Akkermansia muciniphila (SGB9226) | 239935 | β-glucuronidase activity | 1.01 (0.94–1.07) | 0.853 | 0.913 | 1 (0.92–1.08) | 0.984 | 0.984 | 1 (0.92–1.08) | 0.971 | 0.985 |
| *Adlercreutzia mucosicola* | *580026* | *Equol producer* | *Below relative abundance prevalence threshold* | | | | | | | | |
| *Bifidobacterium angulatum* | *1683* | *β-glucuronidase and β-glucusidase activity* | *Below relative abundance prevalence threshold* | | | | | | | | |
| *Bifidobacterium breve* | *1685* | *β-glucuronidase and β-glucusidase activity, equol production* | *Below relative abundance prevalence threshold* | | | | | | | | |
| *Bifidobacterium dentium* | *1689* | *β-glucuronidase activity* | *Below relative abundance prevalence threshold* | | | | | | | | |
| *Clostridium paraputrificum* | *29363* | *β-glucuronidase and β-glucusidase activity* | *Below relative abundance prevalence threshold* | | | | | | | | |
| *Clostridium perfringens* | *1502* | *β-glucuronidase and β-glucusidase activity* | *Below relative abundance prevalence threshold* | | | | | | | | |
| *Collinsella massiliensis* | *1232426* | *Sulfatase activity* | *Below relative abundance prevalence threshold* | | | | | | | | |
| *Enterococcus faecalis* | *1351* | *β-glucuronidase and β-glucusidase activity* | *Below relative abundance prevalence threshold* | | | | | | | | |
| *Enterococcus faecium* | *1352* | *β-glucosidase activity* | *Below relative abundance prevalence threshold* | | | | | | | | |
| *Marvinbryantia formatexigens* | *168384* | *β-glucuronidase activity* | *Below relative abundance prevalence threshold* | | | | | | | | |
| *Lactobacillus acidophilus* | *1579* | *β-glucuronidase and β-glucusidase activity* | *Below relative abundance prevalence threshold* | | | | | | | | |
| *Lactobacillus gasseri* | *1596* | *β-glucuronidase activity* | *Below relative abundance prevalence threshold* | | | | | | | | |
| *Latilactobacillus sakei* | *1599* | *Equol producer* | *Below relative abundance prevalence threshold* | | | | | | | | |
| *Paraclostridium bifermentans* | *1490* | *β-glucuronidase and β-glucusidase activity* | *Below relative abundance prevalence threshold* | | | | | | | | |
| *Pediococcus pentosaceus* | *1255* | *Equol producer* | *Below relative abundance prevalence threshold* | | | | | | | | |
| *Proteus mirabilis* | *584* | *Equol producer* | *Below relative abundance prevalence threshold* | | | | | | | | |
| *Slackia equolifaciens* | *498718* | *Equol producer* | *Below relative abundance prevalence threshold* | | | | | | | | |
| *Assacharobacter celatus* | *394340* | *Equol producer* | *Not found* | | | | | | | | |
| *Bifidobacterium pseudolongum* | *1694* | *β-glucuronidase and β-glucusidase activity* | *Not found* | | | | | | | | |
| *Butyrivibrio fibrisolvens* | *831* | *β-glucosidase activity* | *Not found* | | | | | | | | |
| *Clostridium innocuum* | *1522* | *β-glucuronidase, β-glucusidase, and 3β-HSD activity* | *Not found* | | | | | | | | |
| *Clostridium septicum* | *1504* | *β-glucosidase activity* | *Not found* | | | | | | | | |
| *Clostridium sporongenes* | *1509* | *β-glucosidase activity* | *Not found* | | | | | | | | |
| *Clostridium sp. Marseille-P2538* | *1816694* | *β-glucosidase activity* | *Not found* | | | | | | | | |
| *Coprococcus sp. ART55/1* | *751585* | *β-glucosidase activity* | *Not found* | | | | | | | | |
| *Coprococcus sp. L2-50* | *NA* | *β-glucosidase activity* | *Not found* | | | | | | | | |
| *Eggerthella sp. CAG:298* | *1262876* | *3β-HSD activity* | *Not found* | | | | | | | | |
| *Lactobacillus intestinalis* | *151781* | *Equol producer* | *Not found* | | | | | | | | |
| *Lactococcus garvieae 20–92* | *1363* | *Equol producer* | *Not found* | | | | | | | | |
| *Peptococcus niger* | *2741* | *Sulfatase activity* | *Not found* | | | | | | | | |
| *Streptococcus agalactiae* | *1311* | *β-glucuronidase activity* | *Not found* | | | | | | | | |
| *Subdoligranulum variabile* | *214851* | *β-glucuronidase activity* | *Not found* | | | | | | | | |
| **Pathway** | **Path code** | | | | | | | | | | |
| Superpathway of β-D-glucuronosides degradation | GLUCUROCAT-PWY | | 0.8 (0.3–2.13) | 0.660 | 0.772 | 1.02 (0.31–3.28) | 0.974 | 0.974 | 1.01 (0.3–3.25) | 0.991 | 0.991 |
| Cholesterol degradation to androstenedione III (anaerobic) | PWY-8151 | | *Not found* | | | | | | | | |
| Androstenedione degradation II (anaerobic) | PWY-8152 | | *Not found* | | | | | | | | |
| Testosterone degradation (anaerobic) | PWY-8155 | | *Not found* | | | | | | | | |
| Testosterone and androsterone degradation to androst-4-en-3,17-dione | PWY-6943 | | *Not found* | | | | | | | | |
| β-D-glucuronide and D-glucuronate degradation | PWY-7427 | | *Not found* | | | | | | | | |
| **Enzyme** | **EC number** | | | | | | | | | | |
| Beta-glucuronidase | 3.2.1.31 | | 1.11 (0.53–2.31) | 0.788 | 0.867 | 1.05 (0.45–2.49) | 0.906 | 0.935 | 1.08 (0.45–2.57) | 0.867 | 0.918 |
| Beta-glucosidase | 3.2.1.21 | | 2.57 (0.75–9.53) | 0.142 | 0.323 | 3.01 (0.7–14.36) | 0.150 | 0.494 | 3.06 (0.7–14.79) | 0.149 | 0.468 |
| Arylsulfatase | 3.1.6.1 | | 1.76 (0.97–3.27) | 0.067 | 0.201 | 1.51 (0.75–3.1) | 0.255 | 0.584 | 1.54 (0.75–3.21) | 0.238 | 0.555 |
| Aryl-sulfate sulfotransferase | 2.8.2.22 | | 0.99 (0.85–1.14) | 0.846 | 0.915 | 0.92 (0.76–1.1) | 0.348 | 0.730 | 0.92 (0.76–1.1) | 0.365 | 0.723 |
| 17-beta-estradiol 17-dehydrogenase | 1.1.1.62 | | 1.01 (0.81–1.26) | 0.921 | 0.933 | 0.95 (0.72–1.23) | 0.685 | 0.838 | 0.95 (0.72–1.24) | 0.686 | 0.838 |
| Bile acid hydrolase | 3.5.1.24 | | 1.44 (0.39–5.45) | 0.584 | 0.727 | 1.81 (0.39–8.45) | 0.446 | 0.731 | 1.82 (0.39–8.66) | 0.444 | 0.717 |
| 3-alpha-(17-beta)-hydroxysteroid dehydrogenase (NAD(+)) | 1.1.1.239 | | *Below relative abundance prevalence threshold* | | | | | | | | |
| 3-beta-(or 20-alpha)-hydroxysteroid dehydrogenase | 1.1.1.210 | | *Below relative abundance prevalence threshold* | | | | | | | | |
| Steryl-sulfatase | 3.1.6.2 | | *Not found* | | | | | | | | |
| Aryl sulfotransferase | 2.8.2.1 | | *Not found* | | | | | | | | |
| 3-alpha-hydroxysteroid dehydrogenase | 1.1.1.213 | | *Not found* | | | | | | | | |
| 3-beta-hydroxysteroid 3-dehydrogenase | 1.1.1.270 | | *Not found* | | | | | | | | |
| Transferred entry 1.1.1.87 (now 17β-hydroxysteroid dehydrogenase type 2) | 1.1.1.155 | | *Not found* | | | | | | | | |
| 20-alpha-hydroxysteroid dehydrogenase | 1.1.1.149 | | *Not found* | | | | | | | | |
| Unspecific monooxygenase | 1.14.14.1 | | *Not found* | | | | | | | | |
| 4-methoxybenzoate monooxygenase (O-demethylase) | 1.14.99.15 | | *Not found* | | | | | | | | |
| 2'-hydroxydaidzein reductase | 1.3.1.51 | | *Not found* | | | | | | | | |

**Supplementary Table 3.** XGBoost performance and top 50 results for each metagenomic feature type (genera, species, pathways, enzymes) contributing to the prediction of breast cancer status in both adjusted and unadjusted models.

| | GENUS | | SPECIES | |
|---|---|---|---|---|
| | Unadjusted | Confounders included as features | Unadjusted | Confounders included as features |
| *pesudo-R2* | *0.153* | *0.188* | *0.163* | *0.136* |
| *AUC* | *0.723* | *0.756* | *0.735* | *0.714* |
| *sensitivity* | *0.686* | *0.800* | *0.900* | *0.957* |
| *specificity* | *0.743* | *0.657* | *0.329* | *0.229* |

**GENUS**

| Unadjusted | | | Confounders included as features | | |
|---|---|---|---|---|---|
| Feature | Scaled Importance | Proportion | Feature | Scaled Importance | Proportion |
| g__Alistipes | 1.00 | 0.063 | g__Blautia | 1.00 | 0.066 |
| g__Blautia | 0.91 | 0.058 | g__Hungatella | 0.89 | 0.058 |
| g__Hungatella | 0.87 | 0.055 | g__Alistipes | 0.83 | 0.055 |
| g__Candidatus_Avimonas | 0.58 | 0.037 | g__GGB3057 | 0.57 | 0.038 |
| g__GGB3057 | 0.44 | 0.028 | g__Candidatus_Avimonas | 0.55 | 0.036 |
| g__Faecalicatena | 0.40 | 0.025 | g__GGB79634 | 0.34 | 0.022 |
| g__GGB13404 | 0.38 | 0.024 | g__Lachnoclostridium | 0.32 | 0.021 |
| g__Intestinimonas | 0.33 | 0.021 | g__Christensenellaceae_unclassified | 0.31 | 0.020 |
| g__Christensenellaceae_unclassified | 0.33 | 0.021 | g__Faecalicatena | 0.30 | 0.020 |
| g__GGB9635 | 0.33 | 0.021 | g__GGB79734 | 0.30 | 0.020 |
| g__GGB9063 | 0.32 | 0.021 | g__Intestinimonas | 0.29 | 0.019 |
| g__GGB79734 | 0.30 | 0.019 | g__GGB9063 | 0.27 | 0.018 |
| g__GGB79634 | 0.30 | 0.019 | g__GGB13404 | 0.26 | 0.017 |
| g__GGB9760 | 0.28 | 0.018 | g__GGB9760 | 0.23 | 0.015 |
| g__GGB80140 | 0.28 | 0.018 | g__GGB80140 | 0.23 | 0.015 |
| g__GGB45432 | 0.25 | 0.016 | g__GGB9635 | 0.22 | 0.015 |
| g__Vescimonas | 0.24 | 0.015 | g__GGB9350 | 0.21 | 0.014 |
| g__Staphylococcus | 0.24 | 0.015 | g__Ruthenibacterium | 0.21 | 0.014 |
| g__Eggerthella | 0.24 | 0.015 | g__Dorea | 0.21 | 0.014 |
| g__Lachnoclostridium | 0.23 | 0.015 | ethnic_collapsed.White | 0.20 | 0.013 |
| g__Anaerobutyricum | 0.22 | 0.014 | g__Clostridium | 0.20 | 0.013 |
| g__Ruthenibacterium | 0.22 | 0.014 | ethnic_collapsed.Non-White | 0.20 | 0.013 |
| g__Adlercreutzia | 0.22 | 0.014 | g__GGB9237 | 0.20 | 0.013 |
| g__GGB9237 | 0.21 | 0.013 | s_age | 0.20 | 0.013 |
| g__Candidatus_Cibionibacter | 0.21 | 0.013 | g__Staphylococcus | 0.20 | 0.013 |
| g__GGB9350 | 0.19 | 0.012 | g__Anaerobutyricum | 0.20 | 0.013 |
| g__Lachnospiraceae_unclassified | 0.19 | 0.012 | g__Vescimonas | 0.19 | 0.013 |
| g__Clostridium | 0.19 | 0.012 | g__GGB51959 | 0.19 | 0.012 |
| g__GGB9707 | 0.19 | 0.012 | g__Intestinibacter | 0.18 | 0.012 |
| g__Intestinibacter | 0.18 | 0.012 | g__Faecalibacillus | 0.18 | 0.012 |
| g__Parabacteroides | 0.17 | 0.011 | g__GGB9707 | 0.17 | 0.011 |
| g__Faecalibacillus | 0.17 | 0.011 | g__Collinsella | 0.16 | 0.011 |
| g__Mediterraneibacter | 0.16 | 0.010 | g__Phascolarctobacterium | 0.15 | 0.010 |
| g__Dorea | 0.16 | 0.010 | g__Lachnospiraceae_unclassified | 0.15 | 0.010 |
| g__GGB9719 | 0.16 | 0.010 | g__GGB9712 | 0.15 | 0.010 |
| g__Firmicutes_unclassified | 0.16 | 0.010 | g__Lacrimispora | 0.14 | 0.010 |
| g__Phascolarctobacterium | 0.16 | 0.010 | g__Eggerthella | 0.14 | 0.009 |
| g__Bifidobacterium | 0.15 | 0.010 | g__Bacteroides | 0.14 | 0.009 |
| g__GGB3819 | 0.15 | 0.010 | g__Bilophila | 0.14 | 0.009 |
| g__GGB9712 | 0.15 | 0.010 | g__Clostridiaceae_unclassified | 0.14 | 0.009 |
| g__Lachnospira | 0.14 | 0.009 | g__GGB58158 | 0.13 | 0.009 |
| g__Eubacteriales_Family_XIII_Incertae | 0.13 | 0.009 | g__Bifidobacterium | 0.13 | 0.009 |
| g__Gemmiger | 0.13 | 0.009 | g__Eubacteriales_Family_XIII_Incertae | 0.13 | 0.008 |
| g__Streptococcus | 0.13 | 0.008 | g__GGB45432 | 0.12 | 0.008 |
| g__GGB58158 | 0.13 | 0.008 | g__Mediterraneibacter | 0.12 | 0.008 |
| g__Solibaculum | 0.12 | 0.008 | g__Firmicutes_unclassified | 0.12 | 0.008 |
| g__Bilophila | 0.12 | 0.008 | g__Escherichia | 0.11 | 0.008 |
| g__Gordonibacter | 0.12 | 0.008 | g__Candidatus_Cibionibacter | 0.11 | 0.007 |
| g__GGB9608 | 0.12 | 0.008 | g__Parabacteroides | 0.11 | 0.007 |
| g__Eubacteriales_unclassified | 0.11 | 0.007 | g__Oscillibacter | 0.11 | 0.007 |

**SPECIES**

<table>
<tr><th colspan="3">Unadjusted</th><th colspan="3">Confounders included as features</th></tr>
<tr><th>Feature</th><th>Scaled Importance</th><th>Proportion</th><th>Feature</th><th>Scaled Importance</th><th>Proportion</th></tr>
<tr><td>s__Anaerotruncus_rubiinfantis|t__SGB25416</td><td>1</td><td>0.056</td><td>s__Anaerotruncus_rubiinfantis|t__SGB25416</td><td>1</td><td>0.066</td></tr>
<tr><td>s__Alistipes_shahii|t__SGB2295</td><td>0.91</td><td>0.051</td><td>s__Alistipes_shahii|t__SGB2295</td><td>0.77</td><td>0.051</td></tr>
<tr><td>s__Lachnospiraceae_bacterium_AM48_27BH|t__SGB4706</td><td>0.77</td><td>0.043</td><td>s__Blautia_obeum|t__SGB4811</td><td>0.52</td><td>0.034</td></tr>
<tr><td>s__Alistipes_finegoldii|t__SGB2301</td><td>0.71</td><td>0.040</td><td>s__Lachnospiraceae_bacterium_AM48_27BH|t__SGB4706</td><td>0.48</td><td>0.032</td></tr>
<tr><td>s__Sutterella_wadsworthensis|t__SGB9286</td><td>0.65</td><td>0.037</td><td>s__Alistipes_finegoldii|t__SGB2301</td><td>0.46</td><td>0.030</td></tr>
<tr><td>s__Blautia_obeum|t__SGB4811</td><td>0.62</td><td>0.035</td><td>s__Mediterraneibacter_massiliensis|t__SGB4595</td><td>0.34</td><td>0.022</td></tr>
<tr><td>s__Candidatus_Avimonas_narfia|t__SGB14941</td><td>0.56</td><td>0.031</td><td>s__GGB9760_SGB15373|t__SGB15373</td><td>0.34</td><td>0.022</td></tr>
<tr><td>s__Alistipes_putredinis|t__SGB2318</td><td>0.56</td><td>0.031</td><td>s__Alistipes_putredinis|t__SGB2318</td><td>0.33</td><td>0.022</td></tr>
<tr><td>s__Mediterraneibacter_massiliensis|t__SGB4595</td><td>0.46</td><td>0.026</td><td>s__Mediterraneibacter_butyricigenes|t__SGB25493</td><td>0.33</td><td>0.021</td></tr>
<tr><td>s__Bacteroides_clarus|t__SGB1832</td><td>0.44</td><td>0.025</td><td>s__Blautia_SGB4815|t__SGB4815</td><td>0.29</td><td>0.019</td></tr>
<tr><td>s__Candidatus_Cibionibacter_quicibialis|t__SGB15286</td><td>0.34</td><td>0.019</td><td>s__Sutterella_wadsworthensis|t__SGB9286</td><td>0.28</td><td>0.019</td></tr>
<tr><td>s__Clostridia_bacterium_UC5_1_1D1|t__SGB14995</td><td>0.32</td><td>0.018</td><td>s__Faecalibacillus_intestinalis|t__SGB6754</td><td>0.28</td><td>0.018</td></tr>
<tr><td>s__Blautia_SGB4815|t__SGB4815</td><td>0.30</td><td>0.017</td><td>s__Candidatus_Avimonas_narfia|t__SGB14941</td><td>0.27</td><td>0.018</td></tr>
<tr><td>s__Mediterraneibacter_butyricigenes|t__SGB25493</td><td>0.30</td><td>0.017</td><td>s_age</td><td>0.27</td><td>0.018</td></tr>
<tr><td>s__Clostridium_fessum|t__SGB4705</td><td>0.29</td><td>0.016</td><td>s__Clostridium_fessum|t__SGB4705</td><td>0.25</td><td>0.017</td></tr>
<tr><td>s__Anaerobutyricum_hallii|t__SGB4532</td><td>0.28</td><td>0.016</td><td>s__Clostridium_sp_AM49_4BH|t__SGB4652</td><td>0.24</td><td>0.016</td></tr>
<tr><td>s__Collinsella_SGB14861|t__SGB14861</td><td>0.27</td><td>0.015</td><td>s__Anaerobutyricum_hallii|t__SGB4532</td><td>0.22</td><td>0.015</td></tr>
<tr><td>s__Dorea_sp_AF36_15AT|t__SGB4552</td><td>0.27</td><td>0.015</td><td>s__Bacteroides_xylanisolvens|t__SGB1867</td><td>0.21</td><td>0.014</td></tr>
<tr><td>s__GGB3057_SGB4059|t__SGB4059</td><td>0.26</td><td>0.015</td><td>s__Christensenellaceae_bacterium_NSJ_63|t__SGB14127</td><td>0.20</td><td>0.013</td></tr>
<tr><td>s__Faecalibacillus_intestinalis|t__SGB6754</td><td>0.26</td><td>0.014</td><td>s__GGB3057_SGB4059|t__SGB4059</td><td>0.20</td><td>0.013</td></tr>
<tr><td>s__Clostridium_saudiense|t__SGB6178</td><td>0.25</td><td>0.014</td><td>s__Clostridium_saudiense|t__SGB6178</td><td>0.19</td><td>0.013</td></tr>
<tr><td>s__Lachnospiraceae_bacterium|t__SGB4782</td><td>0.24</td><td>0.014</td><td>s__GGB9712_SGB15244|t__SGB15244</td><td>0.19</td><td>0.013</td></tr>
<tr><td>s__GGB9760_SGB15373|t__SGB15373</td><td>0.23</td><td>0.013</td><td>s__Candidatus_Cibionibacter_quicibialis|t__SGB15286</td><td>0.19</td><td>0.012</td></tr>
<tr><td>s__Bacteroides_xylanisolvens|t__SGB1867</td><td>0.23</td><td>0.013</td><td>s__Eubacterium_rectale|t__SGB4933</td><td>0.18</td><td>0.012</td></tr>
<tr><td>s__Clostridium_sp_AM49_4BH|t__SGB4652</td><td>0.22</td><td>0.012</td><td>ethnic_collapsed.White</td><td>0.18</td><td>0.012</td></tr>
<tr><td>s__GGB9712_SGB15244|t__SGB15244</td><td>0.21</td><td>0.012</td><td>s__Lachnospira_sp_NSJ_43|t__SGB5087</td><td>0.17</td><td>0.011</td></tr>
<tr><td>s__GGB9063_SGB13982|t__SGB13982</td><td>0.20</td><td>0.011</td><td>ethnic_collapsed.Non-White</td><td>0.17</td><td>0.011</td></tr>
<tr><td>s__GGB80140_SGB15224|t__SGB15224</td><td>0.20</td><td>0.011</td><td>s__Methylobacterium_SGB15164|t__SGB15164</td><td>0.15</td><td>0.010</td></tr>
<tr><td>s__GGB13404_SGB14252|t__SGB14252</td><td>0.19</td><td>0.011</td><td>s__Collinsella_SGB14861|t__SGB14861</td><td>0.15</td><td>0.010</td></tr>
<tr><td>s__Actinomyces_SGB17168|t__SGB17168</td><td>0.18</td><td>0.010</td><td>s__Clostridium_sp_AF36_4|t__SGB4644</td><td>0.15</td><td>0.010</td></tr>
<tr><td>s__Dorea_formicigenerans|t__SGB4575</td><td>0.18</td><td>0.010</td><td>s__Lachnospiraceae_bacterium|t__SGB4781</td><td>0.15</td><td>0.010</td></tr>
<tr><td>s__Lachnospiraceae_bacterium|t__SGB4781</td><td>0.18</td><td>0.010</td><td>s__Intestinimonas_butyriciproducens|t__SGB15126</td><td>0.14</td><td>0.009</td></tr>
<tr><td>s__Christensenellaceae_bacterium_NSJ_63|t__SGB14127</td><td>0.17</td><td>0.010</td><td>s__GGB3005_SGB3996|t__SGB3996</td><td>0.14</td><td>0.009</td></tr>
<tr><td>s__Clostridium_sp_AF36_4|t__SGB4644</td><td>0.17</td><td>0.010</td><td>s__Ruminococcus_sp_AF41_9|t__SGB25497</td><td>0.14</td><td>0.009</td></tr>
<tr><td>s__Blautia_luti|t__SGB4829</td><td>0.17</td><td>0.009</td><td>s__GGB80140_SGB15224|t__SGB15224</td><td>0.14</td><td>0.009</td></tr>
<tr><td>s__Bacteroides_ovatus|t__SGB1871</td><td>0.17</td><td>0.009</td><td>s__Actinomyces_SGB17168|t__SGB17168</td><td>0.14</td><td>0.009</td></tr>
<tr><td>s__GGB3005_SGB3996|t__SGB3996</td><td>0.16</td><td>0.009</td><td>s__Adlercreutzia_equolifaciens|t__SGB14797</td><td>0.13</td><td>0.009</td></tr>
<tr><td>s__Intestinimonas_butyriciproducens|t__SGB15126</td><td>0.16</td><td>0.009</td><td>s__Staphylococcus_SGB6340|t__SGB6340</td><td>0.13</td><td>0.009</td></tr>
<tr><td>s__Lachnospira_sp_NSJ_43|t__SGB5087</td><td>0.16</td><td>0.009</td><td>s__GGB13404_SGB14252|t__SGB14252</td><td>0.13</td><td>0.009</td></tr>
<tr><td>s__Hungatella_hathewayi|t__SGB4614</td><td>0.15</td><td>0.008</td><td>s__Bacteroides_ovatus|t__SGB1871</td><td>0.13</td><td>0.008</td></tr>
<tr><td>s__Blautia_hydrogenotrophica|t__SGB4677</td><td>0.15</td><td>0.008</td><td>s__Blautia_luti|t__SGB4829</td><td>0.13</td><td>0.008</td></tr>
<tr><td>s__GGB9635_SGB15106|t__SGB15106</td><td>0.15</td><td>0.008</td><td>s__GGB79734_SGB15291|t__SGB15291</td><td>0.12</td><td>0.008</td></tr>
<tr><td>s__Escherichia_coli|t__SGB10068</td><td>0.14</td><td>0.008</td><td>s__GGB9522_SGB14921|t__SGB14921</td><td>0.12</td><td>0.008</td></tr>
<tr><td>s__Anaerostipes_hadrus|t__SGB4547</td><td>0.14</td><td>0.008</td><td>s__Lacrimispora_celerecrescens|t__SGB4868</td><td>0.12</td><td>0.008</td></tr>
<tr><td>s__Blautia_glucerasea|t__SGB4804</td><td>0.14</td><td>0.008</td><td>s__Ruthenibacterium_lactatiformans|t__SGB15271</td><td>0.12</td><td>0.008</td></tr>
<tr><td>s__GGB79734_SGB15291|t__SGB15291</td><td>0.13</td><td>0.007</td><td>s__GGB3653_SGB4964|t__SGB4964</td><td>0.12</td><td>0.008</td></tr>
<tr><td>s__Faecalicatena_fissicatena|t__SGB4871</td><td>0.13</td><td>0.007</td><td>s__Phascolarctobacterium_SGB4573|t__SGB4573</td><td>0.11</td><td>0.008</td></tr>
<tr><td>s__Eggerthella_lenta|t__SGB14809</td><td>0.12</td><td>0.007</td><td>s__Lachnospiraceae_bacterium|t__SGB4782</td><td>0.11</td><td>0.007</td></tr>
<tr><td>s__GGB3653_SGB4964|t__SGB4964</td><td>0.12</td><td>0.007</td><td>s__Parabacteroides_distasonis|t__SGB1934</td><td>0.11</td><td>0.007</td></tr>
<tr><td>s__Bacteroides_caccae|t__SGB1877</td><td>0.11</td><td>0.006</td><td>s__Faecalicatena_fissicatena|t__SGB4871</td><td>0.11</td><td>0.007</td></tr>
</table>

| | PATHWAYS Unadjusted | PATHWAYS Confounders included as features | ENZYMES Unadjusted | ENZYMES Confounders included as features |
|---|---|---|---|---|
| pesudo-R2 | 0.074 | 0.032 | 0.297 | 0.246 |
| AUC | 0.663 | 0.648 | 0.816 | 0.783 |
| sensitivity | 0.857 | 0.886 | 0.886 | 0.843 |
| specificity | 0.429 | 0.386 | 0.529 | 0.543 |

| PATHWAYS | | | | | | ENZYMES | | | | | |
|---|---|---|---|---|---|---|---|---|---|---|---|
| Unadjusted | | | Confounders included as features | | | Unadjusted | | | Confounders included as features | | |
| Feature | Scaled Importance | Proportion | Feature | Scaled Importance | Proportion | Feature | Scaled Importance | Proportion | Feature | Scaled Importance | Proportion |
| PWY-7992: superpathway of menaquinol-8 biosynthesis III | 1.00 | 0.058 | PWY-7992: superpathway of menaquinol-8 biosynthesis III | 1.00 | 0.056 | 1.8.98.1 | 1.00 | 0.050 | 1.8.98.1 | 1.00 | 0.060 |
| RIBOSYN2-PWY: flavin biosynthesis I (bacteria and plants) | 0.81 | 0.047 | RIBOSYN2-PWY: flavin biosynthesis I (bacteria and plants) | 0.74 | 0.041 | 3.5.1.81 | 0.79 | 0.040 | 2.7.1.113 | 0.46 | 0.027 |
| PWY-7883: anhydromuropeptides recycling II | 0.74 | 0.043 | PWY-5130: 2-oxobutanoate degradation I | 0.57 | 0.032 | 2.7.1.113 | 0.68 | 0.034 | 3.5.1.81 | 0.42 | 0.025 |
| PWY-5130: 2-oxobutanoate degradation I | 0.67 | 0.039 | PWY-7883: anhydromuropeptides recycling II | 0.56 | 0.032 | 2.5.1.120 | 0.66 | 0.033 | 6.3.4.15 | 0.39 | 0.023 |
| PWY-6901: superpathway of glucose and xylose degradation | 0.61 | 0.035 | ethnic_collapsed.Non-White | 0.55 | 0.031 | 3.4.13.21 | 0.64 | 0.032 | 2.5.1.120 | 0.35 | 0.021 |
| PWY-6876: isopropanol biosynthesis (engineered) | 0.58 | 0.034 | PWY-5973: cis-vaccenate biosynthesis | 0.44 | 0.024 | 4.2.1.115 | 0.54 | 0.027 | 3.4.13.21 | 0.35 | 0.021 |
| PWY-8131: 5'-deoxyadenosine degradation II | 0.55 | 0.032 | PWY-7761: NAD salvage pathway II (PNC IV cycle) | 0.41 | 0.023 | 3.4.22.8 | 0.53 | 0.026 | 1.2.99.5 | 0.30 | 0.018 |
| PWY-7761: NAD salvage pathway II (PNC IV cycle) | 0.54 | 0.031 | PWY-6876: isopropanol biosynthesis (engineered) | 0.37 | 0.021 | 1.4.99.6 | 0.52 | 0.026 | 1.4.99.6 | 0.27 | 0.016 |
| P42-PWY: incomplete reductive TCA cycle | 0.49 | 0.028 | P42-PWY: incomplete reductive TCA cycle | 0.37 | 0.021 | 1.2.99.5 | 0.52 | 0.026 | 2.4.1.329 | 0.24 | 0.014 |
| METHGLYUT-PWY: superpathway of methylglyoxal degradation | 0.48 | 0.028 | PWY-6901: superpathway of glucose and xylose degradation | 0.35 | 0.019 | 2.4.1.319 | 0.46 | 0.023 | 3.4.22.8 | 0.23 | 0.014 |
| PWY-7200: superpathway of pyrimidine deoxyribonucleoside salvage | 0.37 | 0.022 | PWY-7013: (S)-propane-1,2-diol degradation | 0.33 | 0.018 | 2.4.1.329 | 0.38 | 0.019 | 2.4.1.319 | 0.21 | 0.012 |
| PWY-7371: 1,4-dihydroxy-6-naphthoate biosynthesis II | 0.36 | 0.021 | PWY-8131: 5'-deoxyadenosine degradation II | 0.30 | 0.017 | 1.10.9.1 | 0.37 | 0.019 | 4.2.1.115 | 0.20 | 0.012 |
| PWY-7013: (S)-propane-1,2-diol degradation | 0.30 | 0.018 | PWY-6588: pyruvate fermentation to acetone | 0.30 | 0.017 | 6.3.2.45 | 0.37 | 0.019 | 1.18.1.2 | 0.17 | 0.010 |
| PWY-6588: pyruvate fermentation to acetone | 0.30 | 0.018 | METHGLYUT-PWY: superpathway of methylglyoxal degradation | 0.29 | 0.016 | 2.1.1.173 | 0.36 | 0.018 | 6.3.4.2 | 0.17 | 0.010 |
| PWY-5920: superpathway of heme b biosynthesis from glycine | 0.28 | 0.017 | ethnic_collapsed.White | 0.29 | 0.016 | 6.3.4.15 | 0.35 | 0.017 | 2.1.1.173 | 0.17 | 0.010 |
| PWY-621: sucrose degradation III (sucrose invertase) | 0.28 | 0.016 | PWY-7315: dTDP-N-acetylthomosamine biosynthesis | 0.23 | 0.013 | 5.1.3.8 | 0.34 | 0.017 | 1.10.9.1 | 0.16 | 0.010 |
| PWY-7316: dTDP-N-acetylviosamine biosynthesis | 0.28 | 0.016 | GLCMANNANAUT-PWY: superpathway of N-acetylglucosamine, N-acetylma | 0.23 | 0.013 | 2.3.1.57 | 0.34 | 0.017 | 3.5.3.19 | 0.16 | 0.010 |
| P441-PWY: superpathway of N-acetylneuraminate degradation | 0.25 | 0.015 | PWY-7371: 1,4-dihydroxy-6-naphthoate biosynthesis II | 0.23 | 0.013 | 3.2.1.172 | 0.33 | 0.016 | 4.2.1.3 | 0.16 | 0.010 |
| PWY-7332: superpathway of UDP-N-acetylglucosamine-derived O-antigen buildi | 0.25 | 0.014 | THRESYN-PWY: superpathway of L-threonine biosynthesis | 0.22 | 0.013 | 2.1.1.61 | 0.30 | 0.015 | 5.1.3.8 | 0.16 | 0.009 |
| PWY-6803: phosphatidylcholine acyl editing | 0.24 | 0.014 | PWY-5920: superpathway of heme b biosynthesis from glycine | 0.21 | 0.012 | 3.5.5.1 | 0.30 | 0.015 | 2.1.1.61 | 0.16 | 0.009 |
| PENTOSE-P-PWY: pentose phosphate pathway | 0.24 | 0.014 | PWY-6607: guanosine nucleotides degradation I | 0.21 | 0.012 | 4.2.1.45 | 0.28 | 0.014 | 1.1.1.159 | 0.15 | 0.009 |
| PWY-6607: guanosine nucleotides degradation I | 0.23 | 0.013 | NAGLIPASYN-PWY: lipid IVA biosynthesis (E. coli) | 0.21 | 0.011 | 1.18.1.2 | 0.28 | 0.014 | 6.3.2.45 | 0.15 | 0.009 |
| NAGLIPASYN-PWY: lipid IVA biosynthesis (E. coli) | 0.23 | 0.013 | PWY-6703: preQ0 biosynthesis | 0.20 | 0.011 | 2.7.2.11 | 0.27 | 0.014 | 2.2.1.9 | 0.15 | 0.009 |
| PWY-6902: chitin degradation II (Vibrio) | 0.22 | 0.013 | PWY-7234: inosine-5'-phosphate biosynthesis III | 0.20 | 0.011 | 2.3.1.190 | 0.27 | 0.014 | 1.1.1.291 | 0.15 | 0.009 |
| PWY-5677: succinate fermentation to butanoate | 0.21 | 0.012 | PWY-621: sucrose degradation III (sucrose invertase) | 0.19 | 0.011 | 4.2.1.130 | 0.26 | 0.013 | 1.6.5.5 | 0.14 | 0.009 |
| PWY0-1061: superpathway of L-alanine biosynthesis | 0.21 | 0.012 | PWY-6549: L-glutamine biosynthesis III | 0.19 | 0.010 | 1.4.99.5 | 0.25 | 0.012 | 2.7.7.38 | 0.14 | 0.009 |
| PWY490-3: nitrate reduction VI (assimilatory) | 0.20 | 0.012 | RUMP-PWY: formaldehyde oxidation I | 0.19 | 0.010 | 1.1.1.36 | 0.24 | 0.012 | 2.7.1.74 | 0.14 | 0.008 |
| PWY-6703: preQ0 biosynthesis | 0.20 | 0.012 | PWY-7316: dTDP-N-acetylviosamine biosynthesis | 0.19 | 0.010 | 2.8.1.13 | 0.24 | 0.012 | 2.6.1.33 | 0.14 | 0.008 |
| PWY-6518: bile acids epimerization | 0.19 | 0.011 | PWY-7323: superpathway of GDP-mannose-derived O-antigen building bloc | 0.19 | 0.010 | 4.2.1.3 | 0.23 | 0.012 | 3.1.3.27 | 0.14 | 0.008 |
| P108-PWY: pyruvate fermentation to propanoate I | 0.19 | 0.011 | PWY-6803: phosphatidylcholine acyl editing | 0.19 | 0.010 | 1.1.1.130 | 0.22 | 0.011 | 6.3.5.6 | 0.14 | 0.008 |
| HISDEG-PWY: L-histidine degradation I | 0.19 | 0.011 | PWY-6902: chitin degradation II (Vibrio) | 0.18 | 0.010 | 4.2.2.22 | 0.22 | 0.011 | 2.5.1.9 | 0.14 | 0.008 |
| PWY-5384: sucrose degradation IV (sucrose phosphorylase) | 0.19 | 0.011 | PWY0-1061: superpathway of L-alanine biosynthesis | 0.18 | 0.010 | 3.4.21.83 | 0.21 | 0.011 | 3.5.5.1 | 0.13 | 0.008 |
| PWY-8073: lipid IVA biosynthesis (P. putida) | 0.18 | 0.010 | PWY-7237: myo-, chiro- and scyllo-inositol degradation | 0.18 | 0.010 | 1.4.7.1 | 0.21 | 0.010 | 2.3.1.190 | 0.13 | 0.008 |
| CRNFORCAT-PWY: creatinine degradation I | 0.17 | 0.010 | PENTOSE-P-PWY: pentose phosphate pathway | 0.18 | 0.010 | 2.8.1.10 | 0.21 | 0.010 | 4.1.1.37 | 0.13 | 0.008 |
| PWY-1861: formaldehyde assimilation II (assimilatory RuMP Cycle) | 0.17 | 0.010 | PWY-6823: molybdopterin biosynthesis | 0.17 | 0.010 | 2.1.1.43 | 0.20 | 0.010 | 3.2.1.78 | 0.12 | 0.007 |
| PWY-7234: inosine-5'-phosphate biosynthesis III | 0.17 | 0.010 | PWY-6595: superpathway of guanosine nucleotides degradation (plants) | 0.17 | 0.010 | 1.6.99.5 | 0.20 | 0.010 | 5.3.1.4 | 0.12 | 0.007 |
| P125-PWY: superpathway of (R,R)-butanediol biosynthesis | 0.17 | 0.010 | ALLANTOINDEG-PWY: superpathway of allantoin degradation in yeast | 0.17 | 0.009 | 2.7.1.55 | 0.19 | 0.010 | 7.2.4.1 | 0.12 | 0.007 |
| PWY-7315: dTDP-N-acetylthomosamine biosynthesis | 0.16 | 0.009 | PWY-6700: queuosine biosynthesis I (de novo) | 0.17 | 0.009 | 1.1.1.60 | 0.18 | 0.009 | 3.2.1.172 | 0.12 | 0.007 |
| PWY-5265: peptidoglycan biosynthesis II (staphylococci) | 0.16 | 0.009 | P125-PWY: superpathway of (R,R)-butanediol biosynthesis | 0.16 | 0.009 | 3.4.11.4 | 0.18 | 0.009 | 1.5.1.28 | 0.12 | 0.007 |
| PWY-7115: C4 photosynthetic carbon assimilation cycle, NAD-ME type | 0.16 | 0.009 | PWY-7197: pyrimidine deoxyribonucleotide phosphorylation | 0.15 | 0.009 | 2.7.8.20 | 0.18 | 0.009 | 1.6.99.5 | 0.12 | 0.007 |
| HEME-BIOSYNTHESIS-II-1: heme b biosynthesis V (aerobic) | 0.16 | 0.009 | P441-PWY: superpathway of N-acetylneuraminate degradation | 0.15 | 0.008 | 1.2.1.2 | 0.17 | 0.009 | 2.3.1.57 | 0.12 | 0.007 |
| LACTOSECAT-PWY: lactose and galactose degradation I | 0.16 | 0.009 | LACTOSECAT-PWY: lactose and galactose degradation I | 0.15 | 0.008 | 1.6.5.5 | 0.17 | 0.009 | 4.2.1.45 | 0.12 | 0.007 |
| PWY-6572: chondroitin sulfate degradation I (bacterial) | 0.15 | 0.009 | PWY-5136: fatty acid β-oxidation II (plant peroxisome) | 0.15 | 0.008 | 1.2.1.59 | 0.17 | 0.009 | 4.2.1.130 | 0.12 | 0.007 |
| ALLANTOINDEG-PWY: superpathway of allantoin degradation in yeast | 0.15 | 0.009 | PWY-7200: superpathway of pyrimidine deoxyribonucleoside salvage | 0.14 | 0.008 | 5.1.99.1 | 0.17 | 0.008 | 2.5.1.76 | 0.12 | 0.007 |
| PWY-7237: myo-, chiro- and scyllo-inositol degradation | 0.15 | 0.009 | COA-PWY: coenzyme A biosynthesis I (prokaryotic) | 0.14 | 0.008 | 2.6.1.33 | 0.15 | 0.008 | 2.5.1.74 | 0.12 | 0.007 |
| PWY-7434: terminal O-glycans residues modification (via type 2 precursor disac | 0.15 | 0.009 | s_age | 0.13 | 0.008 | 2.3.1.19 | 0.15 | 0.008 | 1.3.99.4 | 0.12 | 0.007 |
| PYRIDNUCSAL-PWY: NAD salvage pathway I (PNC VI cycle) | 0.15 | 0.009 | PWY-3841: folate transformations II (plants) | 0.13 | 0.007 | 2.2.1.9 | 0.15 | 0.007 | 4.4.1.15 | 0.12 | 0.007 |
| PWY-7211: superpathway of pyrimidine deoxyribonucleotides de novo biosynthe | 0.15 | 0.009 | PYRIDNUCSAL-PWY: NAD salvage pathway I (PNC VI cycle) | 0.13 | 0.007 | 7.2.4.1 | 0.15 | 0.007 | 3.5.4.31 | 0.12 | 0.007 |
| PWY-6595: superpathway of guanosine nucleotides degradation (plants) | 0.14 | 0.008 | HISDEG-PWY: L-histidine degradation I | 0.13 | 0.007 | 2.4.1.320 | 0.13 | 0.006 | 3.5.2.2 | 0.11 | 0.007 |
| PWY-6143: CMP-pseudaminate biosynthesis | 0.12 | 0.007 | PWY0-1479: tRNA processing | 0.13 | 0.007 | 3.5.3.19 | 0.13 | 0.006 | ethnic_collapsed. | 0.11 | 0.007 |

**Supplementary Table 4**. Logistic Regression results for top XGBoost predictors from each metagenomic feature type (genera, species, pathways, enzymes).

| **Metagenomic Feature** | **Model 0 (Unadjusted)** n=138 / AIC 183.8 | | **Model 1 (Confounder Set)** n=134 / AIC 169.4 | | **Model 2 (Diet Quality)** n=134 / AIC 174.9 | |
|---|---|---|---|---|---|---|
| | **OR (95% CI)** | **p** | **OR (95% CI)** | **p** | **OR (95% CI)** | **p** |
| **Genus** | | | | | | |
| Christensenellaceae unclassified | 0.83 (0.74–0.93) | ***0.001*** | 0.80 (0.69–0.92) | ***0.002*** | 0.80 (0.69–0.92) | ***0.002*** |
| GGB9635 | 0.89 (0.82–0.95) | ***0.001*** | 0.88 (0.81–0.95) | ***0.003*** | 0.88 (0.80–0.95) | ***0.002*** |
| GGB3057 | 0.81 (0.70–0.91) | ***0.001*** | 0.77 (0.63–0.90) | ***0.003*** | 0.76 (0.63–0.90) | ***0.003*** |
| GGB80140 | 0.86 (0.78–0.94) | ***0.002*** | 0.85 (0.76–0.95) | ***0.005*** | 0.85 (0.76–0.95) | ***0.006*** |
| Intestinimonas | 0.87 (0.79–0.96) | ***0.006*** | 0.85 (0.75–0.96) | ***0.008*** | 0.85 (0.75–0.95) | ***0.008*** |
| GGB13404 | 0.85 (0.77–0.94) | ***0.002*** | 0.85 (0.75–0.96) | ***0.010*** | 0.85 (0.74–0.96) | ***0.010*** |
| Faecalicatena | 1.04 (0.97–1.13) | 0.292 | 1.11 (1.01–1.23) | ***0.035*** | 1.11 (1.01–1.23) | ***0.038*** |
| Candidatus Avimonas | 0.86 (0.78–0.95) | ***0.003*** | 0.89 (0.79–0.99) | ***0.041*** | 0.89 (0.79–1.00) | ***0.044*** |
| GGB79634 | 1.05 (0.95–1.17) | 0.333 | 1.12 (1.00–1.27) | 0.056 | 1.13 (1.00–1.28) | 0.052 |
| GGB79734 | 0.91 (0.85–0.98) | ***0.019*** | 0.92 (0.84–1.01) | 0.081 | 0.92 (0.83–1.01) | 0.071 |
| Hungatella | 1.14 (1.03–1.25) | ***0.009*** | 1.10 (0.98–1.24) | 0.103 | 1.11 (0.98–1.25) | 0.093 |
| GGB9063 | 0.89 (0.81–0.98) | ***0.017*** | 0.91 (0.82–1.02) | 0.114 | 0.91 (0.82–1.02) | 0.119 |
| Alistipes | 0.77 (0.56–0.94) | ***0.043*** | 0.84 (0.64–1.02) | 0.117 | 0.84 (0.64–1.02) | 0.119 |
| Lachnoclostridium | 1.06 (0.98–1.16) | 0.163 | 1.05 (0.94–1.17) | 0.365 | 1.06 (0.95–1.18) | 0.327 |
| Blautia | 1.00 (0.73–1.39) | 0.972 | 1.12 (0.74–1.54) | 0.494 | 1.11 (0.74–1.54) | 0.504 |
| **Species** | | | | | | |
| Anaerotruncus rubiinfantis (SGB25416) | 0.78 (0.68–0.89) | ***0.000*** | 0.76 (0.64–0.89) | ***0.001*** | ***0.75 (0.63–0.89)*** | ***0.001*** |
| GGB3057 SGB4059 (SGB4059) | 0.81 (0.70–0.91) | ***0.001*** | 0.77 (0.63–0.90) | ***0.003*** | ***0.76 (0.63–0.90)*** | ***0.003*** |
| Sutterella wadsworthensis (SGB9286) | 0.87 (0.79–0.95) | ***0.004*** | 0.86 (0.77–0.96) | ***0.007*** | ***0.86 (0.76–0.95)*** | ***0.007*** |
| Lachnospiraceae bacterium AM48-27BH (SGB4706) | 0.86 (0.78–0.94) | ***0.001*** | 0.87 (0.78–0.97) | ***0.011*** | ***0.87 (0.78–0.97)*** | ***0.011*** |
| Alistipes putredinis (SGB2318) | 0.88 (0.81–0.95) | ***0.001*** | 0.89 (0.80–0.97) | ***0.012*** | ***0.88 (0.79–0.97)*** | ***0.009*** |
| Blautia obeum (SGB4811) | 0.78 (0.63–0.91) | ***0.007*** | 0.79 (0.61–0.95) | ***0.031*** | ***0.79 (0.61–0.95)*** | ***0.030*** |
| Candidatus Avimonas narfia (SGB14941) | 0.86 (0.78–0.95) | ***0.003*** | 0.89 (0.79–0.99) | ***0.041*** | ***0.89 (0.79–1.00)*** | ***0.044*** |
| Clostridium fessum (SGB4705) | 0.88 (0.80–0.96) | ***0.007*** | 0.90 (0.81–1.00) | 0.057 | ***0.90 (0.80–1.00)*** | ***0.045*** |
| Mediterraneibacter massiliensis (SGB4595) | 0.87 (0.77–0.97) | ***0.018*** | 0.87 (0.75–1.00) | 0.060 | 0.87 (0.75–1.00) | 0.057 |
| Blautia SGB4815 (SGB4815) | 0.89 (0.81–0.96) | ***0.006*** | 0.91 (0.82–1.01) | 0.068 | 0.90 (0.81–1.00) | 0.061 |
| Alistipes shahii (SGB2295) | 0.93 (0.86–0.99) | ***0.039*** | 0.92 (0.84–1.01) | 0.071 | 0.92 (0.83–1.01) | 0.068 |
| Alistipes finegoldii (SGB2301) | 0.90 (0.83–0.97) | ***0.010*** | 0.92 (0.84–1.01) | 0.098 | 0.92 (0.84–1.01) | 0.098 |
| Faecalibacillus intestinalis (SGB6754) | 0.94 (0.87–1.01) | 0.095 | 0.94 (0.86–1.03) | 0.201 | 0.94 (0.86–1.03) | 0.191 |
| Candidatus Cibionibacter quicibialis (SGB15286) | 0.94 (0.85–1.02) | 0.144 | 0.98 (0.88–1.09) | 0.718 | 0.98 (0.88–1.09) | 0.728 |
| Anaerobutyricum hallii (SGB4532) | 0.96 (0.88–1.03) | 0.270 | 0.99 (0.90–1.10) | 0.909 | 0.99 (0.90–1.09) | 0.848 |
| Mediterraneibacter butyricigenes (SGB25493) | 0.95 (0.86–1.04) | 0.269 | 1.00 (0.90–1.12) | 0.971 | 1.00 (0.90–1.13) | 0.951 |
| **Pathway** | | | | | | |
| PWY-8131: 5'-deoxyadenosine degradation II | 2.18 (1.05–4.83) | ***0.043*** | 3.19 (1.36–8.15) | ***0.010*** | 3.29 (1.39–8.54) | ***0.009*** |
| PWY-5130: 2-oxobutanoate degradation I | 0.52 (0.29–0.84) | ***0.015*** | 0.47 (0.25–0.82) | ***0.015*** | 0.46 (0.24–0.81) | ***0.013*** |
| P42-PWY: incomplete reductive TCA cycle | 0.55 (0.34–0.83) | ***0.008*** | 0.53 (0.30–0.86) | ***0.017*** | 0.52 (0.30–0.85) | ***0.016*** |
| PWY-7761: NAD salvage pathway II (PNC IV cycle) | 0.50 (0.25–0.94) | ***0.039*** | 0.44 (0.20–0.90) | ***0.031*** | 0.44 (0.20–0.91) | ***0.033*** |
| PWY-6876: isopropanol biosynthesis (engineered) | 0.75 (0.60–0.92) | ***0.008*** | 0.78 (0.59–0.99) | 0.050 | 0.77 (0.59–0.98) | ***0.047*** |
| PWY-7371: 1,4-dihydroxy-6-naphthoate biosynthesis II | 0.79 (0.67–0.92) | ***0.003*** | 0.84 (0.70–1.01) | 0.073 | 0.84 (0.69–1.01) | 0.069 |
| PWY-7992: superpathway of menaquinol-8 biosynthesis III | 0.81 (0.70–0.93) | ***0.004*** | 0.86 (0.73–1.01) | 0.074 | 0.86 (0.72–1.01) | 0.070 |
| PWY-7013: (S)-propane-1,2-diol degradation | 1.36 (0.81–2.35) | 0.261 | 1.62 (0.90–2.97) | 0.111 | 1.66 (0.91–3.08) | 0.099 |
| PWY-5920: superpathway of heme b biosynthesis from glycine | 1.20 (1.06–1.38) | ***0.007*** | 1.12 (0.95–1.32) | 0.188 | 1.12 (0.95–1.33) | 0.170 |
| METHGLYUT-PWY: superpathway of methylglyoxal degradation | 0.87 (0.66–1.12) | 0.291 | 0.84 (0.60–1.12) | 0.249 | 0.84 (0.60–1.13) | 0.266 |
| PWY-6588: pyruvate fermentation to acetone | 1.16 (0.73–1.86) | 0.525 | 1.36 (0.79–2.40) | 0.276 | 1.36 (0.79–2.40) | 0.276 |
| PWY-5973: cis-vaccenate biosynthesis | 2.14 (0.58–8.26) | 0.260 | 1.51 (0.32–7.25) | 0.603 | 1.49 (0.31–7.20) | 0.613 |
| RIBOSYN2-PWY: flavin biosynthesis I (bacteria and plants) | 0.27 (0.03–1.95) | 0.203 | 0.68 (0.06–7.33) | 0.752 | 0.61 (0.05–6.97) | 0.691 |
| PWY-7883: anhydromuropeptides recycling II | 0.96 (0.80–1.16) | 0.701 | 0.98 (0.79–1.22) | 0.877 | 0.98 (0.79–1.22) | 0.868 |
| PWY-6901: superpathway of glucose and xylose degradation | 2.36 (0.58–10.42) | 0.240 | 0.91 (0.15–4.88) | 0.914 | 0.95 (0.15–5.41) | 0.958 |
| **Enzyme** | | | | | | |
| EC 1.8.98.1 (Dihydromethanophenazine:CoB–CoM heterodisulfide reductase) | 0.48 (0.32–0.68) | ***0.000*** | 0.50 (0.31–0.77) | ***0.002*** | 0.49 (0.30–0.76) | ***0.002*** |
| EC 2.7.1.113 (Deoxyguanosine kinase) | 0.75 (0.63–0.87) | ***0.000*** | 0.75 (0.61–0.90) | ***0.004*** | 0.75 (0.61–0.90) | ***0.003*** |
| EC 3.4.13.21 (Dipeptidase E) | 0.39 (0.21–0.66) | ***0.001*** | 0.38 (0.18–0.72) | ***0.006*** | 0.37 (0.18–0.7) | ***0.005*** |
| EC 1.2.7.12 (Formylmethanofuran dehydrogenase, previously 1.2.99.5) | 0.81 (0.71–0.92) | ***0.001*** | 0.80 (0.68–0.94) | ***0.008*** | 0.80 (0.68–0.94) | ***0.008*** |
| EC 3.5.1.81 (N-acyl-D-amino-acid deacylase) | 1.34 (1.11–1.67) | ***0.004*** | 1.37 (1.09–1.75) | ***0.009*** | 1.38 (1.10–1.79) | ***0.008*** |
| EC 2.4.1.319 (Beta-1,4-mannooligosaccharide phosphorylase) | 0.79 (0.67–0.91) | ***0.002*** | 0.81 (0.67–0.98) | ***0.027*** | 0.81 (0.67–0.97) | ***0.026*** |
| EC 4.2.1.115 (UDP-N-acetylglucosamine 4,6-dehydratase (inverting)) | 1.18 (1.02–1.38) | ***0.030*** | 1.22 (1.02–1.47) | ***0.032*** | 1.23 (1.03–1.49) | ***0.028*** |
| EC 3.4.22.8 (Clostripain) | 0.67 (0.46–0.94) | ***0.030*** | 0.63 (0.40–0.94) | ***0.036*** | 0.63 (0.40–0.95) | ***0.036*** |
| EC 2.4.1.329 (Sucrose 6(F)-phosphate phosphorylase) | 1.38 (1.06–1.86) | ***0.024*** | 1.37 (1.02–1.92) | ***0.047*** | 1.40 (1.03–1.97) | ***0.042*** |
| EC 7.1.1.6 (plastoquinol–plastocyanin reductase, previously 1.10.9.1) | 0.74 (0.55–0.95) | ***0.029*** | 0.73 (0.51–0.99) | 0.057 | 0.72 (0.50–0.98) | 0.055 |
| EC 2.1.1.61 (tRNA (5-methylaminomethyl-2-thiouridylate)-methyltransferase) | 0.95 (0.82–1.10) | 0.516 | 0.86 (0.72–1.01) | 0.073 | 0.85 (0.72–1.01) | 0.069 |
| EC 2.5.1.120 (Aminodeoxyfutalosine synthase) | 0.80 (0.68–0.93) | ***0.005*** | 0.85 (0.71–1.02) | 0.078 | 0.85 (0.71–1.01) | 0.072 |
| EC 5.1.3.8 (N-acylglucosamine 2-epimerase) | 0.76 (0.48–1.17) | 0.217 | 0.62 (0.35–1.08) | 0.101 | 0.60 (0.32–1.06) | 0.088 |
| EC 1.4.99.6 (D-arginine dehydrogenase) | 1.25 (1.08–1.46) | ***0.003*** | 1.11 (0.94–1.32) | 0.220 | 1.12 (0.94–1.33) | 0.203 |
| EC 6.3.4.15 (Biotin–[biotin carboxyl-carrier protein] ligase) | 0.20 (0.03–1.09) | 0.068 | 0.49 (0.06–3.76) | 0.487 | 0.46 (0.05–3.77) | 0.468 |
| EC 2.1.1.173 (23S rRNA (guanine(2445)-N(2))-methyltransferase) | 0.88 (0.24–3.10) | 0.835 | 1.27 (0.27–5.58) | 0.756 | 1.30 (0.28–5.88) | 0.734 |

**Supplementary Table 5**. Logistic regression results for plasma metabolites (n=182) for cases compared to controls for the top 50 metabolites.

| Metabolite (n=182) | n outliers removed | OPLS-DA | Model 0 (Unadjusted) n=138 / AIC 183.8 | | | Model 1 (Confounder Set) n=134 / AIC 169.4 | | | Model 2 (Diet Quality) n=134 / AIC 174.9 | | |
|---|---|---|---|---|---|---|---|---|---|---|---|
| | | VIP Score | OR (95% CI) | p | q | OR (95% CI) | p | q | OR (95% CI) | p | q |
| Genistein | 0 | ***2.56*** | 1.35 (1.13–1.65) | ***0.002*** | ***0.212*** | 1.46 (1.17–1.87) | ***0.001*** | ***0.133*** | 1.49 (1.18–1.93) | ***0.001*** | ***0.143*** |
| Hypoxanthine | 2 | 0.99 | 0.40 (0.20–0.73) | ***0.005*** | ***0.212*** | 0.28 (0.12–0.59) | ***0.001*** | ***0.133*** | 0.28 (0.12–0.60) | ***0.002*** | ***0.143*** |
| Daidzein | 0 | ***2.32*** | 1.10 (1.02–1.22) | 0.032 | 0.294 | 1.15 (1.06–1.29) | ***0.003*** | ***0.158*** | 1.16 (1.06–1.29) | ***0.003*** | ***0.159*** |
| Matairesinol | 2 | ***2.06*** | 0.55 (0.35–0.80) | ***0.004*** | ***0.212*** | 0.48 (0.28–0.77) | ***0.004*** | ***0.161*** | 0.48 (0.28–0.76) | ***0.004*** | ***0.159*** |
| Glutaryl-carnitine | 0 | 0.73 | 2.05 (1.06–4.09) | 0.036 | 0.311 | 3.30 (1.48–7.92) | ***0.005*** | ***0.183*** | 3.33 (1.49–8.03) | ***0.005*** | ***0.178*** |
| Methionine | 1 | 0.61 | 0.39 (0.15–0.97) | 0.049 | 0.316 | 0.21 (0.06–0.65) | ***0.009*** | ***0.231*** | 0.21 (0.06–0.65) | ***0.009*** | ***0.238*** |
| Hyocholic acid 3-sulfate | 5 | ***1.76*** | 1.45 (1.04–2.06) | 0.032 | 0.294 | 1.85 (1.19–3.01) | ***0.009*** | ***0.231*** | 1.84 (1.19–3.01) | ***0.009*** | ***0.238*** |
| Murocholic acid | 0 | ***2.75*** | 1.37 (1.12–1.69) | ***0.003*** | ***0.212*** | 1.37 (1.08–1.76) | ***0.011*** | ***0.244*** | 1.37 (1.08–1.76) | ***0.011*** | ***0.239*** |
| 3-Methylglutaryl-carnitine | 1 | ***1.72*** | 2.01 (1.20–3.51) | ***0.010*** | ***0.233*** | 2.26 (1.20–4.45) | ***0.014*** | ***0.244*** | 2.30 (1.21–4.63) | ***0.014*** | ***0.245*** |
| Estriol (E3) | 0 | ***1.63*** | 1.06 (1.00–1.12) | 0.048 | 0.316 | 1.09 (1.02–1.18) | ***0.014*** | ***0.244*** | 1.09 (1.02–1.19) | ***0.015*** | ***0.245*** |
| Pinoresinol | 5 | 1.38 | 0.70 (0.51–0.92) | 0.017 | 0.280 | 0.64 (0.44–0.90) | ***0.015*** | ***0.244*** | 0.64 (0.43–0.90) | ***0.015*** | ***0.245*** |
| DHEA-sulfate | 0 | ***1.55*** | 1.37 (0.99–1.92) | 0.061 | 0.339 | 1.72 (1.12–2.75) | ***0.016*** | ***0.249*** | 1.72 (1.12–2.75) | ***0.017*** | ***0.245*** |
| 16a-Hydroxyestrone | 2 | ***1.77*** | 0.44 (0.21–0.87) | 0.024 | 0.280 | 0.39 (0.17–0.82) | 0.018 | 0.251 | 0.37 (0.16–0.80) | ***0.016*** | ***0.245*** |
| Glycoursodeoxycholic acid | 0 | ***2.42*** | 1.32 (1.08–1.63) | ***0.009*** | ***0.233*** | 1.33 (1.05–1.71) | 0.020 | 0.251 | 1.33 (1.06–1.72) | 0.019 | 0.251 |
| Indoxyl sulfate | 1 | 0.92 | 0.70 (0.45–1.06) | 0.094 | 0.396 | 0.55 (0.32–0.90) | 0.021 | 0.251 | 0.55 (0.32–0.90) | 0.022 | 0.265 |
| Tryptophan | 0 | 0.72 | 0.23 (0.06–0.78) | 0.022 | 0.280 | 0.19 (0.04–0.78) | 0.026 | 0.275 | 0.20 (0.04–0.80) | 0.027 | 0.265 |
| Valeric acid | 1 | 0.84 | 0.46 (0.23–0.87) | 0.021 | 0.280 | 0.41 (0.18–0.88) | 0.028 | 0.275 | 0.40 (0.17–0.87) | 0.027 | 0.265 |
| Taurochenodexycholic acid | 0 | 1.17 | 0.91 (0.79–1.03) | 0.143 | 0.490 | 0.83 (0.7–0.97) | 0.028 | 0.275 | 0.83 (0.69–0.97) | 0.029 | 0.265 |
| Caproic acid | 0 | ***1.60*** | 0.19 (0.06–0.58) | ***0.006*** | ***0.212*** | 0.2 (0.04–0.77) | 0.029 | 0.275 | 0.19 (0.04–0.75) | 0.027 | 0.265 |
| Taurodeoxycholic acid 3-sulfate | 0 | 0.98 | 0.86 (0.72–0.99) | 0.056 | 0.327 | 0.82 (0.68–0.97) | 0.032 | 0.288 | 0.82 (0.67–0.97) | 0.029 | 0.265 |
| Taurolithocholic acid | 0 | ***1.99*** | 0.82 (0.68–0.97) | 0.026 | 0.280 | 0.79 (0.63–0.98) | 0.034 | 0.293 | 0.79 (0.63–0.98) | 0.035 | 0.279 |
| Butyric acid | 0 | 0.84 | 0.65 (0.41–1.00) | 0.056 | 0.327 | 0.57 (0.32–0.95) | 0.037 | 0.302 | 0.56 (0.32–0.96) | 0.039 | 0.279 |
| Aldosterone | 1 | 1.06 | 0.65 (0.37–1.12) | 0.124 | 0.467 | 0.5 (0.26–0.95) | 0.040 | 0.302 | 0.5 (0.25–0.94) | 0.036 | 0.279 |
| O-Desmethylangolensin | 0 | 1.29 | 1.10 (0.94–1.28) | 0.236 | 0.587 | 1.2 (1.01–1.45) | 0.043 | 0.302 | 1.2 (1.01–1.45) | 0.046 | 0.279 |
| Chenodeoxycholic acid 3-sulfate | 3 | 0.76 | 0.90 (0.73–1.10) | 0.293 | 0.641 | 0.78 (0.61–0.99) | 0.043 | 0.302 | 0.78 (0.61–0.98) | 0.040 | 0.279 |
| Allocholic acid | 0 | 0.49 | 0.96 (0.89–1.04) | 0.297 | 0.641 | 0.91 (0.82–1) | 0.045 | 0.302 | 0.9 (0.81–0.99) | 0.042 | 0.279 |
| Testosterone | 0 | 1.40 | 1.46 (0.89–2.46) | 0.140 | 0.490 | 1.8 (1.02–3.31) | 0.048 | 0.302 | 1.83 (1.03–3.4) | 0.045 | 0.279 |
| Biliverdin | 1 | 0.64 | 0.66 (0.43–1.00) | 0.056 | 0.327 | 0.62 (0.37–0.98) | 0.050 | 0.302 | 0.6 (0.36–0.97) | 0.042 | 0.279 |
| Ursodeoxycholic acid 7-sulfate | 3 | ***1.79*** | 1.28 (1.04–1.60) | 0.026 | 0.280 | 1.28 (1.01–1.66) | 0.050 | 0.302 | 1.28 (1–1.65) | 0.057 | 0.304 |
| LysoPE(20:4) | 0 | 1.33 | 1.65 (0.78–3.63) | 0.199 | 0.539 | 2.6 (1.02–7.05) | 0.051 | 0.302 | 2.68 (1.04–7.35) | 0.046 | 0.279 |
| Sphingosine-1P (d18:1) | 0 | 0.08 | 0.66 (0.46–0.89) | ***0.010*** | ***0.233*** | 0.71 (0.48–0.99) | 0.053 | 0.302 | 0.7 (0.48–0.98) | 0.052 | 0.298 |
| Androsterone 3-sulfate | 0 | 1.49 | 1.31 (0.99–1.76) | 0.067 | 0.341 | 1.4 (1–2.02) | 0.054 | 0.302 | 1.4 (1–2.01) | 0.058 | 0.304 |
| Acetic acid | 1 | 1.18 | 2.03 (1.03–4.15) | 0.045 | 0.316 | 2.16 (1–4.91) | 0.056 | 0.302 | 2.31 (1.03–5.43) | 0.046 | 0.279 |
| Phenylalanine | 0 | 0.23 | 0.49 (0.15–1.52) | 0.223 | 0.564 | 0.25 (0.06–1) | 0.056 | 0.302 | 0.25 (0.06–1.03) | 0.061 | 0.309 |
| Adipoyl-carnitine | 0 | 0.92 | 0.82 (0.56–1.19) | 0.307 | 0.641 | 0.62 (0.37–1.02) | 0.065 | 0.331 | 0.62 (0.36–1.03) | 0.069 | 0.338 |
| Acetyl-carnitine | 0 | ***1.76*** | 2.27 (1.1–4.87) | 0.029 | 0.294 | 2.23 (0.97–5.41) | 0.065 | 0.331 | 2.4 (0.99–6.14) | 0.057 | 0.304 |
| Coumestrol | 0 | 0.98 | 0.97 (0.94–1.00) | 0.069 | 0.341 | 0.97 (0.93–1) | 0.068 | 0.336 | 0.96 (0.93–1) | 0.052 | 0.298 |
| Tyrosine | 0 | 0.22 | 0.56 (0.22–1.38) | 0.210 | 0.539 | 0.36 (0.11–1.06) | 0.070 | 0.337 | 0.35 (0.11–1.07) | 0.072 | 0.338 |
| p-Cresol sulfate | 1 | ***2.08*** | 0.74 (0.53–1.01) | 0.063 | 0.340 | 0.71 (0.48–1.03) | 0.076 | 0.346 | 0.71 (0.47–1.03) | 0.081 | 0.349 |
| Cholic acid | 0 | 0.27 | 0.9 (0.77–1.05) | 0.192 | 0.539 | 0.85 (0.71–1.01) | 0.076 | 0.346 | 0.85 (0.7–1.01) | 0.071 | 0.338 |
| Taurolithocholic acid 3-sulfate | 5 | ***1.70*** | 0.77 (0.6–0.98) | 0.038 | 0.311 | 0.78 (0.58–1.02) | 0.078 | 0.347 | 0.78 (0.58–1.02) | 0.084 | 0.351 |
| Dehydrocholic acid | 0 | 1.27 | 1.12 (0.97–1.32) | 0.142 | 0.490 | 1.15 (0.98–1.38) | 0.095 | 0.412 | 1.16 (0.99–1.39) | 0.077 | 0.349 |
| LysoPE(18:2) | 0 | 0.88 | 1.17 (0.76–1.81) | 0.483 | 0.727 | 1.59 (0.93–2.82) | 0.098 | 0.414 | 1.66 (0.96–3.01) | 0.080 | 0.349 |
| Cortisol | 0 | ***1.67*** | 0.50 (0.24–1.01) | 0.060 | 0.339 | 0.49 (0.21–1.13) | 0.101 | 0.417 | 0.47 (0.19–1.09) | 0.085 | 0.351 |
| Androstenedione | 0 | 0.62 | 1.28 (0.8–2.05) | 0.303 | 0.641 | 1.58 (0.92–2.78) | 0.104 | 0.420 | 1.61 (0.93–2.88) | 0.097 | 0.377 |
| DHEA | 0 | 0.56 | 1.12 (0.79–1.6) | 0.525 | 0.741 | 1.41 (0.93–2.17) | 0.108 | 0.429 | 1.42 (0.94–2.2) | 0.102 | 0.386 |
| Tauroursodeoxycholic acid 3-sulfate | 0 | ***2.00*** | 1.09 (1.00–1.19) | 0.048 | 0.316 | 1.09 (0.98–1.21) | 0.111 | 0.429 | 1.09 (0.98–1.21) | 0.108 | 0.401 |
| L-Carnitine | 0 | 0.20 | 0.87 (0.29–2.54) | 0.795 | 0.889 | 0.33 (0.08–1.27) | 0.113 | 0.430 | 0.29 (0.07–1.19) | 0.092 | 0.365 |
| Corticosterone | 0 | 1.47 | 0.74 (0.52–1.04) | 0.092 | 0.396 | 0.73 (0.49–1.07) | 0.117 | 0.433 | 0.7 (0.45–1.05) | 0.088 | 0.356 |
| Glycitein | 5 | 0.53 | 1.21 (0.97–1.53) | 0.097 | 0.402 | 1.23 (0.95–1.62) | 0.122 | 0.435 | 1.22 (0.94–1.62) | 0.141 | 0.435 |

**Supplementary Table 6**. Logistic regression results for stool metabolites (n=185) for cases compared to controls for the top 50 metabolites.

| Metabolite (n=185) | n outliers removed | PLS-DA | Model 0 (Unadjusted) (n=139) / AIC 174.0 | | | Model 1 (Confounder Set) n=135 / AIC 165.2 | | | Model 2 (Diet Quality) n=135 / AIC 170.6 | | |
|---|---|---|---|---|---|---|---|---|---|---|---|
| | | VIP Scores | OR (95% CI) | p | q | OR (95% CI) | p | q | OR (95% CI) | p | q |
| Adipoyl-carnitine | 0 | ***2.45*** | 1.14 (1.07–1.22) | ***7.62E-05*** | ***0.005*** | 1.26 (1.15–1.40) | ***4.51E-06*** | ***0.001*** | 1.27 (1.16–1.42) | ***3.59E-06*** | ***0.001*** |
| DHEA-sulfate | 0 | ***1.54*** | 1.36 (1.17–1.59) | ***6.37E-05*** | ***0.005*** | 1.38 (1.16–1.68) | ***4.94E-04*** | ***0.033*** | 1.39 (1.17–1.69) | ***4.47E-04*** | ***0.029*** |
| Genistein | 0 | ***2.30*** | 1.38 (1.20–1.62) | ***3.30E-05*** | ***0.005*** | 1.36 (1.15–1.64) | ***0.001*** | ***0.033*** | 1.37 (1.16–1.65) | ***4.70E-04*** | ***0.029*** |
| Oleoyl-carnitine | 0 | 1.32 | 1.43 (1.20–1.72) | ***1.05E-04*** | ***0.005*** | 1.39 (1.13–1.75) | ***0.003*** | ***0.140*** | 1.40 (1.13–1.77) | ***0.003*** | ***0.141*** |
| 3-Methylcrotonyl-L-carnitine | 0 | 1.37 | 1.16 (1.04–1.31) | ***0.010*** | ***0.059*** | 1.26 (1.09–1.49) | ***0.004*** | ***0.140*** | 1.26 (1.09–1.49) | ***0.004*** | ***0.141*** |
| 7-Ketodeoxycholic acid | 0 | 1.14 | 1.21 (1.09–1.40) | ***0.002*** | ***0.022*** | 1.21 (1.07–1.40) | ***0.005*** | ***0.169*** | 1.21 (1.07–1.41) | ***0.005*** | ***0.163*** |
| Butyryl-carnitine | 1 | 1.27 | 1.71 (1.28–2.38) | ***0.001*** | ***0.014*** | 1.63 (1.16–2.38) | ***0.007*** | ***0.175*** | 1.64 (1.17–2.42) | ***0.007*** | ***0.166*** |
| Ursodeoxycholic acid 7-sulfate | 0 | 1.20 | 1.10 (1.04–1.16) | ***0.001*** | ***0.014*** | 1.09 (1.02–1.16) | ***0.008*** | ***0.175*** | 1.09 (1.03–1.16) | ***0.007*** | ***0.166*** |
| Palmitoyl-carnitine | 0 | 1.15 | 1.38 (1.17–1.67) | ***0.000*** | ***0.011*** | 1.32 (1.07–1.65) | ***0.012*** | ***0.208*** | 1.32 (1.07–1.66) | ***0.013*** | ***0.200*** |
| Androsterone 3-sulfate | 0 | 0.81 | 1.10 (1.02–1.20) | ***0.014*** | ***0.066*** | 1.13 (1.02–1.25) | ***0.018*** | ***0.208*** | 1.13 (1.03–1.26) | ***0.017*** | ***0.200*** |
| Glutaryl-carnitine | 0 | 0.87 | 1.08 (1.02–1.15) | ***0.014*** | ***0.066*** | 1.10 (1.02–1.19) | ***0.019*** | ***0.208*** | 1.10 (1.02–1.19) | ***0.019*** | ***0.200*** |
| Daidzein | 0 | 0.71 | 1.21 (1.02–1.45) | ***0.031*** | ***0.092*** | 1.30 (1.05–1.63) | ***0.019*** | ***0.208*** | 1.30 (1.05–1.65) | ***0.020*** | ***0.200*** |
| Glycitein | 0 | 0.63 | 1.17 (0.95–1.44) | 0.149 | 0.242 | 1.37 (1.06–1.81) | ***0.020*** | ***0.208*** | 1.37 (1.06–1.82) | ***0.019*** | ***0.200*** |
| Linoleyl-carnitine | 0 | 1.11 | 1.40 (1.16–1.72) | ***0.001*** | ***0.014*** | 1.30 (1.05–1.64) | ***0.020*** | ***0.208*** | 1.30 (1.05–1.65) | ***0.020*** | ***0.200*** |
| Octanoyl-carnitine | 0 | ***2.35*** | 0.8 (0.63–1.01) | 0.070 | 0.144 | 0.71 (0.53–0.94) | ***0.020*** | ***0.208*** | 0.70 (0.51–0.93) | ***0.017*** | ***0.200*** |
| Lithocholic acid 3-sulfate | 0 | 1.07 | 1.17 (1.07–1.3) | ***0.001*** | ***0.017*** | 1.13 (1.02–1.26) | ***0.023*** | ***0.208*** | 1.13 (1.02–1.26) | ***0.022*** | ***0.200*** |
| Arachidonoyl-carnitine | 0 | 1.35 | 1.22 (1.08–1.43) | ***0.005*** | ***0.042*** | 1.18 (1.03–1.39) | ***0.023*** | ***0.208*** | 1.18 (1.03–1.39) | ***0.022*** | ***0.200*** |
| Cholic acid 3-sulfate | 0 | ***1.66*** | 1.08 (1.04–1.13) | ***0.000*** | ***0.011*** | 1.06 (1.01–1.12) | ***0.023*** | ***0.208*** | 1.06 (1.01–1.12) | ***0.025*** | ***0.200*** |
| Stearoyl-carnitine | 0 | 1.05 | 1.44 (1.17–1.8) | ***0.001*** | ***0.015*** | 1.38 (1.05–1.85) | ***0.024*** | ***0.208*** | 1.42 (1.06–1.93) | ***0.021*** | ***0.200*** |
| Deoxycholic acid 3-sulfate | 0 | 1.13 | 1.14 (1.06–1.25) | ***0.002*** | ***0.022*** | 1.12 (1.02–1.23) | ***0.025*** | ***0.208*** | 1.12 (1.02–1.24) | ***0.023*** | ***0.200*** |
| Hypoxanthine | 0 | 1.06 | 1.82 (1.26–2.74) | ***0.002*** | ***0.026*** | 1.66 (1.08–2.65) | ***0.027*** | ***0.208*** | 1.66 (1.08–2.67) | ***0.027*** | ***0.200*** |
| Dioxolithocholic acid | 0 | 0.81 | 1.3 (1.07–1.59) | ***0.009*** | ***0.059*** | 1.31 (1.04–1.69) | ***0.027*** | ***0.208*** | 1.33 (1.05–1.74) | ***0.026*** | ***0.200*** |
| Lithocholic acid | 2 | 0.33 | 2.18 (1.28–3.88) | ***0.006*** | ***0.045*** | 2.07 (1.1–4.05) | ***0.027*** | ***0.208*** | 2.09 (1.11–4.14) | ***0.027*** | ***0.200*** |
| Hyodeoxycholic acid | 0 | 1.25 | 0.83 (0.66–1.03) | 0.107 | 0.195 | 0.75 (0.57–0.96) | ***0.028*** | ***0.208*** | 0.74 (0.57–0.96) | ***0.027*** | ***0.200*** |
| Caproic acid | 0 | ***1.79*** | 0.81 (0.7–0.93) | ***0.004*** | ***0.041*** | 0.83 (0.7–0.98) | ***0.028*** | ***0.208*** | 0.83 (0.7–0.98) | ***0.029*** | ***0.204*** |
| 12-Ketochenodeoxycholic acid | 0 | 0.68 | 1.07 (1.01–1.15) | ***0.030*** | ***0.092*** | 1.09 (1.01–1.18) | ***0.029*** | ***0.208*** | 1.1 (1.01–1.2) | ***0.024*** | ***0.200*** |
| Ursodeoxycholic acid 3-sulfate | 0 | 0.84 | 1.06 (1.02–1.1) | ***0.009*** | ***0.059*** | 1.06 (1.01–1.11) | ***0.030*** | ***0.208*** | 1.06 (1.01–1.11) | ***0.031*** | ***0.205*** |
| Dehydrocholic acid | 0 | 0.58 | 1.05 (1–1.11) | 0.072 | 0.144 | 1.07 (1.01–1.15) | ***0.034*** | ***0.221*** | 1.08 (1.01–1.15) | ***0.030*** | ***0.205*** |
| Glycoursodeoxycholic acid | 3 | 0.88 | 1.32 (1.12–1.59) | ***0.002*** | ***0.022*** | 1.23 (1.01–1.52) | 0.044 | 0.274 | 1.24 (1.02–1.54) | ***0.039*** | ***0.248*** |
| p-Cresol sulfate | 0 | 0.71 | 1.2 (1.03–1.43) | ***0.024*** | ***0.091*** | 1.22 (1.01–1.49) | 0.045 | 0.274 | 1.23 (1.01–1.51) | 0.043 | 0.263 |
| Tryptophan | 0 | 1.00 | 1.62 (1.22–2.21) | ***0.001*** | ***0.020*** | 1.42 (1.01–2.02) | 0.046 | 0.274 | 1.41 (1.01–2.02) | 0.049 | 0.292 |
| Succinyl-carnitine | 0 | 0.65 | 1.27 (1.02–1.59) | ***0.033*** | ***0.095*** | 1.3 (1–1.72) | 0.055 | 0.301 | 1.31 (1–1.75) | 0.055 | 0.298 |
| L-Carnitine | 0 | 0.84 | 1.35 (1.06–1.75) | ***0.019*** | ***0.075*** | 1.35 (1–1.86) | 0.055 | 0.301 | 1.36 (1–1.9) | 0.056 | 0.298 |
| Glycochenodeoxycholic acid 3-sulfate | 0 | 0.94 | 1.11 (1.04–1.18) | ***0.003*** | ***0.027*** | 1.08 (1–1.17) | 0.058 | 0.301 | 1.08 (1–1.17) | 0.058 | 0.298 |
| Matairesinol | 0 | 0.57 | 1.2 (0.99–1.46) | 0.063 | 0.138 | 1.24 (1–1.57) | 0.061 | 0.301 | 1.24 (1–1.58) | 0.062 | 0.298 |
| LysoPC(16:1) | 1 | 0.92 | 1.33 (1.09–1.67) | ***0.008*** | ***0.059*** | 1.26 (0.99–1.63) | 0.063 | 0.301 | 1.27 (0.99–1.64) | 0.063 | 0.298 |
| Cholesterol | 0 | 0.94 | 1.26 (1.02–1.55) | ***0.031*** | ***0.092*** | 1.28 (0.99–1.68) | 0.064 | 0.301 | 1.31 (1–1.76) | 0.055 | 0.298 |
| Cortisol | 0 | 1.05 | 1.05 (1–1.11) | 0.054 | 0.124 | 1.06 (1–1.14) | 0.064 | 0.301 | 1.07 (1–1.14) | 0.060 | 0.298 |
| beta-Muricholic acid | 4 | 0.69 | 1.28 (1.03–1.64) | ***0.034*** | ***0.097*** | 1.3 (1–1.75) | 0.067 | 0.301 | 1.32 (1–1.8) | 0.061 | 0.298 |
| Isolithocholic acid 3-sulfate | 0 | 1.32 | 1.16 (1.07–1.27) | ***0.001*** | ***0.019*** | 1.1 (1–1.23) | 0.071 | 0.301 | 1.1 (1–1.23) | 0.075 | 0.298 |
| Pinoresinol | 0 | 0.79 | 1.32 (1.07–1.65) | ***0.011*** | ***0.059*** | 1.25 (0.99–1.61) | 0.073 | 0.301 | 1.25 (0.99–1.61) | 0.074 | 0.298 |
| Norursodeoxycholic acid | 0 | 1.05 | 1.09 (1.02–1.18) | ***0.019*** | ***0.075*** | 1.08 (1–1.18) | 0.073 | 0.301 | 1.08 (1–1.18) | 0.071 | 0.298 |
| Alloisolithocholic acid | 4 | ***1.65*** | 0.87 (0.74–1.01) | 0.067 | 0.144 | 0.84 (0.68–1.02) | 0.076 | 0.301 | 0.84 (0.69–1.02) | 0.079 | 0.298 |
| alfa-Muricholic acid | 0 | ***1.68*** | 0.97 (0.91–1.02) | 0.248 | 0.345 | 0.94 (0.87–1.01) | 0.078 | 0.301 | 0.94 (0.87–1) | 0.071 | 0.298 |
| Acetyl-carnitine | 2 | 0.97 | 1.43 (1.1–1.9) | ***0.010*** | ***0.059*** | 1.34 (0.98–1.89) | 0.078 | 0.301 | 1.34 (0.98–1.89) | 0.078 | 0.298 |
| Chenodeoxycholic acid 3-sulfate | 0 | 0.86 | 1.06 (1.01–1.1) | ***0.009*** | ***0.059*** | 1.05 (1–1.1) | 0.078 | 0.301 | 1.05 (1–1.1) | 0.070 | 0.298 |
| Sphinganine-1P (d18:0) | 0 | 0.40 | 1.23 (1.02–1.49) | ***0.030*** | ***0.092*** | 1.23 (0.98–1.57) | 0.080 | 0.301 | 1.23 (0.98–1.57) | 0.081 | 0.298 |
| Norcholic acid | 5 | ***1.53*** | 1.29 (1.07–1.59) | ***0.011*** | ***0.062*** | 1.25 (0.98–1.62) | 0.080 | 0.301 | 1.26 (0.98–1.64) | 0.082 | 0.298 |
| 3-Methylglutaryl-carnitine | 0 | 0.44 | 1.04 (0.96–1.14) | 0.348 | 0.450 | 1.1 (0.99–1.25) | 0.081 | 0.301 | 1.1 (0.99–1.25) | 0.084 | 0.298 |
| 4-Methoxyestrone | 0 | 1.40 | 1.08 (1–1.17) | 0.071 | 0.144 | 1.09 (0.99–1.2) | 0.081 | 0.301 | 1.09 (0.99–1.21) | 0.073 | 0.298 |

**Supplementary Table 7**. DIABLO loading values for the model fit to the top selected metagenomic features (species, pathways, enzymes) and metabolites (plasma, stool).

| Species | | | |
|---|---|---|---|
| Component 1 | | Component 2 | |
| Top Featurs | loading | Top Features | loading |
| Anaerotruncus rubiinfantis | 0.45 | Anaerobutyricum hallii | 0.51 |
| Alistipes putredinis | 0.36 | GGB3057 SGB4059 | -0.43 |
| C. Avimonas narfia | 0.29 | Mediterraneibacter butyricigenes | 0.32 |
| Alistipes finegoldii | 0.27 | Sutterella wadsworthensis | -0.28 |
| Anaerobutyricum hallii | 0.27 | C. Avimonas narfia | -0.27 |
| Blautia obeum | 0.27 | Alistipes finegoldii | -0.27 |
| Alistipes shahii | 0.26 | C. Cibionibacter quicibialis | 0.24 |
| Mediterraneibacter butyricigenes | 0.26 | Anaerotruncus rubiinfantis | -0.23 |
| L. bacterium AM48-27BH | 0.24 | L. bacterium AM48-27BH | 0.23 |
| Blautia SGB4815 | 0.21 | Blautia obeum | -0.14 |
| C. Cibionibacter quicibialis | 0.20 | Clostridium fessum | -0.12 |
| Faecalibacillus intestinalis | 0.17 | Alistipes putredinis | 0.11 |
| GGB3057 SGB4059 | 0.14 | Alistipes shahii | 0.10 |
| Clostridium fessum | 0.12 | Blautia SGB4815 | 0.04 |
| Mediterraneibacter massiliensis | 0.11 | Mediterraneibacter massiliensis | 0.01 |
| Sutterella wadsworthensis | 0.10 | Faecalibacillus intestinalis | 0.00 |

| Pathways | | | |
|---|---|---|---|
| Component 1 | | Component 2 | |
| Top Featurs | loading | Top Features | loading |
| PWY-7761 | 0.40 | METHGLYUT-PWY | -0.50 |
| PWY-7992 | 0.37 | PWY-6901 | -0.46 |
| P42-PWY | 0.36 | PWY-5973 | -0.43 |
| PWY-7371 | 0.36 | PWY-5920 | -0.31 |
| PWY-5130 | 0.34 | P42-PWY | -0.25 |
| PWY-5973 | -0.30 | RIBOSYN2-PWY | 0.22 |
| PWY-6876 | 0.29 | PWY-7883 | -0.20 |
| PWY-6901 | -0.23 | PWY-5130 | -0.17 |
| PWY-6588 | 0.21 | PWY-7371 | -0.13 |
| PWY-5920 | -0.21 | PWY-7013 | 0.13 |
| RIBOSYN2-PWY | -0.08 | PWY-7992 | -0.12 |
| PWY-8131 | 0.03 | PWY-6876 | -0.12 |
| PWY-7883 | -0.02 | PWY-8131 | 0.11 |
| PWY-7013 | 0.02 | PWY-7761 | -0.03 |
| METHGLYUT-PWY | 0.01 | PWY-6588 | 0.01 |

| Enzymes | | | |
|---|---|---|---|
| Component 1 | | Component 2 | |
| Top Features | loading | Top Features | loading |
| EC 1.8.98.1 | 0.50 | EC 3.4.22.8 | -0.39 |
| EC 1.2.7.12 | 0.46 | EC 4.2.1.115 | 0.37 |
| EC 2.5.1.120 | 0.34 | EC 6.3.4.15 | 0.37 |
| EC 2.4.1.319 | 0.30 | EC 2.1.1.61 | -0.36 |
| EC 3.4.13.21 | 0.30 | EC 5.1.3.8 | -0.31 |
| EC 2.7.1.113 | 0.25 | EC 7.1.1.6 | -0.29 |
| EC 2.1.1.173 | -0.24 | EC 1.4.99.6 | -0.22 |
| EC 7.1.1.6 | 0.21 | EC 2.7.1.113 | -0.21 |
| EC 2.4.1.329 | -0.17 | EC 2.4.1.329 | 0.21 |
| EC 1.4.99.6 | -0.14 | EC 2.1.1.173 | -0.19 |
| EC 5.1.3.8 | 0.13 | EC 3.4.13.21 | -0.16 |
| EC 6.3.4.15 | 0.10 | EC 2.4.1.319 | 0.13 |
| EC 3.5.1.81 | -0.07 | EC 1.8.98.1 | 0.13 |
| EC 2.1.1.61 | -0.04 | EC 2.5.1.120 | -0.11 |
| EC 3.4.22.8 | 0.03 | EC 1.2.7.12 | 0.11 |
| EC 4.2.1.115 | 0.01 | EC 3.5.1.81 | 0.04 |

| Plasma Metabolites | | | |
|---|---|---|---|
| Component 1 | | Component 2 | |
| Top Featurs | loading | Top Features | loading |
| Murocholic acid | -0.58 | Methionine | -0.55 |
| Glycoursodeoxycholic acid | -0.39 | Matairesinol | -0.41 |
| Hexanoyl-carnitine | -0.30 | Hypoxanthine | -0.32 |
| Genistein | -0.29 | Murocholic acid | -0.31 |
| Estriol | -0.21 | Daidzein | 0.24 |
| Hyocholic acid 3-sulfate | -0.20 | Glycoursodeoxycholic acid | -0.22 |
| LysoPC(15:0) | 0.19 | Linoleyl-carnitine | 0.21 |
| DHEA-sulfate | -0.18 | 3-Methylglutaryl-carnitine | -0.19 |
| Daidzein | -0.18 | Glutaryl-carnitine | 0.18 |
| Caproic acid | 0.17 | Genistein | 0.17 |
| Matairesinol | 0.17 | DHEA-sulfate | 0.14 |
| Linoleyl-carnitine | -0.15 | 16a-Hydroxyestrone | -0.14 |
| 16a-Hydroxyestrone | 0.15 | Hexanoyl-carnitine | -0.13 |
| 3-Methylglutaryl-carnitine | -0.12 | Pinoresinol | 0.10 |
| Glutaryl-carnitine | -0.12 | Estriol | 0.09 |
| Pinoresinol | -0.09 | LysoPC(15:0) | -0.02 |
| Hypoxanthine | 0.06 | Hyocholic acid 3-sulfate | 0.02 |
| Methionine | 0.03 | Caproic acid | 0.00 |

| Stool Metabolites | | | |
|---|---|---|---|
| Component 1 | | Component 2 | |
| Top Features | loading | Top Features | loading |
| Oleoyl-carnitine | -0.28 | Daidzein | 0.41 |
| Palmitoyl-carnitine | -0.27 | 3-Methylcrotonyl-L-carnitine | 0.37 |
| DHEA-sulfate | -0.26 | Adipoyl-carnitine | 0.36 |
| Ursodeoxycholic acid 7-sulfate | -0.26 | Genistein | 0.30 |
| Linoleyl-carnitine | -0.25 | DHEA-sulfate | 0.29 |
| Lithocholic acid 3-sulfate | -0.25 | Octanoyl-carnitine | -0.29 |
| Stearoyl-carnitine | -0.24 | Hyodeoxycholic acid | -0.28 |
| Dioxolithocholic acid | -0.24 | Glycoursodeoxycholic acid | -0.19 |
| 7-Ketodeoxycholic acid | -0.24 | Palmitoyl-carnitine | -0.19 |
| Deoxycholic acid 3-sulfate | -0.23 | Glutaryl-carnitine | -0.18 |
| Ursodeoxycholic acid 3-sulfate | -0.23 | Stearoyl-carnitine | -0.17 |
| Glycoursodeoxycholic acid | -0.23 | Ursodeoxycholic acid 3-sulfate | -0.14 |
| Cholic acid 3-sulfate | -0.23 | 12-Ketochenodeoxycholic acid | -0.13 |
| Butyryl-carnitine | -0.20 | Ursodeoxycholic acid 7-sulfate | -0.12 |
| Androsterone 3-sulfate | -0.18 | Oleoyl-carnitine | -0.12 |
| Genistein | -0.18 | Dioxolithocholic acid | -0.11 |
| 12-Ketochenodeoxycholic acid | -0.17 | Butyryl-carnitine | 0.10 |
| Adipoyl-carnitine | -0.16 | Deoxycholic acid 3-sulfate | -0.07 |
| Glutaryl-carnitine | -0.13 | Androsterone 3-sulfate | 0.06 |
| Daidzein | -0.12 | Cholic acid 3-sulfate | 0.05 |
| Hyodeoxycholic acid | 0.08 | 7-Ketodeoxycholic acid | 0.04 |
| 3-Methylcrotonyl-L-carnitine | -0.06 | Linoleyl-carnitine | -0.03 |
| Octanoyl-carnitine | -0.03 | Lithocholic acid 3-sulfate | 0.00 |

**Supplementary Table 2.** Describing the metabolites measured in the sample for blood plasma and stool.

| | | Plasma | | | | Stool | | | |
|---|---|---|---|---|---|---|---|---|---|
| Class | Metabolite | Non-zero values (case/con) | Cases mean (SD) | Controls mean (SD) | Units | Non-zero values (case/con) | Cases mean (SD) | Controls mean (SD) | Units |
| Estrogens | Estradiol (E2) | 49 / 45 | 63.34 (96.33) | 85.29 (185.84) | pM | 69 / 70 | 0.6 (0.77) | 0.52 (0.85) | nmol/g |
| Estrogens | 17a-Estradiol | | *Not found* | | | 0 / 0 | | | nmol/g |
| Estrogens | 16-Ketoestradiol | 66 / 67 | 71.77 (82.38) | 138.15 (271.45) | pM | 18 / 23 | 0.02 (0.1) | 0.01 (0.03) | nmol/g |
| Estrogens | 2-Hydroxyestradiol | 58 / 62 | 28.51 (44.41) | 34.71 (51.64) | pM | 69 / 70 | 0.54 (1.07) | 0.47 (0.91) | nmol/g |
| Estrogens | 2-Methoxyestradiol | 0 / 0 | | | pM | 28 / 38 | 0.01 (0.02) | 0.04 (0.06) | nmol/g |
| Estrogens | 4-Hydroxyestradiol | 0 / 0 | | | pM | | *Not found* | | |
| Estrogens | 4-Methoxyestradiol | 22 / 21 | 38.51 (85.4) | 81.86 (193.12) | pM | 50 / 54 | 0.04 (0.07) | 0.04 (0.07) | nmol/g |
| Estrogens | Estradiol 17-glucuronide | 3 / 4 | 0.0001 (0.0004) | 0.0001 (0.0005) | µM | 0 / 0 | | | nmol/g |
| Estrogens | Estradiol 3-glucuronide | 0 / 0 | | | µM | 0 / 0 | | | nmol/g |
| Estrogens | Estradiol 3-sulfate | 1 / 2 | 0.00002 (0.0001) | 0.00005 (0.0003) | µM | 0 / 0 | | | nmol/g |
| Estrogens | Estradiol 17-sulfate | | *Not found* | | | 0 / 0 | | | nmol/g |
| Estrogens | Estradiol 3-sulfate, 17-glucuronide | | *Not found* | | | 0 / 0 | | | nmol/g |
| Estrogens | Estrone (E1) | 68 / 70 | 1250.58 (182.02) | 1220.63 (235.15) | pM | 69 / 70 | 11.34 (14.88) | 11.95 (24.85) | nmol/g |
| Estrogens | 16a-Hydroxyestrone | 68 / 70 | 289.17 (93.43) | 420.07 (444.03) | pM | 0 / 0 | | | nmol/g |
| Estrogens | 16-Epiestriol | 0 / 0 | | | pM | 68 / 67 | 0.31 (0.6) | 0.31 (0.92) | nmol/g |
| Estrogens | 17a-Epiestriol | 36 / 33 | 495.1 (1447.3) | 1355.03 (4070.8) | pM | 36 / 30 | 0.2 (0.4) | 0.08 (0.21) | nmol/g |
| Estrogens | 2-Hydroxyestrone | 50 / 49 | 193.4 (427.4) | 152.53 (265.09) | pM | 62 / 64 | 10.4 (38.51) | 5.08 (12.34) | nmol/g |
| Estrogens | 2-Hydroxyestrone-3-methylether | 0 / 0 | | | pM | 26 / 22 | 0.1 (0.2) | 0.15 (0.51) | nmol/g |
| Estrogens | 2-Methoxyestrone | 0 / 0 | | | pM | 0 / 0 | | | nmol/g |
| Estrogens | 4-Hydroxyestrone | 27 / 18 | 43.91 (105.21) | 69.41 (283.18) | pM | 5 / 4 | 0.03 (0.13) | 0.33 (2.73) | nmol/g |
| Estrogens | 4-Methoxyestrone | 0 / 0 | | | pM | 60 / 54 | 0.17 (0.33) | 0.1 (0.17) | nmol/g |
| Estrogens | Estrone 3-glucuronide | 68 / 70 | 0.51 (0.24) | 0.5 (0.2) | µM | 0 / 0 | | | nmol/g |
| Estrogens | Estrone 3-sulfate | 68 / 70 | 0.002 (0.003) | 0.002 (0.002) | µM | 2 / 0 | 0.01 (0.07) | 0 (0) | nmol/g |
| Estrogens | Estriol (E3) | 65 / 59 | 149.5 (501.32) | 86.82 (228.78) | pM | 60 / 54 | 0.3 (0.4) | 0.1 (0.2) | nmol/g |
| Estrogens | Estriol 16-glucuronide | | *Not found* | | | 0 / 0 | | | nmol/g |
| Estrogens | Estriol 17-glucuronide | | *Not found* | | | 0 / 0 | | | nmol/g |
| Estrogens | Estriol 3-glucuronide | | *Not found* | | | 0 / 0 | | | nmol/g |
| Estrogens | Estetrol | 50 / 47 | 215.23 (385.77) | 413.94 (594.33) | pM | 10 / 11 | 0.0519 (0.1512) | 0.1101 (0.4562) | nmol/g |
| Phytoestrogens | Coumestrol | 43 / 54 | 150.52 (590.11) | 196.38 (513.98) | pM | 69 / 70 | 1.6 (3.8) | 0.69 (0.76) | nmol/g |
| Phytoestrogens | Genistein | 68 / 70 | 20841.9 (59060.8) | 6392.6 (8153.7) | pM | 69 / 70 | 48.53 (148.88) | 3.61 (10.89) | nmol/g |
| Phytoestrogens | Glycitein | 64 / 69 | 111.29 (153.32) | 96.34 (163.6) | pM | 69 / 70 | 24.9 (34.43) | 16.45 (28.4) | nmol/g |
| Phytoestrogens | Daidzein | 67 / 63 | 395.6 (1073.66) | 211.35 (314.41) | pM | 69 / 70 | 35.82 (140.4) | 10.82 (25.81) | nmol/g |
| Phytoestrogens | Dihydrodaidzein | 68 / 70 | 1770.63 (2634.13) | 1996.86 (2110.83) | pM | 69 / 70 | 21.92 (48.16) | 22.72 (41.19) | nmol/g |
| Phytoestrogens | Equol | 66 / 70 | 337.59 (1674.53) | 130.24 (229.87) | pM | 69 / 70 | 30.4 (127.77) | 4.07 (9.88) | nmol/g |
| Phytoestrogens | O-Desmethylangolensin | 68 / 70 | 1078.54 (3579.15) | 598.4 (888.89) | pM | 69 / 70 | 25.34 (63.42) | 15.35 (33.25) | nmol/g |
| Phytoestrogens | Pinoresinol | 68 / 65 | 208.18 (248.44) | 439.07 (1325.21) | pM | 69 / 70 | 4.97 (9.22) | 1.97 (2.76) | nmol/g |
| Phytoestrogens | Pinoresinol diglucoside | 0 / 0 | | | µM | 0 / 0 | | | nmol/g |
| Phytoestrogens | Lariciresinol | 66 / 68 | 62.87 (79.39) | 130.25 (469.33) | pM | 0 / 0 | | | nmol/g |
| Phytoestrogens | Secoisolariciresinol | 68 / 70 | 136.91 (153.28) | 285.71 (1199.04) | pM | 51 / 55 | 12.99 (38.89) | 13.87 (64.76) | nmol/g |
| Phytoestrogens | Secoisolariciresinol diglucoside | 0 / 0 | | | µM | 0 / 0 | | | nmol/g |
| Phytoestrogens | Enterodiol | 68 / 70 | 804 (271.8) | 1035.4 (1192.9) | pM | 69 / 70 | 28.35 (38.58) | 16.62 (36.85) | nmol/g |
| Phytoestrogens | Enterolactone | 68 / 70 | 244.88 (445.9) | 301.22 (418.24) | pM | 69 / 70 | 354.96 (175.22) | 345.26 (172.54) | nmol/g |
| Phytoestrogens | Matairesinol | 68 / 70 | 115.6 (107.13) | 314.45 (834.18) | pM | 69 / 70 | 2.39 (6.32) | 0.83 (1.09) | nmol/g |
| Steroids | Corticosterone | 68 / 70 | 2.81 (2.16) | 3.26 (2.28) | µg/L | 46 / 51 | 2.14 (3.94) | 1.56 (2.03) | ng/g |
| Steroids | 18-Hydroxycorticosterone | | *Not found* | | | 65 / 68 | 23.2 (33) | 30.67 (95.05) | ng/g |
| Steroids | 11-Deoxycorticosterone | 68 / 70 | 0.03 (0.01) | 0.04 (0.03) | µg/L | 68 / 70 | 3.5 (4.88) | 4.59 (11.3) | ng/g |
| Steroids | Tetrahydrodeoxycorticosterone | | *Not found* | | | 0 / 0 | | | ng/g |
| Steroids | Cortisol | 68 / 70 | 88.88 (31.49) | 97.65 (28.79) | µg/L | 36 / 26 | 0.25 (0.39) | 0.16 (0.48) | ng/g |
| Steroids | 6b-Hydroxycortisol | 68 / 70 | 1.58 (0.63) | 1.65 (0.64) | µg/L | 0 / 0 | | | ng/g |
| Steroids | 18-Hydroxycortisol | | *Not found* | | | 62 / 59 | 1.58 (2.12) | 1.56 (3.99) | ng/g |
| Steroids | 11-Deoxycortisol | 68 / 70 | 1.51 (0.98) | 1.59 (1.34) | µg/L | 69 / 70 | 0.72 (0.63) | 0.56 (0.5) | ng/g |
| Steroids | 21-Deoxycortisol | 68 / 70 | 0.34 (0.34) | 0.33 (0.21) | µg/L | 62 / 66 | 28.5 (34.52) | 35.1 (79.02) | ng/g |
| Steroids | 18-Oxocortisol | | *Not found* | | | 48 / 55 | 2.82 (5.05) | 2.85 (5.08) | ng/g |
| Steroids | Cortisone | 68 / 70 | 26.1 (6.78) | 26.65 (6.85) | µg/L | 52 / 53 | 0.89 (1.31) | 1.6 (6.39) | ng/g |
| Steroids | Uricortisone | 68 / 70 | 2.93 (1.94) | 3.36 (1.98) | µg/L | 0 / 0 | | | ng/g |
| Steroids | Pregnanediol 3-glucuronide | 68 / 70 | 0.01 (0.01) | 0.04 (0.15) | µM | 0 / 0 | | | ng/g |
| Steroids | Pregnenolone | 68 / 70 | 0.26 (0.28) | 0.24 (0.16) | µg/L | 69 / 70 | 615.39 (1077.12) | 432.42 (423.85) | ng/g |
| Steroids | Pregnenolone 3-sulfate | 68 / 70 | 0.04 (0.04) | 0.04 (0.02) | µM | 42 / 27 | 1.2 (2.44) | 0.57 (1.54) | ng/g |
| Steroids | 17a-Hydroxypregnenolone | 68 / 70 | 0.22 (0.12) | 0.28 (0.35) | µg/L | 68 / 67 | 1.03 (1.93) | 1.08 (2.44) | ng/g |
| Steroids | Progesterone | 68 / 70 | 0.03 (0.06) | 0.04 (0.04) | µg/L | 69 / 70 | 30.3 (44.05) | 54.84 (149.91) | ng/g |
| Steroids | 17a-Hydroxyprogesterone | 67 / 68 | 0.29 (0.32) | 0.28 (0.25) | µg/L | 68 / 69 | 4.02 (5.09) | 4.35 (5.59) | ng/g |
| Steroids | Aldosterone | 68 / 70 | 0.07 (0.03) | 0.09 (0.06) | µg/L | 0 / 0 | | | ng/g |
| Steroids | Androsterone 3-sulfate | 68 / 70 | 0.76 (0.54) | 0.57 (0.45) | µM | 63 / 58 | 0.88 (2.21) | 0.7 (3.13) | ng/g |
| Steroids | Androstenedione | 68 / 70 | 0.3 (0.14) | 0.28 (0.14) | µg/L | 0 / 0 | | | ng/g |
| Steroids | DHEA | 68 / 70 | 3.52 (2.32) | 3.29 (2.05) | µg/L | 68 / 68 | 19.12 (36.58) | 14.63 (30.47) | ng/g |
| Steroids | DHEA-sulfate | 68 / 70 | 1.81 (1.15) | 1.47 (0.94) | µM | 69 / 70 | 821.2 (888.2) | 410.42 (708.14) | ng/g |
| Steroids | Testosterone | 68 / 70 | 0.2 (0.09) | 0.19 (0.11) | µg/L | | *Not found* | | |
| Steroids | Dihydrotestosterone | 68 / 70 | 0.03 (0.01) | 0.02 (0.01) | µg/L | 60 / 62 | 0.78 (0.74) | 0.74 (0.79) | ng/g |
| Sterol | Cholesterol | 68 / 70 | 176241 (64762) | 189173 (69160) | nM | 69 / 70 | 366302 (326583) | 274085 (293117) | nmol/g |
| Sterol | Cholesterol 3-sulfate | 68 / 70 | 0.56 (0.45) | 0.65 (0.46) | µM | 69 / 70 | 2984.11 (1571.5) | 2489.06 (1311.84) | nmol/g |
| Sterol | 7-Dehydrocholesterol | 61 / 66 | 11.05 (8.77) | 15.07 (18.46) | nM | 69 / 70 | 512.03 (526.79) | 344.78 (336.5) | nmol/g |
| Sterol | Desmosterol | 68 / 70 | 122.57 (51.56) | 140.45 (70.46) | nM | 69 / 70 | 975.26 (871.4) | 1032.5 (1691.46) | nmol/g |
| Sterol | 7-Dehydrodesmosterol | 68 / 70 | 117.25 (686) | 38.84 (25.74) | nM | 62 / 69 | 27.96 (31.97) | 35.62 (39.11) | nmol/g |
| Sterol | Dihydrolanosterol | 68 / 70 | 4.38 (3.68) | 4.21 (2.64) | nM | 69 / 70 | 1347.46 (1275.33) | 1048.01 (1071.02) | nmol/g |
| Sterol | Dihydrolathosterol | 68 / 70 | 294.27 (146.22) | 332.71 (186.08) | nM | 68 / 70 | 237967 (238093) | 285682 (214627) | nmol/g |
| Sterol | Lanosterol | 68 / 70 | 29.82 (17.47) | 32.87 (19.17) | nM | 69 / 70 | 11975.07 (6811.49) | 12558.86 (5658.52) | nmol/g |
| Sterol | Lathosterol | 68 / 70 | 373.14 (181.8) | 412.82 (238.68) | nM | 69 / 70 | 5125.105 (3063.38) | 4295.75 (2317.49) | nmol/g |
| Sterol | Zymostenol | 68 / 70 | 100.52 (64.35) | 91.47 (48.81) | nM | 69 / 70 | 8994.91 (5315.36) | 8309.17 (4505.54) | nmol/g |
| Sterol | Zymosterol | 68 / 70 | 52.04 (30.57) | 49.51 (26.01) | nM | 69 / 70 | 484.38 (538.74) | 532.55 (1116.97) | nmol/g |
| Bile acids | 12-Ketochenodeoxycholic acid | 0 / 0 | | | µM | 61 / 57 | 20.82 (73.81) | 1.85 (8.27) | nmol/g |

| | | | | | | | | | |
|---|---|---|---|---|---|---|---|---|---|
| Bile acids | 12-Ketolithocholic acid | 64 / 68 | 0.02 (0.02) | 0.02 (0.02) | µM | 69 / 69 | 48.28 (62.22) | 37.27 (58.81) | nmol/g |
| Bile acids | 3-Oxocholic acid | 0 / 0 | | | µM | 69 / 70 | 93.61 (273.82) | 18.15 (69.57) | nmol/g |
| Bile acids | 3beta-OH-5-cholestenoic acid | 68 / 70 | 0.67 (0.18) | 0.69 (0.16) | µM | 66 / 68 | 1.1 (1.82) | 0.74 (1.1) | nmol/g |
| Bile acids | 3beta,7alfa-DiOH-5-cholestenoic acid | 68 / 70 | 0.05 (0.02) | 0.05 (0.02) | µM | 67 / 65 | 0.48 (0.32) | 0.45 (0.7) | nmol/g |
| Bile acids | 6,7-Diketolithocholic acid | 0 / 0 | | | µM | 0 / 0 | | | nmol/g |
| Bile acids | 7-Ketodeoxycholic acid | 25 / 18 | 0 (0) | 0 (0) | µM | 69 / 63 | 23.62 (83.27) | 2.32 (10.48) | nmol/g |
| Bile acids | 7-Ketolithocholic acid | 0 / 0 | | | µM | 69 / 70 | 822.95 (704.16) | 681.48 (571.16) | nmol/g |
| Bile acids | 7alfa-OH-3-oxo-4-cholestenoic acid | 68 / 70 | 0.15 (0.05) | 0.14 (0.04) | µM | 41 / 50 | 0.08 (0.12) | 0.1 (0.17) | nmol/g |
| Bile acids | alfa-Muricholic acid | 0 / 0 | | | µM | 48 / 56 | 0.66 (1.54) | 0.47 (0.47) | nmol/g |
| Bile acids | Allocholic acid | 47 / 50 | 0.003 (0.01) | 0.01 (0.02) | µM | 69 / 70 | 10.01 (13.46) | 7.12 (8.28) | nmol/g |
| Bile acids | Allocholic acid 3-sulfate | 0 / 0 | | | µM | | *Not found* | | |
| Bile acids | Alloisolithocholic acid | 67 / 70 | 0.04 (0.03) | 0.04 (0.02) | µM | 69 / 66 | 38.71 (56.75) | 53.07 (61.73) | nmol/g |
| Bile acids | Apocholic acid | 68 / 69 | 0.01 (0.02) | 0.01 (0.01) | µM | 69 / 70 | 126.65 (376.37) | 28.99 (78.25) | nmol/g |
| Bile acids | beta-Muricholic acid | 0 / 0 | | | µM | 67 / 68 | 3.6 (9.48) | 0.99 (1.61) | nmol/g |
| Bile acids | Chenodeoxycholic acid | 68 / 70 | 0.44 (0.62) | 0.85 (2.62) | µM | 69 / 70 | 246.19 (723.15) | 57.81 (170.92) | nmol/g |
| Bile acids | Chenodeoxycholic acid 24-glucuronic | 0 / 0 | | | µM | 4 / 1 | 0.01 (0.05) | 0.0003 (0.002) | nmol/g |
| Bile acids | Chenodeoxycholic acid 3-glucuronide | 0 / 0 | | | µM | 0 / 0 | | | nmol/g |
| Bile acids | Chenodeoxycholic acid 3-sulfate | 68 / 67 | 0.01 (0.02) | 0.02 (0.03) | µM | 40 / 27 | 16.83 (51.73) | 4.66 (12.93) | nmol/g |
| Bile acids | Cholic acid | 68 / 70 | 0.16 (0.31) | 0.38 (0.94) | µM | 69 / 70 | 350.33 (952.47) | 67.01 (249.53) | nmol/g |
| Bile acids | Cholic acid 3-sulfate | 0 / 0 | | | µM | 39 / 19 | 35.43 (103.02) | 7.34 (30.44) | nmol/g |
| Bile acids | Cholic acid 7-sulfate | 0 / 0 | | | µM | 55 / 60 | 0.23 (0.44) | 0.2 (0.28) | nmol/g |
| Bile acids | Dehydrocholic acid | 66 / 64 | 0.01 (0.01) | 0.01 (0.01) | µM | 32 / 24 | 2.01 (9.12) | 0.12 (0.29) | nmol/g |
| Bile acids | Dehydrolithocholic acid | 66 / 69 | 0.01 (0.01) | 0.01 (0.01) | µM | 69 / 70 | 500.92 (333.56) | 424.52 (254.08) | nmol/g |
| Bile acids | Deoxycholic acid | 68 / 70 | 0.61 (0.57) | 0.73 (0.79) | µM | 69 / 70 | 1454.42 (1403.78) | 1399.93 (1880.47) | nmol/g |
| Bile acids | Deoxycholic acid 24-glucuronide | 0 / 0 | | | µM | 1 / 0 | 0.26 (2.14) | 0 (0) | nmol/g |
| Bile acids | Deoxycholic acid 3-glucuronide | 0 / 0 | | | µM | 0 / 0 | | | nmol/g |
| Bile acids | Deoxycholic acid 3-sulfate | 0 / 0 | | | µM | 68 / 67 | 164.82 (368.57) | 57.78 (216.58) | nmol/g |
| Bile acids | DHCA | 0 / 0 | | | µM | 69 / 70 | 2.04 (1.88) | 1.68 (0.94) | nmol/g |
| Bile acids | Dioxolithocholic acid | 0 / 0 | | | µM | 69 / 70 | 27.2 (83.2) | 7.4 (7.7) | nmol/g |
| Bile acids | Glyco-alfa-muricholic acid | 54 / 60 | 0 (0) | 0 (0) | µM | 0 / 0 | | | nmol/g |
| Bile acids | Glyco-beta-muricholic acid | 0 / 0 | | | µM | 0 / 0 | | | nmol/g |
| Bile acids | Glyco-omega-muricholic acid | 68 / 70 | 0.01 (0.01) | 0.02 (0.02) | µM | 0 / 0 | | | nmol/g |
| Bile acids | Glycoallocholic acid | 68 / 70 | 0.01 (0.01) | 0.01 (0.01) | µM | 14 / 12 | 0.04 (0.13) | 0.01 (0.04) | nmol/g |
| Bile acids | Glycochenodeoxycholic acid | 68 / 70 | 0.68 (0.77) | 0.52 (0.47) | µM | 69 / 70 | 3.1 (6.91) | 2.4 (4.79) | nmol/g |
| Bile acids | Glycochenodeoxycholic acid 3-sulfate | 66 / 70 | 0.33 (0.31) | 0.4 (0.38) | µM | 26 / 10 | 0.35 (1.28) | 0.07 (0.27) | nmol/g |
| Bile acids | Glycochenodeoxycholic acid 7-sulfate | 68 / 70 | 1.59 (1.25) | 1.53 (1.29) | µM | | *Not found* | | |
| Bile acids | Glycocholic acid | 68 / 70 | 0.17 (0.24) | 0.13 (0.12) | µM | 69 / 70 | 1.97 (3.9) | 1.38 (2.52) | nmol/g |
| Bile acids | Glycocholic acid 3-sulfate | 0 / 0 | | | µM | 5 / 3 | 0.01 (0.05) | 0.0005 (0.003) | nmol/g |
| Bile acids | Glycodehydrocholic acid | 0 / 0 | | | µM | 0 / 0 | | | nmol/g |
| Bile acids | Glycodeoxycholic acid | 68 / 70 | 0.27 (0.23) | 0.31 (0.33) | µM | 69 / 70 | 3.62 (6.06) | 2.76 (4.22) | nmol/g |
| Bile acids | Glycodeoxycholic acid 3-sulfate | 0 / 0 | | | µM | 51 / 46 | 0.34 (1.273) | 0.08 (0.209) | nmol/g |
| Bile acids | Glycohyocholic acid | 68 / 70 | 0.01 (0.01) | 0.01 (0.01) | µM | 8 / 7 | 0.01 (0.02) | 0.001 (0.01) | nmol/g |
| Bile acids | Glycohyodeoxycholic acid | 0 / 0 | | | µM | 3 / 4 | 0.002 (0.01) | 0.003 (0.02) | nmol/g |
| Bile acids | Glycolithocholic acid | 68 / 70 | 0.02 (0.03) | 0.02 (0.02) | µM | 69 / 70 | 0.28 (0.2) | 0.27 (0.27) | nmol/g |
| Bile acids | Glycolithocholic acid 3-sulfate | 67 / 70 | 1.24 (0.88) | 1.27 (0.61) | µM | 66 / 67 | 5.74 (18.766) | 1.22 (2.4) | nmol/g |
| Bile acids | Glycoursodeoxycholic acid | 68 / 70 | 0.11 (0.17) | 0.05 (0.05) | µM | 68 / 68 | 0.56 (1.77) | 0.25 (0.54) | nmol/g |
| Bile acids | Glycoursodeoxycholic acid 3-sulfate | 66 / 68 | 0.01 (0.005) | 0.01 (0.005) | µM | 20 / 11 | 0.12 (0.74) | 0.01 (0.04) | nmol/g |
| Bile acids | Hyocholic acid | 0 / 0 | | | µM | 62 / 61 | 4.18 (10.11) | 1.15 (2.85) | nmol/g |
| Bile acids | Hyocholic acid 3-sulfate | 67 / 66 | 0.06 (0.04) | 0.04 (0.03) | µM | | *Not found* | | |
| Bile acids | Hyodeoxycholic acid | 64 / 66 | 0.01 (0.004) | 0.01 (0.01) | µM | 69 / 70 | 121.07 (426.18) | 90.61 (125.33) | nmol/g |
| Bile acids | Hyodeoxycholic acid 3-sulfate | 68 / 70 | 0.04 (0.07) | 0.03 (0.02) | µM | | *Not found* | | |
| Bile acids | Isodeoxycholic acid | 0 / 0 | | | µM | 0 / 0 | | | nmol/g |
| Bile acids | Isolithocholic acid | 65 / 69 | 0.03 (0.02) | 0.03 (0.02) | µM | 69 / 70 | 343.23 (192.24) | 306.13 (144.61) | nmol/g |
| Bile acids | Isolithocholic acid 3-sulfate | 0 / 0 | | | µM | 67 / 62 | 13.05 (32.63) | 4.19 (12.05) | nmol/g |
| Bile acids | Lithocholic acid | 68 / 70 | 0.05 (0.04) | 0.06 (0.03) | µM | 69 / 70 | 1046.37 (512.93) | 854.79 (405.04) | nmol/g |
| Bile acids | Lithocholic acid 24-glucuronide | 0 / 0 | | | µM | 0 / 0 | | | nmol/g |
| Bile acids | Lithocholic acid 3-glucuronide | 0 / 0 | | | µM | 0 / 0 | | | nmol/g |
| Bile acids | Lithocholic acid 3-sulfate | 67 / 70 | 0.005 (0.01) | 0 (0.01) | µM | 68 / 69 | 157.3 (380.4) | 42.88 (175.71) | nmol/g |
| Bile acids | Murocholic acid | 68 / 70 | 0.32 (0.61) | 0.11 (0.12) | µM | 69 / 70 | 5.87 (8.37) | 3.32 (3.56) | nmol/g |
| Bile acids | Norcholic acid | 0 / 0 | | | µM | 65 / 69 | 2.13 (2.41) | 1.18 (1.09) | nmol/g |
| Bile acids | Nordeoxycholic acid | 68 / 70 | 0.01 (0.01) | 0.01 (0.01) | µM | 64 / 69 | 0.54 (0.58) | 0.73 (0.88) | nmol/g |
| Bile acids | Norursodeoxycholic acid | | *Not found* | | | 60 / 51 | 0.26 (0.32) | 0.17 (0.23) | nmol/g |
| Bile acids | omega-Muricholic acid | 0 / 0 | | | µM | 51 / 42 | 27.15 (208.7) | 0.44 (0.92) | nmol/g |
| Bile acids | Tauro-alfa-Muricholic acid | 0 / 0 | | | µM | 10 / 4 | 1.2 (9.42) | 0.02 (0.12) | nmol/g |
| Bile acids | Tauro-beta-muricholic acid | 0 / 0 | | | µM | 0 / 0 | | | nmol/g |
| Bile acids | Tauro-omega-muricholic acid | 65 / 70 | 0.01 (0.01) | 0.01 (0.02) | µM | 2 / 1 | 0.001 (0.01) | 0.001 (0.01) | nmol/g |
| Bile acids | Tauroallocholic acid | 0 / 0 | | | µM | 0 / 0 | | | nmol/g |
| Bile acids | Taurochenodeoxycholic acid 3-sulfate | 58 / 64 | 0.01 (0.01) | 0.01 (0.01) | µM | 4 / 1 | 0.133 (0.74) | 0.02 (0.21) | nmol/g |
| Bile acids | Taurochenodexycholic acid | 68 / 70 | 0.05 (0.07) | 0.07 (0.08) | µM | 69 / 70 | 1.53 (4.75) | 0.99 (1.99) | nmol/g |
| Bile acids | Taurocholic acid | 68 / 70 | 0.03 (0.07) | 0.03 (0.06) | µM | 68 / 70 | 1.232 (5.59) | 0.4 (0.89) | nmol/g |
| Bile acids | Taurodehydrocholic acid | 0 / 0 | | | µM | 0 / 0 | | | nmol/g |
| Bile acids | Taurodeoxycholic acid | 67 / 70 | 0.05 (0.07) | 0.07 (0.11) | µM | 66 / 70 | 2.56 (7.12) | 1.44 (5.83) | nmol/g |
| Bile acids | Taurodeoxycholic acid 3-sulfate | 62 / 69 | 0.01 (0.01) | 0.02 (0.02) | µM | 6 / 2 | 0.352 (1.427) | 0.15 (1.24) | nmol/g |
| Bile acids | Taurohyocholic acid | 60 / 63 | 0.001 (0.001) | 0 (0) | µM | 1 / 1 | 0.004 (0.03) | 0.001 (0.01) | nmol/g |
| Bile acids | Taurolithocholic acid | 66 / 70 | 0.004 (0.01) | 0.01 (0.01) | µM | 55 / 58 | 0.54 (1.35) | 0.39 (1.99) | nmol/g |
| Bile acids | Taurolithocholic acid 3-sulfate | 64 / 69 | 0.08 (0.09) | 0.1 (0.09) | µM | 6 / 2 | 0.49 (3.39) | 0.07 (0.52) | nmol/g |
| Bile acids | Tauroisolithocholic acid | | *Not found* | | | 0 / 0 | | | nmol/g |
| Bile acids | Tauroursodeoxycholic acid 3-sulfate | 26 / 17 | 0.003 (0.01) | 0 (0) | µM | 3 / 1 | 0.108 (0.763) | 0.01 (0.11) | nmol/g |
| Bile acids | Tauroursodexycholic acid | 68 / 70 | 0.003 (0.005) | 0 (0) | µM | 68 / 68 | 0.46 (2.3) | 0.11 (0.28) | nmol/g |
| Bile acids | THCA | 53 / 45 | 0.002 (0.002) | 0 (0) | µM | 69 / 70 | 12.25 (14.37) | 8.38 (14.48) | nmol/g |
| Bile acids | Ursocholic acid | 63 / 60 | 0.002 (0.003) | 0 (0) | µM | 69 / 69 | 199.71 (581.27) | 48.45 (168.68) | nmol/g |
| Bile acids | Ursodeoxycholic acid | 68 / 70 | 0.08 (0.11) | 0.09 (0.09) | µM | 36 / 24 | 99.06 (259.25) | 43.25 (163.83) | nmol/g |
| Bile acids | Ursodeoxycholic acid 24-glucuronide | 0 / 0 | | | µM | 0 / 0 | | | nmol/g |
| Bile acids | Ursodeoxycholic acid 3-glucuronide | 67 / 67 | 0.05 (0.03) | 0.05 (0.04) | µM | 0 / 0 | | | nmol/g |
| Bile acids | Ursodeoxycholic acid 3-sulfate | 0 / 0 | | | µM | 57 / 50 | 185.27 (395.21) | 84.07 (234.78) | nmol/g |

| | | | | | | | | | |
|---|---|---|---|---|---|---|---|---|---|
| Bile acids | Ursodeoxycholic acid 7-sulfate | 67 / 68 | 0.01 (0.01) | 0.01 (0.01) | μM | 59 / 47 | 60.05 (170.91) | 4.8 (19.35) | nmol/g |
| Carnitines | L-Carnitine | 68 / 70 | 31.25 (7.78) | 31.17 (5.69) | μM | 69 / 70 | 34.54 (84.12) | 15.45 (18.39) | nmol/g |
| Carnitines | 2-Methylbutyryl-L-carnitine | | *Not found* | | | 69 / 70 | 15.59 (6.87) | 13.54 (4.38) | nmol/g |
| Carnitines | 3-Methylcrotonyl-L-carnitine | 68 / 70 | 0.00407 (0.0016) | 0.00404 (0.0016) | μM | 62 / 57 | 0.05 (0.17) | 0.01 (0.01) | nmol/g |
| Carnitines | 3-Methylglutaryl-carnitine | 68 / 70 | 0.0092 (0.013) | 0.0059 (0.003) | μM | 11 / 8 | 0.15 (0.59) | 0.03 (0.14) | nmol/g |
| Carnitines | Acetyl-carnitine | 68 / 70 | 10.77 (3.62) | 9.47 (2.97) | μM | 69 / 70 | 12.505 (74.33) | 0.78 (1.17) | nmol/g |
| Carnitines | Adipoyl-carnitine | 68 / 70 | 0.017 (0.016) | 0.019 (0.013) | μM | 50 / 27 | 0.1 (0.17) | 0.04 (0.08) | nmol/g |
| Carnitines | Arachidonoyl-carnitine | 68 / 70 | 0.0234 (0.022) | 0.0199 (0.012) | μM | 68 / 61 | 0.46 (1.23) | 0.13 (0.17) | nmol/g |
| Carnitines | Butyryl-carnitine | 68 / 70 | 0.102 (0.0457) | 0.0979 (0.0369) | μM | 69 / 70 | 0.42 (1.06) | 0.12 (0.1) | nmol/g |
| Carnitines | Decanoyl-carnitine | 68 / 70 | 0.230 (0.17) | 0.199 (0.15) | μM | 69 / 70 | 0.27 (1.46) | 0.03 (0.05) | nmol/g |
| Carnitines | Dodecanoyl-carnitine | 68 / 70 | 0.0611 (0.034) | 0.0567 (0.035) | μM | 69 / 70 | 1.61 (8.11) | 0.25 (0.91) | nmol/g |
| Carnitines | Glutaryl-carnitine | 68 / 70 | 0.0743 (0.028) | 0.0645 (0.023) | μM | 31 / 17 | 0.39 (1.65) | 0.05 (0.12) | nmol/g |
| Carnitines | Hexanoyl-carnitine | 68 / 70 | 0.0482 (0.0286) | 0.0378 (0.0229) | μM | 69 / 70 | 0.04 (0.12) | 0.02 (0.02) | nmol/g |
| Carnitines | Isobutyryl-carnitine | | *Not found* | | | 69 / 70 | 0.47 (2.2) | 0.07 (0.08) | nmol/g |
| Carnitines | Isovaleryl-carnitine | | *Not found* | | | 69 / 70 | 0.76 (2.62) | 0.18 (0.2) | nmol/g |
| Carnitines | Linoleyl-carnitine | 68 / 70 | 0.0235 (0.0127) | 0.0192 (0.009) | μM | 69 / 70 | 0.99 (2.61) | 0.23 (0.52) | nmol/g |
| Carnitines | Malonyl-carnitine | 68 / 70 | 0.0131 (0.0026) | 0.0128 (0.00195) | μM | 69 / 70 | 1.62 (2.31) | 1.07 (0.68) | nmol/g |
| Carnitines | Methylmalonyl-carnitine | 68 / 70 | 0.0078 (0.0016) | 0.0077 (0.0017) | μM | 69 / 70 | 15.14 (79.58) | 3.28 (2.77) | nmol/g |
| Carnitines | Myristoyl-carnitine | 68 / 70 | 0.0255 (0.0103) | 0.0249 (0.0097) | μM | 69 / 70 | 2.5 (8.28) | 0.56 (1.78) | nmol/g |
| Carnitines | Octanoyl-carnitine | 68 / 70 | 0.133 (0.099) | 0.110 (0.087) | μM | 69 / 70 | 0.1 (0.29) | 0.11 (0.11) | nmol/g |
| Carnitines | Oleoyl-carnitine | 68 / 70 | 0.3661 (0.163) | 0.3186 (0.19) | μM | 69 / 70 | 34.22 (73.01) | 8.89 (20.3) | nmol/g |
| Carnitines | Palmitoyl-carnitine | 68 / 70 | 0.1065(0.0345) | 0.099 (0.0279) | μM | 69 / 70 | 22.18 (66.17) | 5.14 (14.75) | nmol/g |
| Carnitines | Propionyl-carnitine | 68 / 70 | 0.328 (0.119) | 0.322 (0.129) | μM | 69 / 70 | 1.2 (3.32) | 0.48 (0.65) | nmol/g |
| Carnitines | Stearoyl-carnitine | 68 / 70 | 0.0407 (0.0142) | 0.0397 (0.0106) | μM | 69 / 70 | 16.53 (20.02) | 7.73 (10.34) | nmol/g |
| Carnitines | Succinyl-carnitine | 68 / 70 | 0.0271 (0.006) | 0.0257 (0.006) | μM | 69 / 70 | 2.95 (4.83) | 1.31 (1.54) | nmol/g |
| Carnitines | Valeryl-carnitine | 68 / 70 | 0.0461 (0.0162) | 0.0477 (0.0241) | μM | 69 / 70 | 0.21 (0.23) | 0.17 (0.1) | nmol/g |
| Lipids | LysoPC(14:0) | 68 / 70 | 2.02 (0.99) | 2.27 (0.91) | μM | 69 / 70 | 6.83 (17.85) | 2.47 (3.33) | nmol/g |
| Lipids | LysoPC(15:0) | 68 / 70 | 1.13 (0.46) | 1.31 (0.45) | μM | 69 / 70 | 7.26 (21.53) | 3.85 (5.2) | nmol/g |
| Lipids | LysoPC(16:0) | 68 / 70 | 78.91 (8.33) | 78.46 (7.65) | μM | 69 / 70 | 824.34 (1350.95) | 466.92 (573.62) | nmol/g |
| Lipids | LysoPC(16:1) | 68 / 70 | 4.74 (2.02) | 5.21 (2.35) | μM | 69 / 70 | 15.78 (56.61) | 3.94 (4.9) | nmol/g |
| Lipids | LysoPC(17:0) | 68 / 70 | 0.5 (0.04) | 0.5 (0.04) | μM | 69 / 70 | 110.88 (49.49) | 96.14 (33.15) | nmol/g |
| Lipids | LysoPC(18:0) | 68 / 70 | 99.54 (19.1) | 100.53 (14.74) | μM | 69 / 70 | 359.53 (883.21) | 164.75 (207.65) | nmol/g |
| Lipids | LysoPC(18:1) | 68 / 70 | 40.99 (9.41) | 41.59 (9) | μM | 69 / 70 | 192.51 (256.17) | 142.37 (219.92) | nmol/g |
| Lipids | LysoPC(18:2) | 68 / 70 | 47.72 (11.84) | 47.83 (11.61) | μM | 69 / 70 | 202 (433.34) | 107.83 (202.92) | nmol/g |
| Lipids | LysoPC(19:0) | 68 / 70 | 0.3 (0.17) | 0.31 (0.1) | μM | 69 / 70 | 4.58 (13.88) | 2.51 (6.03) | nmol/g |
| Lipids | LysoPC(20:4) | 68 / 70 | 11.58 (4.16) | 11.37 (3.88) | μM | 69 / 70 | 1.54 (4.36) | 0.61 (0.83) | nmol/g |
| Lipids | LysoPC(20:5) | 68 / 70 | 1.96 (1.37) | 2.45 (1.99) | μM | 58 / 50 | 0.26 (0.55) | 0.11 (0.21) | nmol/g |
| Lipids | LysoPE(14:0) | 68 / 70 | 0.03 (0.02) | 0.04 (0.04) | μM | 69 / 70 | 183.15 (187.32) | 154.75 (125.04) | nmol/g |
| Lipids | LysoPE(15:0) | 68 / 70 | 0.61 (0.07) | 0.59 (0.07) | μM | 69 / 70 | 95.3 (41.55) | 81.76 (25.61) | nmol/g |
| Lipids | LysoPE(16:0) | 68 / 70 | 3.55 (1.07) | 3.72 (1.15) | μM | 69 / 70 | 2315.1 (1983.93) | 1766.57 (1500.62) | nmol/g |
| Lipids | LysoPE(17:0) | 68 / 70 | 0.13 (0.06) | 0.14 (0.05) | μM | 69 / 70 | 125.6 (118.27) | 94.51 (72.72) | nmol/g |
| Lipids | LysoPE(18:0) | 68 / 70 | 4.56 (1.49) | 4.63 (1.34) | μM | 69 / 70 | 130.72 (173.07) | 90.29 (76.98) | nmol/g |
| Lipids | LysoPE(18:1) | 68 / 70 | 5.33 (3.15) | 4.88 (2.86) | μM | 69 / 70 | 221.13 (158.049) | 210.11 (174.046) | nmol/g |
| Lipids | LysoPE(18:2) | 68 / 70 | 7.04 (4.3) | 6.47 (3.38) | μM | 69 / 70 | 192.06 (278.29) | 140.45 (257.68) | nmol/g |
| Lipids | LysoPE(19:0) | 68 / 70 | 0.01 (0.01) | 0.01 (0.01) | μM | 64 / 66 | 1.6 (2.38) | 1.34 (2.67) | nmol/g |
| Lipids | LysoPE(20:3) | 68 / 70 | 0.37 (0.15) | 0.36 (0.16) | μM | 69 / 70 | 3.34 (5.588) | 2.39 (3.123) | nmol/g |
| Lipids | LysoPE(20:4) | 68 / 70 | 2.46 (0.65) | 2.34 (0.78) | μM | 69 / 70 | 6.49 (11.06) | 3.66 (3.43) | nmol/g |
| Lipids | LysoPE(20:5) | 68 / 70 | 0.28 (0.18) | 0.35 (0.26) | μM | 69 / 70 | 1.92 (2.21) | 1.84 (3.18) | nmol/g |
| Lipids | PAF C-16 | 68 / 70 | 0.37 (0.15) | 0.36 (0.11) | μM | 0 / 0 | | | nmol/g |
| Lipids | Sphinganine (d18:0) | 68 / 70 | 0.06 (0.04) | 0.07 (0.04) | μM | 32 / 31 | 7.16 (9.17) | 6.11 (7.4) | nmol/g |
| Lipids | Sphinganine-1P (d18:0) | 68 / 70 | 0.64 (0.38) | 0.66 (0.31) | μM | 69 / 70 | 128.62 (266.79) | 55.1 (106.43) | nmol/g |
| Lipids | Sphingosine (d18:1) | 68 / 70 | 0.01 (0.005) | 0.01 (0) | μM | 12 / 13 | 8.61 (30.3) | 7 (19.79) | nmol/g |
| Lipids | Sphingosine-1P (d18:1) | 68 / 70 | 0.02 (0.01) | 0.03 (0.02) | μM | 0 / 0 | | | nmol/g |
| SCFAs | 2-Methylbutyric acid | 68 / 70 | 0.05 (0.02) | 0.04 (0.02) | μM | 69 / 70 | 65966 (66194) | 76944 (70927) | nmol/g |
| SCFAs | 3-Methylvaleric acid | 0 / 0 | | | μM | 67 / 69 | 206.83 (272.72) | 187.74 (295.71) | nmol/g |
| SCFAs | Acetic acid | 68 / 70 | 36.32 (12.86) | 33.74 (20.2) | μM | 69 / 70 | 5308242 (7365761) | 4425901 (5949922) | nmol/g |
| SCFAs | Butyric acid | 68 / 70 | 0.54 (0.38) | 0.62 (0.35) | μM | 69 / 70 | 1020874 (1360648) | 1027972 (1628820) | nmol/g |
| SCFAs | Caproic acid | 68 / 70 | 0.72 (0.13) | 0.83 (0.26) | μM | 69 / 70 | 31478 (85451) | 51093 (82241) | nmol/g |
| SCFAs | Isobutyric acid | 68 / 70 | 0.27 (0.09) | 0.28 (0.09) | μM | 69 / 70 | 07567.42 (102073.74 | 16213.31 (104002.03 | nmol/g |
| SCFAs | Isocaproic acid | 0 / 0 | | | μM | 69 / 70 | 2406.83 (4909.74) | 1908.43 (2936.32) | nmol/g |
| SCFAs | Isovaleric acid | 68 / 70 | 0.23 (0.14) | 0.21 (0.16) | μM | 69 / 70 | 92038.68 (92059.89) | 101936.1 (93334.95) | nmol/g |
| SCFAs | Propionic acid | 68 / 70 | 1.83 (0.97) | 1.59 (0.69) | μM | 69 / 70 | 1641951 (2437815) | 1039974 (1449960) | nmol/g |
| SCFAs | Valeric acid | 68 / 70 | 0.1 (0.1) | 0.11 (0.06) | μM | 69 / 70 | 141767.86 (168375.5 | 38675.94 (150220.62 | nmol/g |
| Amino acids | Arginine | 68 / 70 | 49.12 (24.78) | 47.7 (20.17) | μM | 69 / 70 | 1083.95 (3646.73) | 622 (1941.05) | nmol/g |
| Amino acids | Isoleucine | 68 / 70 | 65.73 (19.88) | 65.56 (29.92) | μM | 69 / 70 | 36559.99 (20385.37) | 29695.83 (20757.92) | nmol/g |
| Amino acids | Kynurenic acid | 68 / 70 | 0.05 (0.02) | 0.05 (0.02) | μM | 69 / 70 | 102.35 (89.32) | 104.75 (91.9) | nmol/g |
| Amino acids | Kynurenine | 68 / 70 | 3.16 (0.84) | 3.31 (0.78) | μM | 69 / 70 | 23.4 (14.38) | 18.74 (11.5) | nmol/g |
| Amino acids | Leucine | 68 / 70 | 123.33 (29.67) | 126.2 (46.85) | μM | 69 / 70 | 49363.13 (26328.15) | 39375.75 (25193.85) | nmol/g |
| Amino acids | Methionine | 68 / 70 | 34.4 (9) | 38.86 (13.71) | μM | 69 / 70 | 21076.74 (14294.76) | 15639.3 (11757.5) | nmol/g |
| Amino acids | Phenylalanine | 68 / 70 | 37.03 (8.31) | 38.56 (8.17) | μM | 69 / 70 | 12306.91 (10635.12) | 8472.6 (5855.95) | nmol/g |
| Amino acids | Proline | 68 / 70 | 235.64 (70.13) | 229.25 (77.22) | μM | 69 / 70 | 19403.92 (12810.68) | 17067.88 (13588.18) | nmol/g |
| Amino acids | Tryptophan | 68 / 70 | 26.75 (5.2) | 29.13 (6.5) | μM | 69 / 70 | 1282.88 (1009.81) | 813.25 (717.51) | nmol/g |
| Amino acids | Tryptophan-betaine | 68 / 70 | 0.94 (1.27) | 0.92 (0.82) | μM | 69 / 70 | 10.33 (34.42) | 6.1 (20.06) | nmol/g |
| Amino acids | Tyrosine | 68 / 70 | 62.1 (14.94) | 66.45 (19.33) | μM | 69 / 70 | 18199.61 (11749.65) | 13925.5 (10963.79) | nmol/g |
| Uremic toxin | Indoxyl sulfate | 68 / 70 | 1.58 (1.06) | 1.81 (1.13) | μM | 6 / 2 | | | nmol/g |
| Uremic toxin | p-Cresol sulfate | 68 / 70 | 6.2 (4.29) | 7.7 (5.16) | μM | 69 / 70 | 18.54 (60.1) | 4.18 (7.28) | nmol/g |
| Uremic toxin | Phenyl sulfate | 68 / 70 | 1.41 (1.45) | 1.05 (0.7) | μM | 0 / 0 | | | nmol/g |
| Other | Biliverdin | 68 / 70 | 0.43 (0.22) | 0.51 (0.25) | μM | 60 / 64 | 38.89 (72.64) | 19.48 (29.46) | nmol/g |
| Other | Catechol sulfate | 0 / 0 | | | μM | | *Not found* | | |
| Other | Hypoxanthine | 68 / 70 | 139.45 (53.02) | 172.86 (72) | μM | 69 / 70 | 13380.17 (7081.34) | 9625 (5580.04) | nmol/g |
